# Whole-Genome Sequencing Pilot Study of the Central Asian Genetic Diversity Project Reveals Distinct Genetic Histories, Adaptive Processes, and Introgression Events

**DOI:** 10.1101/2025.08.26.25334450

**Authors:** Guanglin He, Haoran Su, Qiuxia Sun, Renkuan Tang, Qingxin Yang, Lintao Luo, Jie Zhong, Zhaxylyk Sabitov, Jing Cheng, Fengxiao Bu, Yu Lu, Chao Liu, Huijun Yuan, Lan-Hai Wei, Maxat Zhabagin, Mengge Wang

**Author notes:** Correspondence (G.H.); (M.Z.); (L.W.); (M.W.).

## Abstract

The underrepresentation of Central Asian genomic data has constrained our understanding of their demographic history and hindered advancements in precision medicine and health equity. Despite the region’s rich historical tapestry, characterized by numerous trans-Eurasian migrations following the advent of agriculture and pastoralism, the genetic contributions of ancient Eurasians to modern Central Asians remain poorly understood. To address this gap, we performed an anthropologically informed Central Asian Genomic Diversity Project and reported the results of pilot whole-genome sequencing work on 166 Central Asians and Afghanistan Hazaras (CAAH) from 20 populations to investigate their demographic history, local adaptation, medical relevance, and archaic introgression. Significant genetic differentiation among CAAH populations was revealed. Tajik, Karluks, Turkmen, and Uzbek individuals exhibited higher proportions of West Eurasian ancestry, whereas the Kyrgyz, Karakalpak, Uyghur, and Hazara populations presented increased ancestry related to ancient Northeast Asians. In contrast, Dungans demonstrated a predominance of East Asian-derived ancestry. Four Turkic-related genetic clusters corresponding to geographic distribution were identified, supporting the “Northeast Asia origin” hypothesis for Turkic groups. Additionally, two Indo-European genetic clines were detected, with Hazaras being notably isolated. Strong genetic affinities were observed between Hazaras and Altaic groups in Siberia and between Dungans and Sino-Tibetan-speaking East Asians, underscoring the impact of ancient long-distance migrations on Eurasian genetic diversity. The recent east-west admixture in CAAH was estimated to have occurred 23-31 generations ago, aligning with the Song and Yuan dynasties and the Mongol Empire period. The mutation spectra of candidate disease-causing variants and pharmacogenomic genes were characterized, indicating that differentiated demographic histories significantly influence the genetic architecture of diseases among different Central Asians. Differential post-admixture adaptation signatures identified in the four genetically distinct groups have substantial effects on immune, metabolic, neural, and physical traits. Shifts in subsistence strategies significantly shaped the genetic architecture of complex traits in Central Asians. Neanderthal-like sequences exhibited varying phenotypic effects across genetically distinct CAAH strains, including susceptibility to immune and psychiatric conditions in West Eurasian-biased CAAH individuals and drug metabolism in East Eurasian-biased CAAH individuals. Denisovan-like segments were primarily linked to type 2 diabetes, etc. This research on Central Asian genomic diversity enhances the understanding of their evolutionary history and admixture events, promoting health equity and advancing precision medicine initiatives.

**Graphical abstract:** 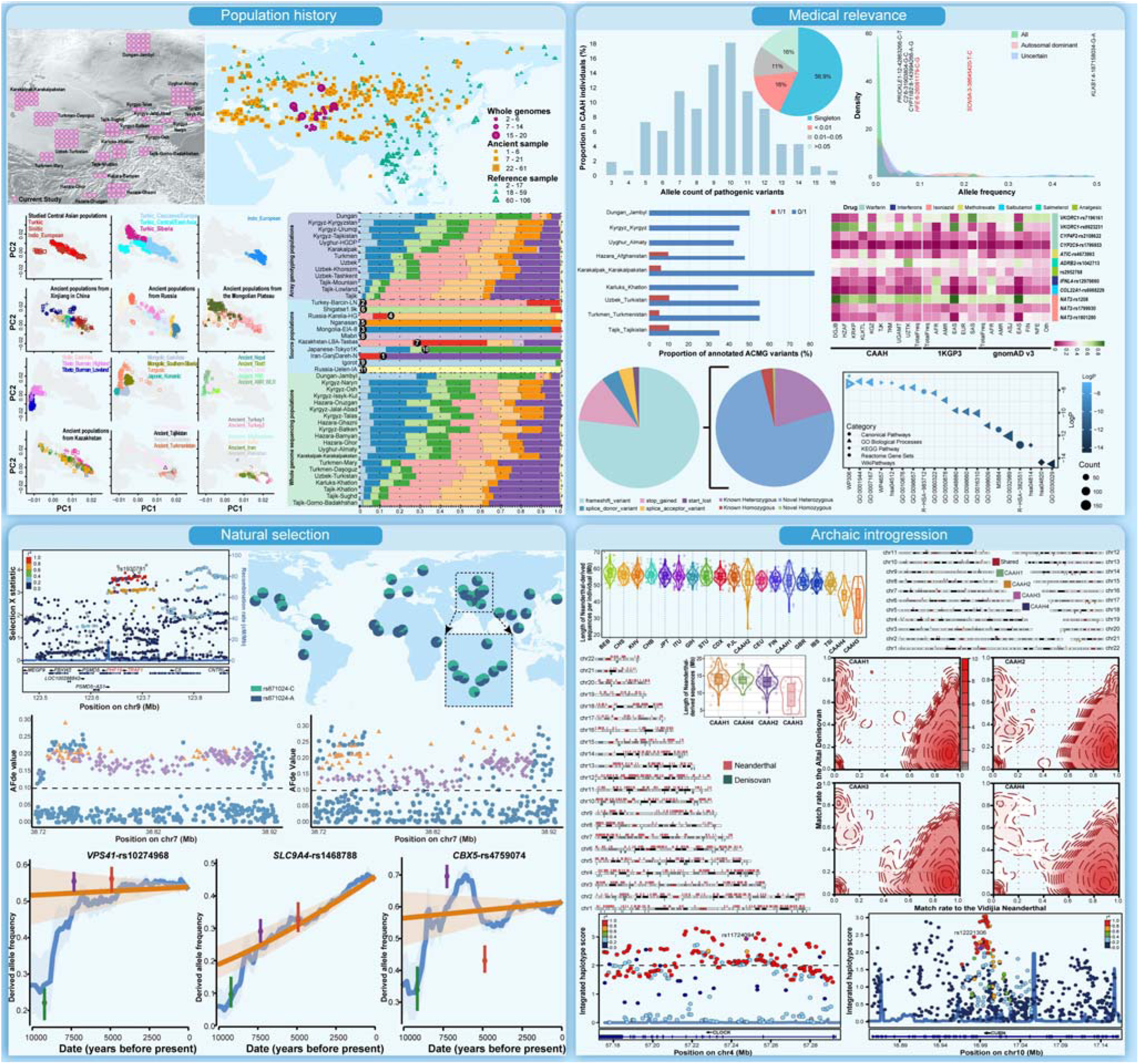

He *et al.* conducted a pilot study on the Central Asian Genomic Diversity Project, utilizing whole-genome sequencing of 166 individuals from 20 Central Asian populations. They identified fine-scale population substructures shaped by complex ancient trans-Eurasian migration and admixture processes. Their comprehensive analysis revealed post-admixture adaptations and archaic introgressions, shedding light on demographic events that influenced medically relevant mutation spectra and adaptations affecting immune, metabolic, neural, and physical traits. Neanderthal introgression segments significantly influence phenotypic traits, including susceptibility to immune and psychiatric disorders, whereas Denisovan-derived sequences have effects on disease susceptibility. This work advances our understanding of Central Asian genomic diversity and evolutionary history as well as their implications for health.

## Introduction

The Human Genome Project (HGP), initially aimed at determining the base pairs of the human genome and identifying, mapping, and sequencing all human genes, has significantly transformed research paradigms and led to numerous discoveries in genomic research^1^. These discoveries have elucidated aspects of demographic processes, adaptive histories, and genetic architectures related to human traits and diseases. Since the release of the human draft genome sequence, advances in sequencing technologies and computational methods, along with the publication of telomere-to-telomere-based and pangenome reference sequences, have facilitated comprehensive analyses of human genomic variations across ethnolinguistically diverse global populations^2–7^. Over the past two decades, there has been a substantial increase in the number of genomic studies; however, the majority of these studies have focused predominantly on individuals of European ancestry^6,8^. This Eurocentric bias not only fails to capture global genetic diversity but also impedes a thorough understanding of the molecular mechanisms underlying variable phenotypes and diseases, thereby exacerbating health inequalities^8–10^. Consequently, geneticists have underscored the importance of expanding large-scale genomic research to include non-European populations and establishing sustainable, diverse genomic initiatives worldwide^5,8–13^. Ongoing efforts to incorporate underrepresented non-European populations into human genomic research include initiatives such as the H3Africa (Human Heredity and Health in Africa) consortium^14^, the Simons Genome Diversity Project (SGDP)^15^, the GenomeAsia 100K Project^16^, the SG10K project^17^, and the PRECISE-SG100K population cohort undertaken by Singapore National Precision Medicine (NPM) workgroups^17,18^. Other significant contributions include the Oceanian genetic resource^19^, the Pakistan Alliance on genetic risk factors for health studies^8^, the BioBank Japan Project^20^, the China Kadoorie Biobank^21^, the Westlake BioBank for Chinese (WBBC) pilot project^22^, the NyuWa genome resource^23^, the China Metabolic Analytics Project (ChinaMAP)^24^, and the 10K Chinese People Genomic Diversity Project (10K_CPGDP)^4^. These genome initiatives are expanding our understanding of human genetic diversity and contributing to the elucidation of demographic history and disease etiology.

Central Asia, positioned as the Eurasian heartland, extends from Russia in the north to Iran and Afghanistan in the south and from the Caspian Sea in the west to China and Mongolia in the east. This region has historically served as a crossroads for the movement of people, languages, goods, and ideas between East and West Eurasia^25–27^. Despite their pivotal role in ancient trans-Eurasian cultural exchanges, Central Asian populations remain underrepresented in human genomic sequencing efforts and medical genome research^6^. Various demographic events have shaped the ethnolinguistic diversity of Central Asia. Anatomically modern humans are believed to have first occupied the region approximately 50 to 40 thousand years ago (kya). During the Paleolithic era, Central Asia was inhabited by a distinctive population known as “Ancient North Eurasian” (ANE), which contributed to the gene pool of Eurasia and the Americas through multiple migration events since the Upper Paleolithic^26,28^. To investigate the relationships among cultural practices, language distributions, and population movements, Narasimhan *et al*. analyzed whole-genome ancient DNA data from 523 individuals, primarily from Central Asia and northernmost South Asia. Their findings revealed that Neolithic individuals from Turan (present-day Turkmenistan, Uzbekistan, Tajikistan, Afghanistan, and Kyrgyzstan) and the central and southern Steppe (present-day Kazakhstan) presented ancestry related to West Siberian hunter-gatherers (WSHG), which were primarily derived from ANE and Eastern European hunter-gatherers (EEHG)^27^. This WSHG-related ancestry was prevalent in eastern Turan and Kazakhstan prior to the arrival of the Yamnaya steppe pastoralists.

Since the Bronze Age (BA), ancient Central Asians have experienced significant gene flow from Western Steppe herders (WSH) and Iranian/Anatolian farmers, which is correlated with the spread of Indo-European languages into Central Asia^27–29^. The “steppe hypothesis” posits that the expansions of groups related to the Yamnaya and Afanasievo cultures from the western steppe into Europe and Asia were facilitated by horse domestication. However, research by Damgaard *et al*. indicated that the Botai individuals in northern Kazakhstan, associated with early horse husbandry, derived most of their ancestry from an ANE-related hunter-gatherer population, which was distinct from the western steppe pastoralists^28,29^. The remaining ancestry was from ancient East Asian (AEA) populations represented by Baikal_EN. Formal modeling of BA Turan populations demonstrated that individuals from these sites predominantly had Iranian farmer-related ancestry, with additional contributions from Anatolian farmers and WSHGs and a minor proportion from Andamanese hunter-gatherers (AHG)^27^. Specifically, the Bactria Margiana Archaeological Complex (BMAC) population exhibited ancestry primarily from Iranian (∼60 to 65%) and Anatolian (∼20 to 25%) farmers, with supplementary contributions from WSHGs (∼10%) and AHGs (∼2 to 5%). Notably, steppe pastoralist-derived ancestry was absent in BMAC groups before 2100 BCE but appeared in outlier individuals from approximately 2100 to 1700 BCE^27^. During the Middle and Late Bronze Age (MLBA), communities in the Bactria region of Uzbekistan presented an increased proportion of steppe-related ancestry compared with early Bronze Age (EBA) populations, reflecting the growing influence of steppe pastoralists in Central Asia^27^. Since the Iron Age (IA), migrations from southern Siberia and East Asia, which are associated primarily with the expansion of proto-Mongolic and proto-Turkic speakers, have substantially contributed to the gene pool of Central Asians^26,30,31^. Although steppe-related ancestry increased from the BA to the IA, it did not entirely replace the previously identified Iranian and Anatolian farmer-related ancestry^27,31^. Analysis of 137 ancient genomes spanning 4,000 years revealed that IA Scythian nomads were highly structured and mixed with eastern steppe pastoralists, forming the Xiongnu confederations^30^. The Xiongnu nomads migrated westward in the second to third centuries BCE, eventually contributing to the formation of Huns in the fourth to fifth centuries AD. These migration and admixture events led to the gradual assimilation and replacement of Indo-European-speaking locals by Turkic groups, transforming Central Asia into a region predominantly characterized by East Asian ancestry.

The spread of farming and herding practices, alongside the historical expansions of Indo-European and Turkic speakers, has significantly influenced the genetic diversity of Central Asia^26,28–30^. From the fifth century AD, a series of centralized empires emerged in Central and East Asia, succeeding in the Xiongnu and Hunnic confederations^30,32^. These included the Turkic, Uyghur, Khitan, and Mongol empires. Genetic, archaeological, and historical evidence indicates that the western expansion of the Mongol Empire profoundly impacted the genetic landscape of present-day Central Asians^32,33^. The complex demographic processes in Central Asia have resulted in a composite genetic architecture, with the region predominantly inhabited by two culturally and linguistically diverse groups: the Turkic/Mongolic groups, including the Kyrgyz, Kazakh, Uyghur, Uzbek, Tatar, Karakalpak, Turkmen, Mongolian, and Kalmyk, and the Indo-European groups, including the Tajik, Russian, Hazara, Armenian, and Ukrainian. Additionally, some populations speak languages that are distinct from these main groups, such as Dungan, Korean, and Chechen. Early findings based on partial low-density genetic markers suggested that Central Asians exhibit the highest genetic diversity among Eurasian populations^34^. To further elucidate the demographic history of Central Asians, genome-wide studies utilizing single nucleotide polymorphisms (SNPs) have been increasingly employed. These studies identified recent admixture events and revealed that gene flow from European, West Asian, East Asian, and South Asian sources has significantly shaped the current genetic landscape of Central Asia^28,33,35,36^. For example, Yunusbayev *et al*. investigated 373 individuals from 22 Turkic groups across Eurasia and reported that these Turkic speakers possessed varying proportions of Asian ancestry originating from Mongolia and southern Siberia^33^. Perle *et al*. examined the admixture history of Yaghnobis and Tajiks and reported that present-day Indo-European-speaking populations in Central Asia have maintained genetic continuity since the Iron Age, with limited recent gene flow from other Eurasian groups^37^. A recent study reported that contemporary Tajik populations derived their ancestry primarily from BMAC and Andronovo-related groups, with highland Tajiks exhibiting additional ancestry related to the Tarim mummies. The study also revealed that the West Eurasian ancestry of the Kyrgyz people was traced primarily back to historical Xinjiang individuals^25^.

Despite these advancements, previous research has been limited by the use of partial genomic data from relatively small populations or sample sizes, leaving gaps in our understanding of the whole landscape of genomic diversity and genetic contributions of spatiotemporally diverse Eurasian populations to Central Asian genetic heritage and adaptive histories. Therefore, there is a pressing need for comprehensive genomic studies of underrepresented Central Asian populations to characterize their genetic diversity fully through large-scale whole-genome sequencing (WGS). The enhanced resolution provided by WGS enables the application of haplotype-sharing methods and integration with data from spatiotemporally distinct ancient individuals. This approach sheds light on modern human population dynamics and the genetic interactions between ancient and contemporary humans. To address the underrepresentation of Central Asian genomic resources, we conducted WGS on 166 individuals from nine ethnic groups (Kyrgyz, Uzbek, Uyghur, Turkmen, Karakalpak, Karluks, Tajik, Hazara, and Dungan) across Central Asia and Afghanistan in the pilot work of the Central Asian Genomic Diversity Project (CAGDP). This study aimed to fill gaps in the current genomic landscape. We extensively analyzed the demographic and adaptive history of ethnolinguistically diverse Central Asians by integrating publicly available WGS and genome-wide SNP data from both ancient and modern Eurasians (**Table S1**). Our findings offer insights into the evolutionary history and local adaptation of contemporary Central Asians and enhance our understanding of the genetic interactions between West and East Eurasians.

## Results

### Overview of population genomic variation

We sequenced the genomes of 166 individuals from 20 populations across Central Asia and Afghanistan via the CAGDP program, with an average depth of 13.66×. This analysis identified 22,994,949 SNPs and 2,952,662 small insertions or deletions (InDels) across 22 autosomes and the X chromosome. Among these, 2,300,814 SNPs (10.0%) and 834,296 InDels (28.3%) were novel and not reported in dbSNP version 156. Most of these unreported SNPs were extremely rare: 89.4% were singletons, 9.1% had a minor allele frequency (MAF) of less than 0.01, and 1.2% had an MAF between 0.01 and 0.05. Among the 22,538,640 biallelic SNPs observed, 40.2% were singletons, 15.1% had an MAF less than 0.01 (excluding singletons), and 14.7% had an MAF between 0.01 and 0.05. We calculated a transition versus transversion ratio (Ti/Tv) of 2.05 from 21,608,632 biallelic autosomal SNPs, which aligns with findings reported in previous studies^17^.

### Patterns of the population structure of Central Asian people in a Eurasian context

We conducted a principal component analysis (PCA) to examine the genetic affinity between newly sequenced Central Asian and Afghanistan Hazaras (CAAH) and Eurasian reference populations via the WGS dataset. The PCA revealed that CAAH were distinctly structured, positioning themselves between the West and East Eurasian clines. Specifically, Uzbek individuals aligned more closely with West Eurasians, whereas Dungan individuals aligned with East Eurasians (**Fig. S1**). To further explore the fine-scale population structure of CAAH, we integrated publicly available ancient and modern Eurasian population data from the Human Origins (HO) dataset. The first principal component (PC1) distinguished West Eurasians from East Eurasians, whereas the second principal component (PC2) differentiated West Eurasians along an east-west cline and East Eurasians along a north-south cline (**Figs. 1B and S2A**). The non-Dungan (CAAH_nDG) groups were distributed along the West Eurasian-related cline, whereas Dungan individuals were situated on the northern branch of the East Eurasian-related cline. Specifically, the Tajik and Karluks people presented close genetic relationships with Indo-European and Turkic-speaking populations in the Caucasus. Uzbek and Turkmen individuals were dispersed among Turkic groups in West Eurasia, whereas Kyrgyz, Uyghur, Karakalpak, and Hazara individuals overlapped with Turkic groups from Central Asia. The Dungan people, in contrast, showed genetic affinities with Mongolic, Tibeto-Burman, and Turkic speakers from China.

**Figure 1.**
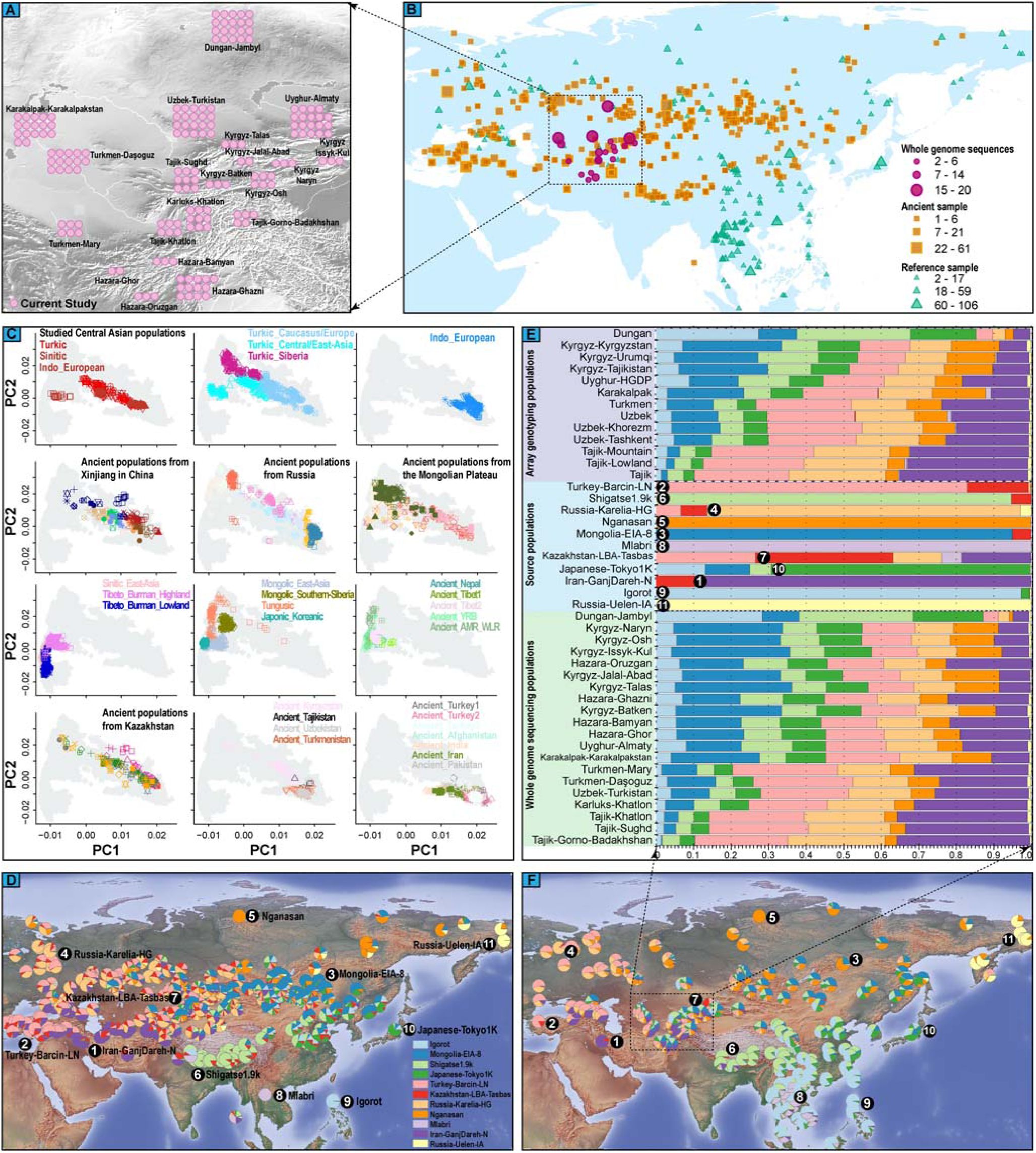
The geographical locations of newly sequenced Central Asians and Afghanistan Hazaras (CAAH) and linguistically close Eurasian reference populations and the population structure revealed based on the merged Human Origins (HO) dataset in the context of Eurasian populations. **(A),** Geographical locations of 20 newly collected Central Asian populations and Indo-European (Indo-European), Sino-Tibetan, and Altaic-speaking reference populations. The color-coded legends of geographical locations are consistent with those used in principal component analysis (PCA). **(B)**, The clustering patterns of 897 modern and ancient Eurasian populations revealed by PCA, in which ancient populations were projected onto the first two PCs. Modern populations are classified according to language family, and ancient populations are classified according to geographical area. The simplified legends of modern and ancient populations are presented, and the detailed population legends are presented in **Figure S2**. (C), Model-based ADMIXTURE results at K=11. Only the top five populations with the specific ancestral component are shown in the plot, and the average admixture coefficient values of each population are presented. The composition of the admixture coefficients for each newly studied Central Asian population is shown. The Kyrgyz people from Talas, Issyk-Kul, Naryn, Osh, Batken, and Jalal-Abad Regions were merged as Kyrgyz_Kyrgyzstan (KGZ); the Turkmen people from the Daşoguz and Mary Regions were merged as Turkmen_Turkmenistan (TKM); the Hazara people from Bamyan, Ghazni, Oruzgan, and Ghor Regions were merged as Hazara_Afghanistan (HZAF); and the Tajik people from the Sughd, Gorno-Badakhshan, and Khatlon Regions were merged as Tajik_Tajikistan (TJK).

In terms of ancient reference populations, CAAH_nDG either overlapped with or were closely related to individuals from IA Xinjiang and historical Kazakhstan. The Dungan people demonstrated a strong genetic connection to ancient populations from the Yellow River basin (YRB), the Qinghai□Xizang Plateau (QXP), and Nepal (**Figs. 1B and S2A**). Notably, Kyrgyz, Uyghur, Karakalpak, and Hazara individuals exhibited a shift toward ancient populations with less West Eurasian ancestry from the Mongolian Plateau (MP), suggesting that more genetic drift was shared with East Eurasian-related ancestry. In contrast, Tajik and some Uzbek/Turkman individuals were closer to populations from the Iranian/Anatolian farmer-related cluster, steppe populations from Russia, ancient individuals from Turan (excluding Kyrgyzstan), and MLBA populations from Kazakhstan, indicating greater allele sharing with West Eurasian-related ancestries, particularly Iranian farmers, Anatolian farmers, and WSH-related ancestries. Additionally, these groups displayed close affinities with ancient individuals from Pakistan (**Fig. 1B**). Tajik, Karluks, Uzbek, and Turkmen individuals also presented close genetic relationships with ancient individuals from Kyrgyzstan. Further analysis of genetic coordinates for CAAH, incorporating linguistically similar populations, revealed that CAAH was distributed between Indo-European speakers from West Eurasia and Mongolic-speaking populations from the MP (**Fig. S2B**). Indo-European-speaking populations appeared to be divided into two east-west genetic clines: one comprising individuals from regions west of the Caucasus and the other from the Caucasus region and Central Asia. Turkic-speaking populations were approximately divided into four genetic clusters corresponding to their geographic locations: the lower left cluster contained East Asians speaking Turkic languages; the upper left cluster comprised individuals from southern Siberia, extending toward Tungusic people; the central cluster included individuals from Central Asia; and the lower right cluster consisted of individuals from West Eurasia, extending toward Indo-European groups in West Eurasia (**Fig. S2B**).

The patterns of genetic affinity were consistent across unsupervised ADMIXTURE analyses (**Fig. S3**). The clustering patterns revealed by PCA and ADMIXTURE (**Figs. 1, S2, and S3**) revealed that geographically distinct but ethnically homogeneous populations, including Tajik, Turkmen, Hazara, and Kyrgyz, possessed similar ancestry compositions and were therefore merged into Tajik_Tajikistan (TJK), Turkmen_Turkmenistan (TKM), Hazara_Afghanistan (HZAF), and Kyrgyz_Kyrgyzstan (KGZ), respectively. In the K=11 model (**Figs. 1C and S3**), KGZ and Karakalpak_Karakalpakstan (KRKP) primarily derived their ancestry from Ulaanzuukh_SlabGrave (related to ancient Northeast Asian (ANA) populations), with additional contributions from Anatolian farmers (represented by Turkey_Barcin_LN), Japanese_Tokyo1K, Nganasan, Russia_Karelia_HG (related to EEHG), Iranian farmers (represented by Iran_GanjDareh_N), Shigatse1.9k/Lhasa1k (related to northern East Asian (NEA) populations), and Igorot/Atayal (related to southern East Asian (SEA) populations). Other CAAH also presented these eight ancestral components. Notably, Uyghur_Almaty (UGAMT) displayed similar proportions of Anatolian farmers, Iranian farmers, ANA, EEHG, and NEA-related ancestries, with minor contributions from Japanese_Tokyo1K, SEA, and Nganasan-related ancestries. HZAF had a comparable ancestry profile to UGAMT, with predominant Iranian farmer-related components. TJK, Karluks_Khatlon (KLKTL), Uzbek_Turkistan (UZTK), and TKM derived the majority of their ancestry from Anatolian and Iranian farmer-related lineages, supplemented by EEHG, ANA, Nganasan, Japanese_Tokyo1K, NEA, and SEA-related ancestries. Dungan, known for historical migration, had East Asian-derived ancestries (NEA, SEA, and Japanese_Tokyo1K), with additional contributions from ANA, West Eurasian (Anatolian farmer, Iranian farmer, and EEHG), and Nganasan-related ancestries. Outgroup *f_3_*-statistics revealed that KGZ, UGAMT, KRKP, and HZAF shared more alleles with Altaic-speaking populations in northern East Asia and southern Siberia, as well as ancient populations from the ARB, West Liao River (WLR), and YRB; UZTK, KLKTL, TKM, and TJK presented greater allele sharing with modern Siberian populations, early and middle Bronze Age (EMBA) Tarim individuals, and ancient populations with substantial steppe-related ancestry from Kazakhstan and southern Siberia; The Dungan_Jambyl (DGJB) population presented greater allele sharing with East Asian (especially Sinitic-speaking) and YRB ancient populations (**Figs. S4 and S5**), highlighting distinct admixture processes. The hierarchical phylogenetic structure inferred from TreeMix corroborated these differential allele-sharing patterns observed via PCA, ADMIXTURE, and outgroup *f_3_*-statistics (**Fig. S6**).

**Figure 2.**
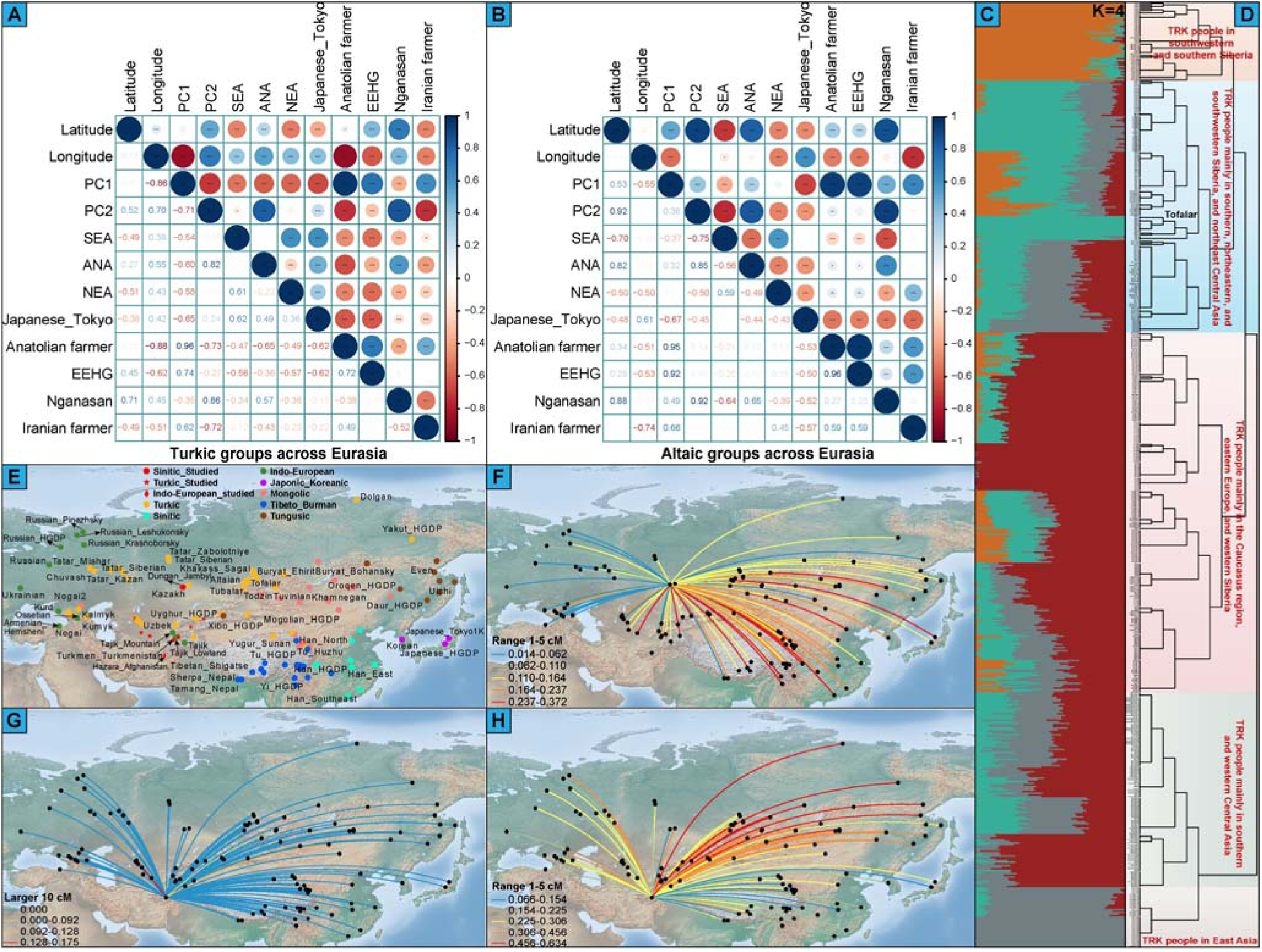
The correlation between geographical and genetic patterns, the fine-scale population structure of 47 Turkic groups, and identity-by-descent (IBD) sharing between Dungan_Jambyl (DGJB)/HZAF and Eurasian reference populations. The correlation between geographical (latitude and longitude) and genetic (PC1, PC2, and estimated ancestral components in Fig. 1E) features and between estimated ancestral components among Turkic groups (**A**) and among Altaic groups (**B**). SEA: southern East Asian ancestry; ANA: ancient Northeast Asian ancestry; NEA: northern East Asian ancestry; EEHG: Eastern European hunter-gatherers. (**C**), Model-based ADMIXTURE results for Turkic-speaking populations at K=4. (**D**), The fineSTRUCTURE-based topology revealed the fine-scale population substructures among the Turkic groups. (**E**), Geographical distribution of reference populations involved in the IBD analysis. Reference populations are assigned different colors according to their linguistic affiliation. (**F**), The summed length of identified IBD blocks in the range of 1–5 cM between the DGJB and Eurasian reference populations. (**G**□**H**), The summed length of identified IBD blocks over 10 cM and in the range of 1–5 cM between the HZAF and Eurasian reference populations.

### Geography-related genetic differentiation among 47 Turkic groups across Eurasia

Turkic people are widely distributed around Central Asia and surrounding regions, and overall patterns of genetic structure remain characterized. The PCA and ADMIXTURE analyses revealed significant genetic differences among 47 geographically distinct Turkic-speaking populations across Eurasia and identified several Turkic-related clusters (**Figs. 1, S2, and S3**). The TreeMix-based topology further illustrated the distinct population structures of Turkic-speaking groups, distinguishing between those primarily from Central Asia, East Asia, and Siberia and those from the Caucasus and Europe (**Fig. S7**). To understand the factors influencing genetic differences among geographically diverse Turkic-speaking populations, we examined the correlations between geographic variables (latitude and longitude) and genetic features (PC coefficients and admixture proportions of ancestral populations). PC1 was negatively correlated with longitude, whereas PC2 was positively correlated with both longitude and latitude. These findings indicated genetic distinctions between the eastern and western Eurasian Turkic groups as well as between the northern and southern Eurasian Turkic groups (**Fig. 2A**). The ancestral components related to EEHG, ANA, and Nganasan were positively correlated with latitude, whereas East Asian and Iranian farmer-related ancestral components were negatively correlated with latitude. The ancestral components associated with East Asians, ANA, and Nganasan were positively correlated with longitude, whereas Anatolian farmers, Iranian farmers, and EEHG-related components presented negative correlations with longitude (**Fig. 2A**). In contrast, the correlation between geographic distance and genetic differentiation in the Altaic-speaking populations differed from that observed in the Turkic groups (**Fig. 2B**). Notably, among Altaic speakers, the proportion of Anatolian farmer-related ancestry was positively correlated with latitude, and the proportion of Iranian farmer-related ancestry was not significantly correlated with latitude.

We then performed admixture modeling on all the included Turkic groups to reconstruct the fine-scale population structure and identified the best-fitting model using four predefined ancestral sources. These sources were maximized in Karachai, Balkar, and Kumyk in the Caucasus region; Altaian_Chelkans and Tubalar in southwestern Siberia; Tofalar in southern Siberia; and Salar_Xunhua in East Asia (**Fig. 2C**). To further explore the fine-scale genetic structure of geographically distinct Turkic groups on the basis of shared haplotypes, we phased the genomes of 589 Turkic individuals and conducted fineSTRUCTURE and identity-by-descent (IBD) analyses (**Figs. 2D and S8**). The resulting dendrogram revealed that while there was continuity in haplotype-based structure, Turkic-speaking populations across Eurasia were also divided into five clusters corresponding to geographic regions (**Fig. 2D**). The IBD analysis estimated significant recent population interactions (within the last 500 years) among Turkic groups in southwestern Siberia, southern Siberia, and northeastern Siberia (**Fig. S8B**). We identified shared IBD segments ranging from 5 to 10 cM, indicating extensive gene flow between 1500 and 500 years ago among these groups, as well as between Turkic groups in southern and southwestern Siberia (**Fig. S8C**). Additionally, we found evidence of ancient genetic connections between the Turkic people in eastern and western Eurasia (**Fig. S8D**).

### Long-distance migration of Dungan and Hazara people shaped their unique genetic makeup

The patterns of genetic relationships among geographically distinct populations mostly follow an isolation-by-distance model. However, the strong genetic affinities observed among geographically disparate Eurasian populations provide direct evidence for long-range migrations. Clustering patterns revealed by PCA, ADMIXTURE, and outgroup *f_3_*-statistics revealed that the DGJB and HZAF populations did not exhibit strong genetic relatedness to geographically or linguistically close Central Asian populations. Instead, they demonstrated closer genetic ties with East Asian populations (**Figs. 1B and S1**□**S4**). This finding was further supported by pairwise Fst genetic distances (**Fig. S9**). Specifically, DGJB shared more alleles with East Asians, particularly Sino-Tibetan-speaking populations, than with geographically proximate Central Asians. HZAF displayed admixture profiles similar to those of Uyghur populations and showed additional affinity for Tungusic groups from northern East Asia and Siberia. We observed that the proportions of inferred ancestries, except for SEA and NEA, in DGJB-related Sino-Tibetan groups correlated positively with latitude. In contrast, the proportions of inferred ancestries other than SEA and Japanese_Tokyo1K correlated negatively with longitude (**Fig. S10A**). In Hazara-related Indo-European groups, the proportions of ancestral components associated with East Asian, ANA, and Iranian farmers were negatively correlated with latitude, whereas these proportions were inversely correlated with longitude (**Fig. S10B**). To further explore genetic connections between DGJB/HZAF and geographically diverse reference populations on the basis of shared haplotypes, we estimated IBD sharing between different population pairs. We then visualized genetic relatedness across geographically diverse populations over various time scales. Unexpectedly, we identified extensive ancient connections between the DGJB and East Asian populations, particularly with the Sino-Tibetan groups (**Figs. 2E, F, and S10C**). Although the HZAF population exhibited substantial IBD sharing with geographically and linguistically distinct populations, such as Turkmen in Uzbekistan, Mongol minorities in Mongolia, and Tuvinian in southern Siberia, the HZAF population presented extensive ancient connections, primarily with Altaic groups in Siberia (**Figs. 2G, H, and S10D**). Overall, the patterns of allele and haplotype sharing between the DGJB and Sino-Tibetan groups, as well as between HZAF and northern East Asian and Siberian populations, along with the predominantly East Asian ancestry in DGJB and the significant East Eurasian ancestry in HZAF, suggested that long-distance migrations of DGJB- or HZAF-related ancestral populations have significantly shaped their genetic profiles.

### Admixture modeling and admixture time estimation of CAAH

We first applied admixture *f_3_*-statistics to characterize the gene pools of the CAAH group. Reference pairs for CAAH, excluding KLKTL and DGJB, with the most negative *f_3_* values, primarily involved one eastern and one western Eurasian source population. This pattern provides clear evidence that CAAH (except KLKTL and DGJB) is a mixture of groups with deep connections to both eastern and western Eurasian sources. Different patterns of admixture signatures for the other two groups highlight the complex admixture landscape of KLKTL and DGJB (**Tables S2**□**S3**). Our analysis revealed that reference pairs involving Nanai and Ulchi produced more significantly negative *f_3_* values in KGZ than did those involving other East Eurasian sources. For UGAMT, UZTK, and HZAF, reference pairs with Han Chinese, Japanese, Nanai, and Ulchi primarily yielded more negative *f_3_* values. Additionally, UGAMT and UZTK presented more negative *f_3_* values when ANA-related ancestries were used as East Eurasian sources. TKM and KRKP presented more negative *f_3_* values with Nanai, Ulchi, Nivkh, and ANA-related ancestries as East Eurasian sources (**Table S2**). Notably, reference pairs involving Anatolian/Iranian farmer-related ancestries generated more negative *f_3_* scores in TJK than those involving other West Eurasian sources did (**Table S3**). To further investigate genetic heterogeneity among geographically distinct CAAH groups, we performed *f_4_*-statistics of the form *f_4_* (CAAH_1_, CAAH_2_; Reference, Mbuti). The observed patterns of population stratification in the descriptive results were consistent with the findings from the symmetric *f_4_*analysis. This analysis revealed significant estimates with absolute Z scores greater than 3, indicating that genetically distinct Central Asian populations presented varying patterns of allele sharing with the western or eastern reference populations (**Figs. 3A and S11**). Compared with other CAAH groups, DGJB presented the greatest genetic drift with AEA-related and ANA-related ancestry sources as well as ancient Central Asians with substantial East Asian ancestry (ACAs_EA)^30^, followed by KGZ, KRKP, and HZAF/UGAMT, which also shared more alleles with AEAs, ANAs, and ACAs_EA. Compared with TJK and TKM, UZTK demonstrated greater genetic affinity for AEAs and ACAs_EA and more genetic drift with ANAs and ancient northern East Asians (ANEAs) than KLKTL did. Furthermore, TKM and KLKTL presented greater allele sharing with AEAs, ANAs, and ACAs_EA than did TJK. The patterns of allele sharing between CAAH and Anatolian farmers, Iranian farmers, and steppe pastoralist-related ancestors contrasted with those observed between CAAH and AEAs, ANAs, and ACAs_EA (**Figs. 3A and S11**).

**Figure 3.**
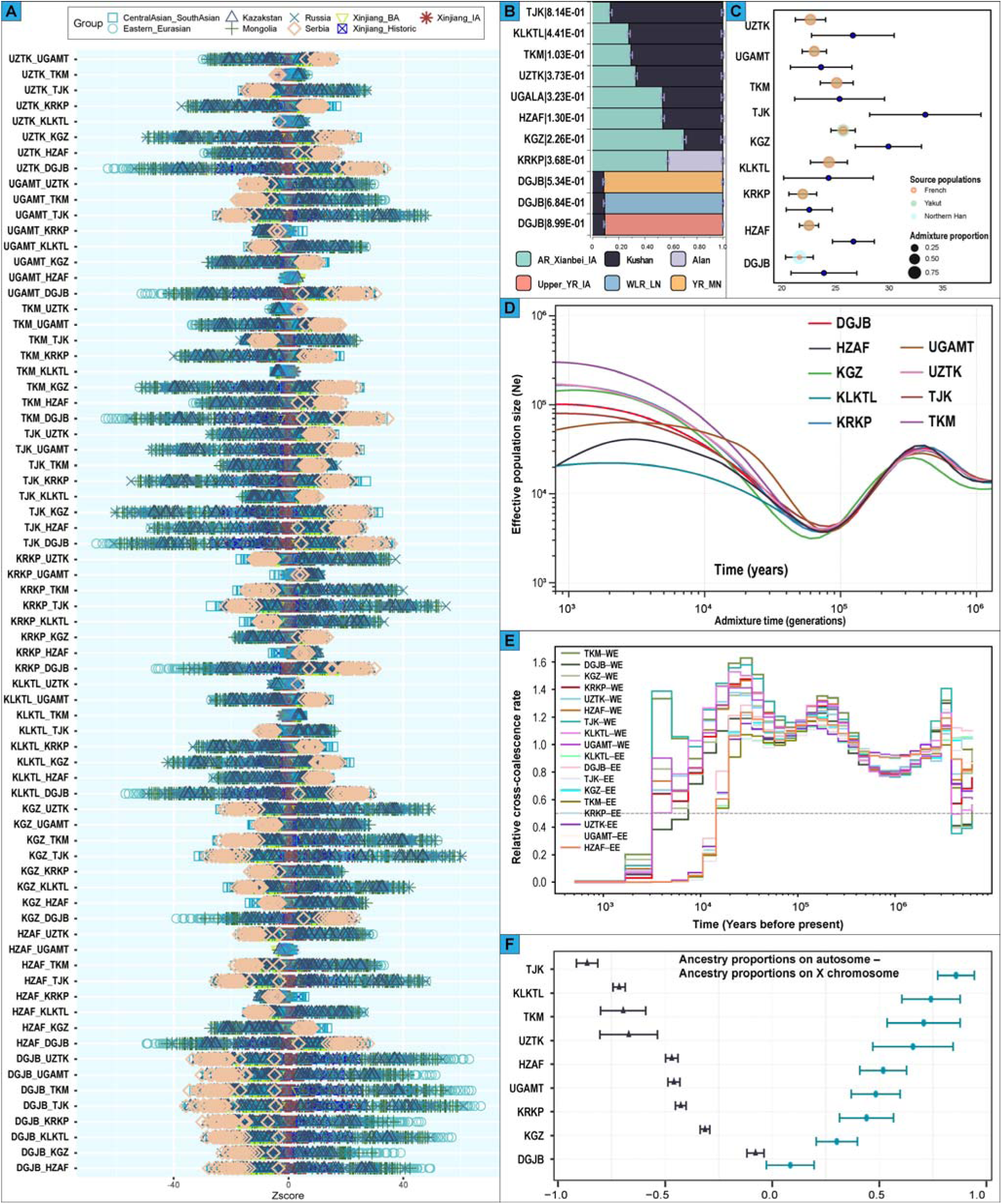
Admixture profiles and sex-biased patterns of CAAH. (**A**), The values of *the f*_4_-statistics of the form *f*_4_(CAAH_1_, CAAH_2_; Reference, Mbuti). Ancient Eurasian groups were used as reference populations. Different colored shapes represent different groups of ancient reference populations. UZTK: Uzbek_Turkistan, UGAMT: Uyghur_Almaty, KRKP: Karakalpak_Karakalpakstan, and KLKTL: Karluks_Khatlon. (**B**), Admixture proportions of CAAH based on qpAdm estimation. (**C**), The admixture proportions were inferred from fastGLOBETROTTER analysis, and admixture times were inferred from fastGLOBETROTTER and MALDER analyses. The blue circles denote the MALDER-based admixture times. Other colored circles denote the fastGLOBETROTTER-based admixture times and ancestry compositions. The color of the circle represents the different surrogate ancestral sources, and the size of the circle reflects the admixture proportion. Admixture times are presented as the mean ± standard deviation. (**D**), Effective population sizes of CAAH. (**E**), The population divergence times between CAAH and East/West Eurasian source populations. EE: East Eurasian population, WE: West Eurasian population. (**F**), Sex-biased patterns of genetic admixture in CAAH are estimated based on autosomal and X-chromosomal genetic variations.

To further model the ancestry profiles of each group, we used qpAdm to evaluate whether a tested group was derived from a candidate set of source populations and, if so, to estimate the corresponding ancestry coefficients. A two-way mixture of Tajikistan_Ksirov_Kushan (30%–86%), a historic nomadic group descended from the BMAC, Hajji_Firuz_C, and Steppe_LBA, and China_AR_Xianbei_IA (14%–70%) fit well for CAAH, except for DGJB and KRKP (**Fig. 3B**). DGJB was best modeled, with only 8.6%–9.3% Tajikistan_Ksirov_Kushan ancestry, requiring 90.7%–91.4% ancestry from ANEAs (China_Upper_YR_IA, China_WLR_LN, or China_YR_MN). KRKP can be modeled as a simple two-way mixture of western and eastern Eurasian steppe pastoralists, with approximately equal proportions of Russia_Alan (42%), a historic nomadic group likely descended from the Sarmatians and contemporary Huns, and China_AR_Xianbei_IA (58%), which represents a mixture of Nganasan, Ami (SEA-related), and Sherpa (NEA-related). We also used haplotype-based SOURCEFIND and fastGLOBETROTTER to characterize ancestry patterns in CAAH based on the WGS dataset. The SOURCEFIND estimates suggested that French, Kalash, Bedouin, Yakut, and Han_Northern, identified in the ADMIXTURE modeling, could serve as potential source surrogates and differentially contributed haplotype fragments to CAAH (**Fig. S12**). Running fastGLOBETROTTER on these proxies indicated that a one-date admixture event fit well for CAAH, except for TJK, where multiple admixture dates were detected (**Figs. 3C and S13**). West Eurasian ancestry, represented by French, contributed between 32% and 45% to KGZ, KRKP, HZAF, and UGAMT but accounted for more than 50% of the ancestry in TKM, UZTK, and KLKTL (57%-62%). The remaining ancestry in these populations was derived from the Yakut people, representing East Eurasian ancestry. DGJB could be modeled as a mixture of 87% Han_Northern and 13% French. The “best-guess” scenario identified two admixture events in TJK, which could be modeled as a mixture of surrogate source populations composed of French, Bedouin, Kalash, Han_Northern, and Yakut in varying proportions (**Fig. S13B**).

We used MALDER and fastGLOBETROTTER to date the inferred east-west admixture events. The linkage disequilibrium (LD)-based MALDER identified a simple admixture event between a West Eurasian source (French) and an East Eurasian source (Yakut or Han_Northern) in CAAH, which occurred approximately 23-34 generations ago (**Fig. 3C**). FastGLOBETROTTER revealed similar admixture dates of approximately 22-25 generations in TKM, KLKTL, UGAMT, and KRKP and relatively recent admixture dates of approximately 22-26 generations in UZTK, HZAF, KGZ, and DGJB (**Fig. 3C**). We simulated two admixture events in TJK: the most recent event occurred approximately nine generations ago, and an older event occurred approximately 50 generations ago. We also applied SMC++ to infer the effective population size (Ne) and found that the Ne of KLKTL was significantly lower than that of the other CAAH populations. Most CAAH populations showed substantial growth over the past 10,000 years; however, the Ne values of KLKTL and UGAMT/HZAF peaked approximately 2,000 and 3,000 years ago, respectively, before declining (**Fig. 3D**). Population divergence time estimates revealed splits between East Eurasians (represented by Northern Han Chinese) and CAAH at 20.8□15.3 kya, between West Eurasians (represented by French) and CAAH at 2.6-7.2 kya, and among CAAH groups at 1.9□3.3 kya (**Figs. 3E and S14A**). Additionally, the proportion of West Eurasian ancestry in CAAH was directly proportional to the divergence times between these groups and East Eurasians and inversely proportional to the divergence times between them and West Eurasians (**Fig. S14B and C**).

### Sex-biased admixture landscape

To investigate sex-biased admixture processes in CAAH, we estimated and compared the admixture proportions of source populations based on variations in autosomal, X-chromosomal, Y-chromosomal, and mtDNA. The difference in ancestry proportions between autosomes and X chromosomes suggests sex-biased admixture, as males inherit only their mother’s X chromosome. The western Eurasian genetic contribution to CAAH was estimated to be 7.8−86.4% for autosomes and 8.4−85.8% for X chromosomes (**Fig. 3F**). A clear bias toward female West Eurasian ancestry was observed in HZAF (Z = −0.43; Student’s t-test, P < 0.05), corresponding to an increase (71.4%) in West Eurasian-associated maternal lineages, such as haplogroups H, J, K, N1, T, U, and W (**Table S4**). Although no significant sex bias was detected in other CAAH populations (−0.20 < Z < 0.13; Student’s t-test, P > 0.05), some populations presented differing proportions of West Eurasian-related paternal or maternal lineages compared with autosome- and X chromosome-based ancestry proportions (**Fig. 3F, Table S4**). All KLKTL individuals belonged to the Y-chromosome haplogroup L1a2a, which is commonly found in South Asian populations. West Eurasian-related paternal lineages were present in 65.0% of KGZ, 35.3% of KRKP, and 30.0% of DGJB, whereas West Eurasian-derived maternal lineages were found in 88.9% of KLKTL, 70.0% of TJK, 55.0% of UZTK, and 31.6% of UGAMT (**Table S4**). Overall, there was no evidence of a simultaneous excess of West Eurasian-related paternal and East Eurasian-related maternal lineages or vice versa in Central Asians.

### Medically relevant and pharmacogenomic variants

We annotated the variants in our dataset against ClinVar and identified 116 pathogenic (level 5) variants across 99 genes. Each participant carried at least two pathogenic variants listed in the ClinVar database, with a median of 9 alleles per individual (range3-16) (**Fig. 4A**). Among the unique pathogenic variants, approximately 56.9% (66 out of 116) were singletons, approximately 16.4% had a MAF less than 0.01, and 13 variants (∼11.2%) had a MAF ranging from 0.01 to 0.05 (**Fig. 4A**). Among the 208 variants annotated as pathogenic or likely pathogenic (level 4), approximately 17.8% (37 out of 208) had a MAF > 0.05 in CAAH, with three having an average MAF < 0.05 and only one having a MAF < 0.05 across all population groups in the Genome Aggregation Database (gnomAD v3.1.2) (**Fig. 4B, Table S5**). Furthermore, a majority (∼51.9%) of these variants were linked to autosomal recessive (AR) disorders, whereas around 20.7% were associated with autosomal dominant (AD) disorders (**Figs. 4B and S15A**). We then reviewed the identified pathogenic or likely pathogenic variants with the American College of Medical Genetics and Genomics (ACMG) Secondary Finding gene panel (ACMG-SF v3.2) to evaluate the potential clinical utility of our genomic data and provide a more comprehensive understanding of medically relevant genetic variants^38^. Surprisingly, 92 out of 166 individuals carried at least one of 10 reportable ACMG variants across seven genes (**Fig. 4C**). Four of these variants were restricted to singletons in *MUTYH* and *BTD*, linked to AR disorders, and LDLR, associated with AD disorders. Additionally, two variants were identified as doubletons in *BRCA2*, associated with AD disorders, and *ATP7B*, linked to AR disorders. Notably, 65 individuals (∼39.2%) carried the *SCN5A*-A1673G (rs1805124, H558R) variant (**Fig. 4B**), which is associated with autosomal dominant ventricular fibrillation and influences the cardiac sodium channel alpha subunit, a crucial element for the electrophysiological function of the heart. Polymorphic variants of the *SCN5A* gene have also been linked to several heart conditions, including Brugada syndrome, progressive heart conduction defects, sick sinus syndrome, atrial fibrillation, and dilated cardiomyopathy. This variant has been identified across various ethnic groups at different frequencies, with the highest prevalence in KRKP (∼41.2%), followed by African populations (**Fig. S15B**). These findings suggest a complex interplay between genetic and environmental factors or the potential misclassification of the variant in the ACMG and other genetic databases.

**Figure 4.**
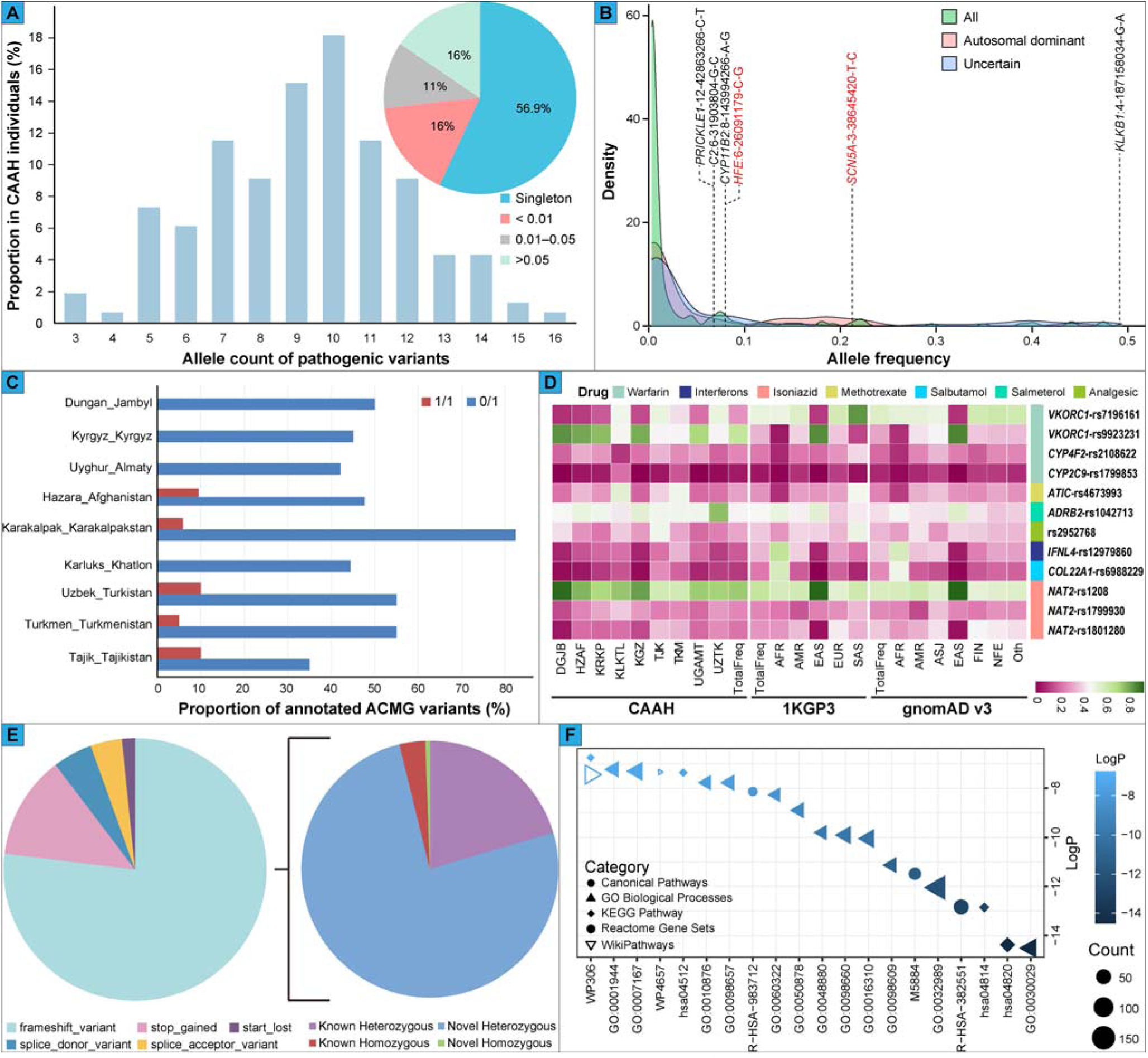
Statistics of medically relevant genetic variants in our dataset. (**A**), The number of alleles of identified pathogenic variants carried by each CAAH individual, alongside the allele frequency (AF) distribution of these variants. (**B**), Density plot displaying the frequencies of pathogenic and likely pathogenic ClinVar variants (n = 208), categorized by the most common inheritance patterns of associated monogenic disease genes; variants with autosomal dominant inheritance or unknown modes are highlighted. The variant with the highest minor allele frequency (MAF) value, three variants with MAFs > 0.05 in CAAH but average MAFs < 0.05 in the Genome Aggregation Database (gnomAD v3.1.2), and two reportable American College of Medical Genetics and Genomics (ACMG) variants with MAFs > 0.05 are presented as gene names: chromosome-base pair position-reference allele-alternative allele. (**C**), Distribution of homozygous (1/1) and heterozygous (0/1) genotypes for annotated ACMG variants across each CAAH population. (**D**), Frequency spectra of derived alleles for pharmacogenomic variants in our dataset, the 1000 Genomes Project, and gnomAD. AFR: African, AMR: American, EAS: East Asian, EUR: European, SAS: South Asian, ASJ: Ashkenazi Jewish, FIN: Finnish, NFE: non-Finnish European, and Oth: Remaining individuals. (**E**), Functional annotations of putative loss-of-function variants and the proportions of novel and known heterozygous and homozygous protein-truncating variants (PTVs) in our dataset. (**F**), The enrichment results of annotated heterozygous PTVs.

We examined the allele frequencies (AFs) of known pharmacogenomic variants in the ADME gene list, as well as variants reported by ClinVar to influence drug response. Significant differences in the AFs of some pharmacogenomic variants were found among CAAH and intercontinental populations (**Fig. 4D**). For example, warfarin, a commonly prescribed oral anticoagulant, reduces blood clotting by inhibiting vitamin K epoxide reductase. Individuals with the *VKORC1*-1639AA (rs9923231) homozygous genotype needed a lower dose than those with the heterozygous genotype did. We observed high derived allele frequencies (DAFs) of rs9923231 (≥ 73.7%) in DGJB, HZAF, KRKP, KGZ, and UGAMT, indicating relatively high East Eurasian ancestry. The DAFs of rs7196161 in *VKORC1* (≥ 42.5%) in the KLKTL, TJK, TKM, UZTK, and non-East Asian reference populations (particularly South Asians) were greater than those in the other CAAH groups. Individuals with the AA genotype (mutant-type) of rs7196161 may require an increased warfarin dose. Additionally, significant AF differences were observed in *CYP4F2*3* (rs2108622) among CAAH, with individuals carrying the T allele (especially homozygous TT) requiring a higher dose of warfarin. The AFs of *CYP2C9*2* (rs1799853, C430T) detected in KLKTL and TKM were greater than those detected in other CAAH populations. The *CYP2C9*2* allele is linked to poor warfarin metabolism, necessitating a reduced dose of warfarin, and is also associated with increased sensitivity to the antiepileptic drug phenytoin and a greater risk of gastrointestinal bleeding with some non-steroidal anti-inflammatory drugs. Therefore, genotyping for *VKORC1*, *CYP2C9*, and *CYP4F2*, along with genotype-guided warfarin administration algorithms, should be employed in genetically diverse Central Asian populations. This approach is recommended before prescribing warfarin to ensure optimal dosing and minimize adverse effects. The AFs of *ATIC* T675C (rs4673993) were notably different among CAAH, with high frequencies (≥ 40%) in KLKTL, TJK, TKM, CDX, and reference South Asians (**Figs. 4D and S15C**). Individuals with the rs4673993 CC genotype may exhibit an enhanced response to methotrexate, indicating that *ATIC* genotyping should be conducted in Central Asians. Additionally, an extremely high DAF of rs1042713 in *ADRB2* was observed in UZTK (77.5%), suggesting a decreased response to salmeterol among UZTK, particularly in individuals with the rs1042713 AA genotype. We also found substantial AF differences in CAAH for variants associated with the response to analgesics (rs2952768), isoniazid [*NAT2*5* (rs1801280), *NAT2*6* (rs1799930), and *NAT2*12* (rs1208)], interferons (rs12979860), and salbutamol (rs6988229).

### Loss-of-function variants and human knockouts

Reconstructing the pattern of human homozygous loss-of-function (LoF) alleles provides valuable insights into potential strategies for disease treatment. Large-scale genomic studies have shown that genetically diverse populations possess numerous LoF variants. Identifying these homozygous LoF alleles allows researchers to assess the phenotypic consequences of gene loss. In our study, we identified 3,436 putative LoF (pLoF) variants, with 76.9% classified as “frameshift_variant,” 12.7% as “stop_gained,” 4.9% as “splice_donor_variant,” 3.9% as “splice_acceptor_variant,” and the remainder as “stop_lost” (**Fig. 4E**). We further analyzed 3,381 high-confidence protein-truncating variants (PTVs), most of which were heterozygous (96.2%) and unique to our dataset (78.8% of the heterozygous PTVs) (**Fig. 4E**). These heterozygous PTVs are located in genes involved in various biological pathways, including system development, cell motility, and signaling molecules and interactions (**Fig. 4F**). Notably, these heterozygous PTVs and their associated pathways were primarily associated with UGAMT, particularly those related to cytoskeletal function in muscle cells, head development, sensory system development, and 22q11.2 copy number variation syndrome (**Fig. S15D**). We also identified 129 homozygous PTVs across 118 genes in our dataset, with only eight not previously reported in dbSNP156. The novel homozygous PTVs included the *CCDC88C* allele in CAAH, the *NOL9* allele and two *CRK* alleles exclusive to UZTK, the *ZMIZ2* allele unique to TJK, the *RELN* allele specific to HZAF, and the *BPIFB3* and *TGM2* alleles found only in KGZ. A genome-wide association study (GWAS) of 19,629 individuals of European ancestry revealed that variants in *RELN* influence regional brain volumes and are associated with cognitive and mental health traits^39^.

### Genomic signatures of positive selection

Several studies have demonstrated that admixed populations have undergone rapid evolution following admixture^40,41^. As populations with significant admixture from East and West Eurasian sources, the AFs in CAAH are generally expected to represent averages of the ancestral populations, weighted by their respective admixture proportions under the admixture rule. However, local adaptation can lead to specific genetic variants in admixed populations with frequencies that diverge from these expectations^41,42^. To identify genomic signatures of positive selection in CAAH and investigate their biological significance, AF deviations from expected values (AFd_e_) and the proportion of variants exhibiting significant AFd_e_ in each gene region were calculated. Given the structural nature of CAAH, we divided it into four groups (CAAH1, CAAH2, CAAH3, and CAAH4) on the basis of fineSTRUCTURE clustering patterns. Genes with a high proportion of variants showing significant AFd_e_ were identified in pathways such as the “metabolic process”, “developmental process”, and “response to stimulus” across CAAH groups, whereas genes in the “immune system” pathway were specifically identified in CAAH1, CAAH2, and CAAH4 (**Fig. S16A**). Notably, variants exhibiting significant AF deviations within the top 10 gene regions across each CAAH population were not associated with notable phenotypic effects (**Fig. 5A–D**).

**Figure 5.**
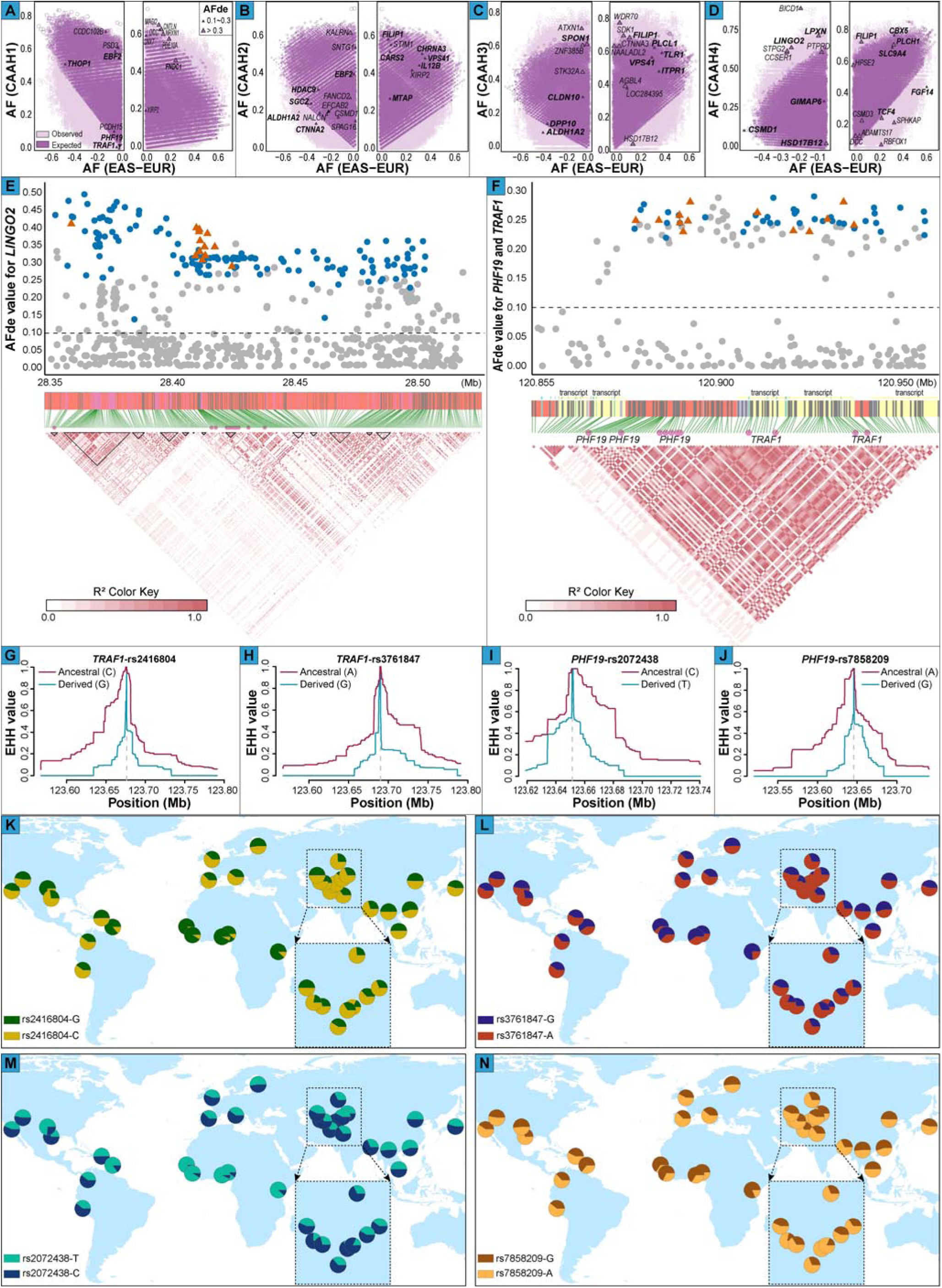
The signatures of biological adaptation identified in genetically diverse CAAH groups and the frequency distribution patterns of specific variants. (**A–D**), Distribution of observed and expected AFs for variants within each CAAH in the context of AF differences between East and West Eurasian populations. The top 10 genes per population, previously unreported in the GWAS Catalog for significant phenotypic effects, are shown in black, whereas well-studied genes are highlighted in red. Linkage patterns and deviations between observed and expected allele frequencies (AFd_e_) of variants in the *LINGO2* (**E**) and *TRAF1*/*PHF19* genes (**F**). The colored markers indicate natural selection signatures on these genes. The blue dots represent variants not previously associated with phenotypic effects in the GWAS Catalog, whereas the orange triangles denote variants that have been reported to exhibit phenotypic effects in the GWAS Catalog. Extended haplotype homozygosity (EHH) curves for *TRAF1*-rs2416804 (**G**), *TRAF1*-rs3761847 (**H**), *PHF19*-rs2072438 (**I**), and *PHF19*-rs7858209 (**J**). AF distributions of *TRAF1*-rs2416804 (**K**), *TRAF1*-rs3761847 (**L**), *PHF19*-rs2072438 (**M**), and *PHF19*-rs7858209 (**N**).

An AF-biased component was found within the *EBF2* region in both the CAAH1 and CAAH2 populations (**Fig. 5A and B**), which plays a critical role in the adipogenic transcriptional cascade^43^. Genetic variants in *EBF2* exhibiting significant AFd_e_ were associated with inguinal hernia^44^. The observed allele frequencies (AF_obs_) of the 3’ UTR variants, rs13257257-G (0.42 vs. 0.18 in CAAH1; 0.39 vs. 0.17 in CAAH2) and rs17054430-C (0.39 vs. 0.17 in CAAH1; 0.34 vs. 0.16 in CAAH2), exceeded the expected allele frequencies (AF_exp_) post-admixture, and rs3829011-A in CAAH2 showed a similar AF deviation (0.38 vs. 0.17). These deviations in *EBF2* variants may impair brown fat formation, potentially promoting white fat synthesis and contributing to increased fat storage and intra-abdominal pressure. Multiple functional loci within *FNDC1*, associated with cellular responses to hypoxia and the positive regulation of cardiomyocyte apoptosis, displayed AF bias in CAAH1 (**Fig. 5A**). Notably, the T allele of rs294883 in *FNDC1*, previously reported to be associated with coronary artery disease^45^, was observed at a higher frequency (0.44) than expected post-admixture (0.15). Genetic variations in *CARS2* influence the synthesis of enzymes involved in serine biosynthesis, thereby affecting protein synthesis (**Fig. 5B**). The variants rs7324648 and rs373373, located within *CARS2* and identified in CAAH2, have been reported to be associated with levels of aspartate aminotransferase^46,47^. The AF_obs_ of rs7324648-A (0.22 vs. 0.04) and rs373373-C (0.21 vs. 0.06) were significantly higher than the expected post-admixture frequencies. The *CLDN10* gene, which plays a crucial role in kidney function and intestinal barrier integrity, contains several functional loci with AF deviations in CAAH3 (**Fig. 5C**). Variants rs77008184, rs2095774, and rs17189719 in *CLDN10* have been reported to be linked to biochemical markers reflecting renal function, including serum uric acid, urate, and cystatin C levels^46,47^. The DAFs of rs77008184 and rs17189719 decreased gradually during the transition from hunter-gatherers to farmers and after the expansion of steppe pastoralists, whereas rs2095774 exhibited the opposite trend (**Fig. S16B–D**). Multiple variants in *LINGO2* that exhibited strong LD demonstrated significant AF deviations in CAAH4 (**Figs. 5D, E, and S16E**). Extended haplotype homozygosity (EHH) curves suggested an increased likelihood of local adaptation (**Fig. S17**). These variants have been reported to be associated with body mass index (rs1412235, rs2183824, rs13288841, rs7873025, and rs16912921), body fat percentage (rs10968577), obesity (rs10968576 and rs1412239), the waist-hip ratio (rs2183825), and high-density lipoprotein cholesterol (HDL-C) levels (rs1412234). The *HSD17B12* gene encodes an enzyme involved in the conversion of steroid precursors, plays a critical role in the biosynthesis of various steroid hormones, and harbors several functional loci with AF deviations in CAAH4 (**Fig. 5D**). Variants rs1061810 and rs7928523 in *HSD17B12* have been reported to be associated with type 2 diabetes and coronary artery disease, respectively^48^. The observed frequencies of rs1061810-A (0.07 vs. 0.36) and rs7928523-T (0.09 vs. 0.36) were lower than the expected post-admixture frequencies.

Post-admixture adaptive evolution was detected in genes associated with immune system pathways. The signals of natural selection in the *TRAF1* gene, encoding a protein integral to the immune response, inflammation, and apoptosis, were identified in CAAH1, which exhibited strong LD (**Figs. 5F and S18A**). Among them, rs2416804 has been reported to be associated with white blood cell count^49^, rs1930781 with asthma^50^, and rs10435844/rs3761847 with rheumatoid arthritis (RA)^51^. The DAFs for rs2416804, rs10435844, and rs3761847 demonstrated a downward trend preceding the agricultural era and following the emergence of nomadic pastoralism, whereas rs1930781 displayed an inverse pattern (**Fig. S18B–E**). Adjacent to *TRAF1*, the *PHF19* gene also harbored several functional loci with AF deviations in CAAH1 (**Figs. 5F and S18A**). Variants rs1953126, rs881375, and rs2072438 are linked to RA^52^, and rs10760123, rs6478484, and rs10818483 are associated with allergic diseases, such as asthma, hay fever, and eczema^53^. Additionally, rs7858209 has been linked to lymphocyte count^47^. These variants, except for rs2072438 and rs7858209, mirrored the DAF trend observed for *TRAF1*-rs1930781, increasing prior to the agricultural period and again following the rise of nomadic pastoralism (**Fig. S18F–L**). The EHH decay for *TRAF1*-rs2416804/rs3761847 and *PHF19*-rs2072438/rs7858209 suggested that their ancestral alleles may have undergone strong positive selection, resulting in significant frequency differences between geographically distinct CAAH groups and between CAAH and other continental populations (**Fig. 5G–N**). Notably, selection statistics (X) for ancient DNA time-series data^54^ further supported evidence of natural selection in these *TRAF1* and *PHF19* variants (**Fig. 6A**). The *MTAP* gene plays a critical role in polyamine metabolism, with the rs871024 and rs7023329 variants identified in CAAH2 being linked to melanoma^55^. Evidence for natural selection acting on rs871024 has been demonstrated through the analysis of ancient DNA time-series data (**Fig. S19A**), revealing a marked increase in DAF during the transition from a hunter-gatherer lifestyle to a farming lifestyle, followed by stabilization during the pastoral era (**Fig. S19B**). Notably, the DAF of rs871024-C exhibited substantial geographical variation, with its highest frequency detected in the HZAF and South Asian reference populations (**Fig. 6B**). The variant rs10896794 in *LPXN*, identified in CAAH4, is associated with inflammatory bowel disease^56^. The AF_obs_ of rs10896794-T (0.50) was lower than the post-admixture AF_exp_ (0.88). Additionally, the variant rs480143 in *PPP4R1L* has been demonstrated to be associated with white blood cell count^49^. The *GIMAP6* gene, recognized for its involvement in lymphocyte survival, autophagy, and immune homeostasis, contains several variants exhibiting AF deviations in CAAH4. Notably, the variant rs62491814 in *GIMAP6* has been reported to be linked to C-reactive protein levels^57^. Furthermore, rs13234724 and rs13226190 within *GIMAP6* have been associated with fibrinogen levels^58^.

**Figure 6.**
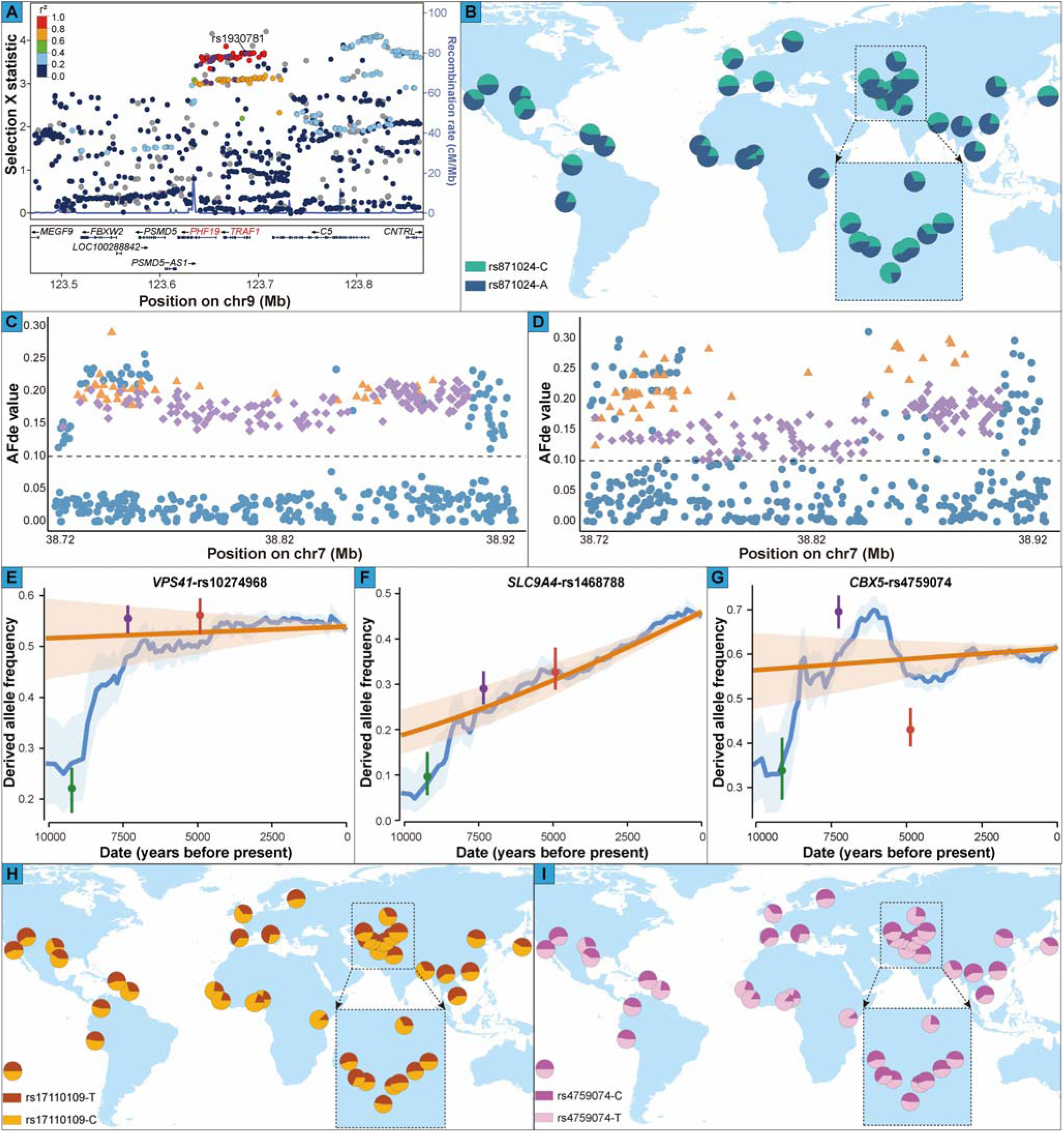
Selection plot, frequency distribution patterns, and trajectories of derived allele frequencies (DAFs) for specific local adaptive signatures. (**A**), Selection plot of *TRAF1* and *PHF19* variants among ancient West Eurasian populations, alongside linkage patterns within the target region centered on rs1930781, which demonstrated the highest AFd_e_ among natural selection signals identified for *TRAF1* and *PHF19* variants with phenotypic effects reported in the GWAS Catalog. The plot was generated via selection X-statistic values derived from ancient West Eurasian populations (Harvard Dataverse: https://doi.org/10.7910/DVN/7RVV9N). (**B**), AF distributions of *MTAP*-rs871024 across global populations. The AFd_e_ distribution of variants within the *VPS41* gene across CAAH2 (**C**) and CAAH3 (**D**). The loci marked in purple represent highly differentiated variants (HDVs) within *VPS41*, exhibiting a West Eurasian-biased frequency in CAAH2 or CAAH3. The orange-marked loci indicate HDVs with East Eurasian-biased frequencies in CAAH2 or CAAH3. Loci, marked in blue, denote variants within the VPS41 gene that lack signals of natural selection. The DAF trajectory over time for *VPS41*-rs10274968 (**E**), *SLC9A4*-rs1468788 (**F**), and *CBX5*-rs4759074 (**G**). AF distributions of *CBX5*-rs17110109 (**H**) and *CBX5*-rs4759074 (**I**) across global populations.

Post-admixture adaptive evolution has been observed in pathways associated with nervous system function and disease. For example, the variant rs4708181 in *FILIP1*, identified in CAAH2, CAAH3, and CAAH4 (**Fig. 5B–D**), has been linked to cortical surface area^59^. The *CHRNA3* gene, which is involved in pathways related to postsynaptic nicotinic acetylcholine receptors and transmission across chemical synapses, contains several functional loci exhibiting AF deviations in CAAH2 (**Fig. 5B**). Specifically, the variant rs112878080 in *CHRNA3* has been associated with the diffusing capacity of carbon monoxide^60^. Additionally, nine variants within this gene have been connected to pulmonary function measures, including the FEV1/FVC ratio and post-bronchodilator FEV1^61^. The trajectories for rs112878080, rs138544659, and rs8040868 indicated that their DAFs had been declining about 5,500 years ago (**Fig. S19C–E**). Several variants in *HDAC9*, also identified in CAAH2, were linked to brain structure or disease (**Fig. S20A**); rs10237280, rs6461386, and rs12700001 were associated with cortical surface area^62^, rs13242758 with cortical thickness^63^, rs12700002 with vertex-wise cortical thickness^63^, and rs10237366 and rs6951745 with whole brain restricted diffusion^64^. Additionally, rs67248060, rs7801037, rs6461387, and rs2073963 in *HDAC9* were reported to be related to male-pattern baldness^65^. The derived alleles of these variants (except rs13242758 and rs2073963) have been steadily decreasing in frequency from about 9,000 to 6,000 years ago and plateaued after the subsistence strategy shifted to nomadic pastoralism (**Fig. S20B–J**), whereas the DAFs of rs13242758 and rs2073963 have been steadily rising from approximately 9,000 to 6,000 years ago (**Fig. S20K and L**). The *SPON1* gene, encoding spondin 1 crucial for axonal guidance and neuronal development, features the variant rs12575169 identified in CAAH3, which is linked to adolescent idiopathic scoliosis^66^, and rs2618516, which is associated with brain connectivity^67^. Strong selection was also observed in *TCF4*, which regulates gene expression during nervous system development. Variants rs2924328 and rs2958162 in *TCF4*, identified in CAAH4, were linked to neuroticism^68^, rs2919451 to risk-taking behavior^69^, rs1452788 to depression, rs7231748 to irritable mood^70^, and rs7228159 to feelings of worry^70^. The DAFs of rs2919451 and rs2924328 exhibited a steady increase beginning approximately 8,000 years ago, followed by a notable decline starting approximately 3,000 years ago (**Fig. S20M and N**).

The AFd_e_ of highly differentiated variants (HDVs) between East and West Eurasians was further examined. The Han and French populations from the Human Genome Diversity Project (HGDP) served as ancestral references for identifying HDVs. SNPs with an Fst value exceeding 0.2 were classified as HDVs, yielding a set of 369,407 variants. HDVs with an AFd_e_ greater than 0.1 in CAAH indicated potential adaptive significance. Notably, several HDVs within the *THOP1* gene, which influences total testosterone and low-density lipoprotein cholesterol levels, were found to be under selection in CAAH1 (**Fig. 5A**). The EHH curves for these variants suggested a heightened likelihood of local adaptation (**Fig. S21A–E**). Over 170 HDVs in *VPS41* exhibited significant AF deviations in CAAH2 and CAAH3 (**Fig. 5B and C**), with rs6943746 and rs10274968 linked to neuroticism and depression, respectively. Most putative natural selection signals within *VPS41* displayed a West Eurasian-biased frequency in CAAH2 and CAAH3. However, candidate HDVs exhibiting a greater AFd_e_ in CAAH3 demonstrated a pronounced East Eurasian bias (**Fig. 6C and D**). The DAF of rs10274968 gradually increased between 9,000 and 7,000 years ago, with minimal changes before reaching a plateau approximately 4,000 years ago (**Fig. 6E**). Positive selection was further detected in several genes related to nervous system function and disorders. Mutations in *ALDH1A2* found in CAAH2 and CAAH3 have been associated with developmental disorders of the central nervous system and tissue maintenance, including whole brain restricted diffusion and osteoarthritis^71^. Candidate HDVs in *CTNNA2* in CAAH2 have been linked to various neurodevelopmental disorders, including multisite chronic pain^72^, whereas candidate HDVs in *PLCL1* identified in CAAH3 have been associated with bipolar disorder, depression, schizophrenia, and cognitive ability (**Fig. 5B and C**). The variant rs1375144 in *DPP10*, identified in CAAH3, has been linked to bipolar disorder, and variants in *PLCH1*, *PTPRD*, and *FGF14* identified in CAAH4 have been associated with insomnia^73^ (**Fig. 5C and D**). Significant signals of biological adaptation observed in CAAH groups have also been correlated with immune-related conditions^74^. For example, *CSMD1*-rs1529316 in CAAH2 and CAAH4 has been linked to multiple sclerosis (MS), *SGCZ*-rs9886428 in CAAH2 has been linked to IgG glycosylation, *IL12B*-rs3213094 in CAAH2 has been linked to psoriasis, *TLR1*-rs5743614/rs6531663 in CAAH3 has been linked to atopic dermatitis and allergic disease, and *SLC9A4*-rs1468788 in CAAH4 has been linked to celiac disease. The DAF of rs3213094 rapidly increased as subsistence strategies transitioned from hunting-gathering to agriculture, followed by a significant decrease preceding the advent of nomadic pastoralism (**Fig. S21F**). The derived variant of rs5743614 has decreased in frequency since approximately 6,000 years ago, whereas the DAF of rs6531663 has increased over the same period (**Fig. S21G and H**). Notably, the frequency of rs1468788 has steadily increased over the past 10,000 years (**Fig. 6F**). Furthermore, these immune-related variants presented significant geographic variation in frequency (**Fig. S22**). Positive selection was also detected at several loci associated with alopecia, notably a CAAH3-specific locus (*ITPR1*-rs13313995) and a CAAH4-specific locus (*CBX5*-rs17110109)^65,75,76^ (**Fig. 5C and D**). Moreover, a candidate HDV (*CBX5*-rs4759074), linked to meat-based diets, displayed a notable AF deviation (0.20) in CAAH4. The DAF of rs4759074 peaked approximately 6,000 years ago, followed by a brief decline before the onset of nomadic pastoralism (**Fig. 6G**). Notably, these two *CBX5* variants presented marked differences in AF between CAAH4 and other CAAH groups, as well as across continental populations (**Fig. 6H and I**).

### Archaic introgression and possible biological functions in human health and disease

The extent of archaic introgression from extinct hominins, both known and unknown, in Central Asian populations remains underexplored. To address this gap, IBDmix was first applied to infer archaic introgression segments (AISs) from the Altai Neanderthals. The distribution patterns of Neanderthal-derived sequences in CAAH1, which exhibited over 50% Western Eurasian ancestry, aligned closely with reference European populations from the 1000 Genomes Project (1KGP); the distribution patterns in CAAH2, which possessed approximately half of the Western Eurasian ancestry, mirrored those observed in South Asian populations (**Fig. 7A**). In contrast, CAAH3 and CAAH4, characterized by greater East Asian ancestry, presented the shortest number of Neanderthal-derived AISs per individual, likely due to sample size bias^77^. On average, each CAAH individual carried approximately 50.01 Mb of Neanderthal-derived sequences, with mean lengths ranging from 52.95 Mb in CAAH1 and CAAH2 to 39.23 Mb in CAAH3. Notably, 104 Neanderthal-introgressed regions were not identified by the 1KGP (**Fig. 7B**), encompassing the *GUSBP1*, *ASTN2*, *ARHGAP15*, *CYP39A1*, and *MAN1A2* genes, each covering more than 1,000 archaic variants that do not exhibit significant phenotypic effects. Additionally, the *BCL11A* segment, which is specific to CAAH, is associated with the development of hematopoietic cells and various hematological disorders, including sickle cell anemia. Notably, 54 Neanderthal-derived sequences were unique to specific CAAH subpopulations: 27 in CAAH1, 18 in CAAH2, 4 in CAAH3, and 5 in CAAH4, with no significant phenotypic effects reported (**Fig. 7B**). The Sprime tool was also utilized to infer AISs from the Altai Neanderthal and Denisovan genomes. We integrated the results from both methods to obtain high-confidence Neanderthal-derived segments for identifying adaptive introgression signals. A total of 1,605 segments likely introgressed from Neanderthals were identified, spanning 392.51 Mb of the genome, along with 132 Denisovan-derived segments, covering 37.17 Mb (**Fig. 7C**). Each individual, on average, carried approximately 13.24 Mb of high-confidence Neanderthal-derived sequences and 0.34 Mb of Denisovan-derived sequences (**Fig. 7D**). These AISs are attributed to a single wave of Neanderthal admixture and two distinct waves of Denisovan admixture across these groups (**Figs. 7E and S23**).

**Figure 7.**
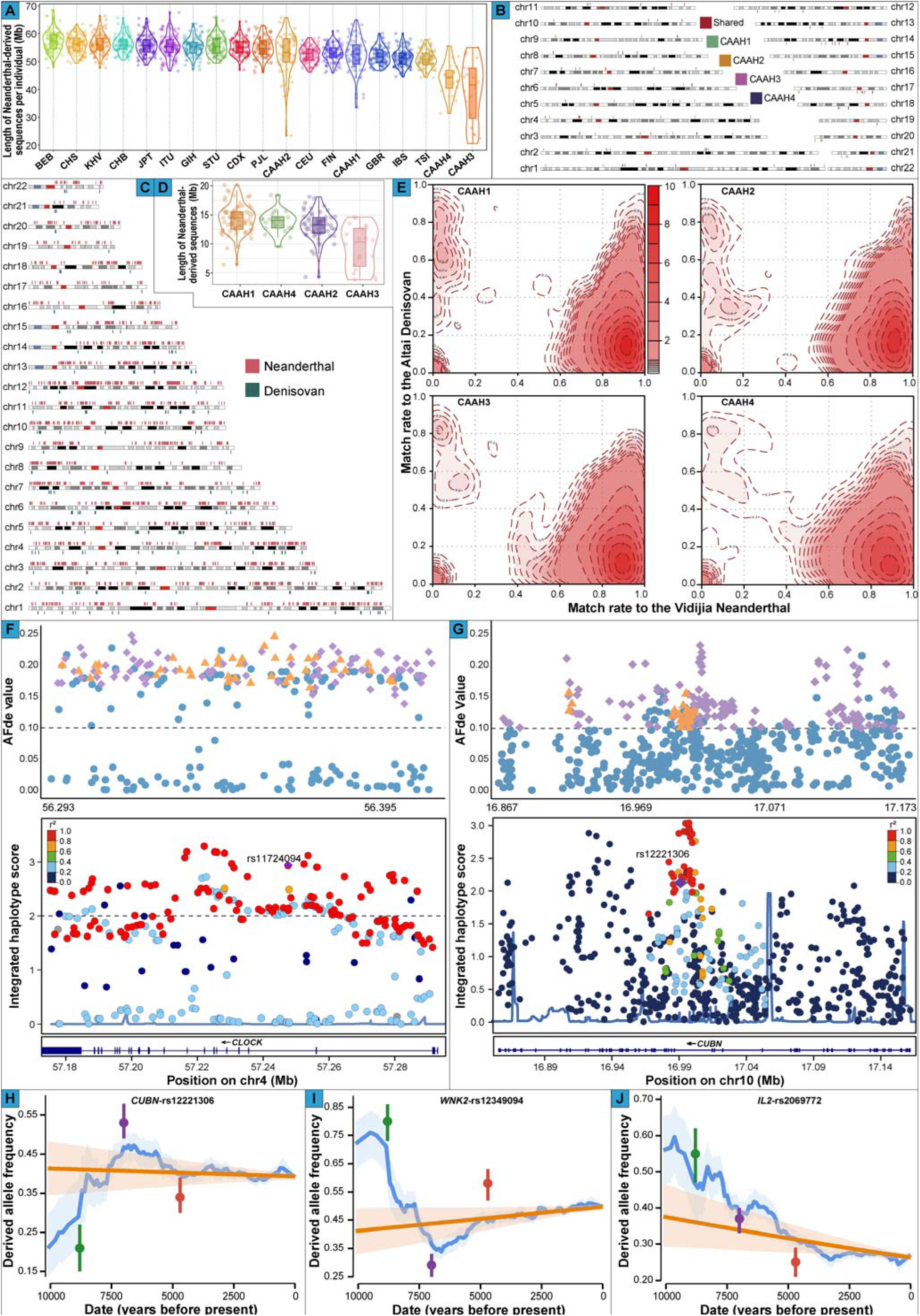
Distribution of Neanderthal- and Denisovan-derived archaic introgression segments (AISs) in the CAAH groups. (**A**), Distribution of Neanderthal-derived AISs identified via the IBDmix method across Eurasian populations. (**B**), Distribution of Neanderthal-derived AISs that are unique to CAAH groups and absent in Eurasian populations in the 1000 Genomes Project (1KGP). The AISs shared among the four CAAH groups are depicted in dark red, whereas those unique to CAAH1, CAAH2, CAAH3, and CAAH4 are represented in dark green, orange, purple, and dark blue, respectively. (**C**), Distribution of high-confidence Neanderthal- and Denisovan-derived AISs on 22 autosomes. (**D**), The distribution of high-confidence Neanderthal-inherited AISs across individuals within each CAAH group, as determined by IBDmix and Sprime. (**E**) Contour density plots of the matching proportions of introgressed fragments to the Neanderthal and Denisovan genomes. (**F**) The distribution of AFd_e_ and integrated haplotype score (iHS) for potential Neanderthal-introgressed variants in the *CLOCK* gene identified in CAAH1. The figure above illustrates the distribution of the AFd_e_. The orange triangles indicate signals of adaptive introgression, the purple diamonds represent signatures of natural selection, and the dark blue diamonds denote nonadaptive introgression and non-natural selection signals in the *CLOCK* gene. The figure below shows the distribution of iHS values and the linkage patterns within the target region centered on rs11724094. The variant positions in these two figures correspond. (**G**) The distribution of AFd_e_ and iHS values for potential Neanderthal-introgressed variants in the *CUBN* gene identified in CAAH1 and the linkage patterns within the target region centered on rs12221306. The variants indicated by different colors in this figure correspond to those shown in Figure 7F. The DAF trajectory over time for *CUBN*-rs12221306 (**H**), *WNK2*-rs12349094 (**I**), and *IL2*-rs2069772 (**J**).

The phenotypic effects of the identified high-confidence Neanderthal- and Denisovan-derived segments were systematically examined. For Neanderthal-derived sequences, genes related to immune function (*TLR1/10*, *OAS1/2/3*, *CCR1/2/3/5*, *CCRL2*, *ERAP2*, *DOCK2,* and *LRRC2*), metabolism (*ZNF444*, *PASK*, *OSBPL10*, *HDLBP*, and *TBC1D1*), dermatological or pigmentation traits (*LAMB3* and *TMEM132D*), skin keratinocyte differentiation (*POU2F3*), neurological characteristics (*ADAMTSL3*, *TENM3*, and *UNC13C*), and reproductive health (*PGR*) were replicated^19,78–80^. Novel signals related to immunity-related (*CXCR5*, *TLR5*, *PLCL2*, *RAB17*, *USP20*, *TRIML1*, *PIBF1*, *C6*, and *HHLA1*) and metabolism-related (*DDC*, *FAH*, *MTRR*, *ASRGL1*, *ACSBG2*, *ALK*, and *CRTC3*) genes were identified. For Denisovan-derived sequences, immune-related genes such as *ARHGEF28*, *BANK1*, and *EPHB2* were replicated, with previously unreported signals in immunity-related (*C1QA*, *C1QB*, *C1QC, GIMAP4/6/7*, *LECT2*, *DEF6*, *XRCC4*, and *PRKCE*) and metabolism-related (*TCF7L2*, *A1CF*, *GOT2*, *FGF1*, *PRKCE*, *PPARD*, *HMGCR*, *RHOQ*, *TBXAS1*, *SLC38A7*, *ALPI*, and *ALPP*) genes. Significant phenotypic associations for Neanderthal-derived segments across CAAH groups were identified at *BUD13*, *ZPR1*, and *APOA5,* which were correlated with lipid metabolites. Associations were also observed between *ARHGEF12* and glaucoma and intraocular pressure^81^; *SGCZ* with psychiatric traits^68^; and *GRB10* with blood glucose, insulin levels, and diabetes^82^. Population-specific Neanderthal-derived sequences, including the *L3MBTL3* segment in CAAH1, which is associated with lipid metabolite levels through the modulation of adipocyte differentiation, have been identified^83^. The *RAD51B* segment in CAAH1, which is crucial for DNA repair via homologous recombination, is linked to immune-related disorders such as RA and asthma^52^. Additional segments within CAAH1 (*ESR1*) and CAAH2 (*PPP6R3*) are associated with bone mineral density^84^. The Neanderthal-inherited *MAD1L1* segment in CAAH2 is associated with psychiatric conditions, including schizophrenia and depression^85^. The *SLCO1B1* segment in CAAH3, encoding organic anion transporting polypeptide 1B1, is linked to hepatic uptake of clinically utilized drugs, such as statins and methotrexate, alongside endogenous compounds, including bilirubin, biliverdin, and deoxycholic acid. The *UBE2L3* fragment in CAAH4 is associated with serum lipid levels and autoimmune disorders, such as Crohn’s disease, inflammatory bowel disease, and systemic lupus erythematosus (SLE)^86^. Neanderthal-derived segments were further implicated in various diseases, including cardiovascular diseases, gout, respiratory conditions such as chronic obstructive pulmonary disease, and autoimmune disorders such as MS. Phenotypic associations of Denisovan-derived sequences were predominantly noted in genes linked to height, type 2 diabetes, and Takayasu arteritis. Additionally, archaic variants in *HHAT* (rs115453328) identified in CAAH1 were associated with obsessive-compulsive traits; variants in *RASA3* (rs61971965) in CAAH3 were associated with grey matter density; and variants in *IL2RA* (rs61839660) in CAAH4 were associated with immune-related conditions, including lymphocyte count and allergic disease. Pathway analysis revealed that several biological pathways related mainly to muscle cell differentiation, the nervous system, and muscle tissue development were shared between Neanderthal- and Denisovan-derived sequences (**Fig. S24**).

Evidence for archaic adaptive introgression has been established^16^; however, few studies have evaluated its impact on Central Asian and adjacent populations. Here, adaptive introgression signals in CAAH were examined via high-confidence AISs and signatures of local adaptation. Notably, no significant signals of Neanderthal-derived adaptive introgression were detected in CAAH2, nor were any notable Denisovan-inherited adaptive introgression signals observed among the CAAH groups. A Neanderthal-derived haplotype at *CLOCK* within CAAH1 was identified, consisting of 42 archaic SNPs (aSNPs), with rs11724094 linked to male-pattern baldness (**Fig. 7F**). The variant rs12221306 at *CUBN*, associated with metabolite levels, and rs12349094 at *WNK2*, associated with educational attainment, were identified in CAAH1 (**Fig. 7G**). The DAF of rs12221306 increased prior to the advent of agriculture, whereas that of rs12349094 exhibited the opposite trend (**Fig. 7H and I**). Several aSNPs linked to immune conditions have also been identified in CAAH1, including rs6445975 at *PXK*, which is associated with SLE; rs2069772 at *IL2*, which is linked to allergic sensitization^87^; and rs1929863 at *CDC42BPA*, which is correlated with white blood cell count. Notably, the DAF of rs2069772 has decreased since 10,000 years ago, suggesting that environmental factors, including changes in pathogen exposure, may influence the necessity for specific immune-related variants (**Fig. 7J**). Several Neanderthal-like adaptive haplotypes containing multiple aSNPs were identified in CAAH3 and CAAH4; however, no significant overall phenotypic effects were observed. The variant rs73187291 at *DGKH* in CAAH3 was associated with insulin-like growth factor 1 levels^46^, whereas rs12634, a 3’ UTR variant of *TOR1B* identified in CAAH4, has been linked to prostate cancer. Overall, archaic introgression did not have a significant effect on the genetic basis of local adaptation in Central Asian populations.

## Discussion

Central Asia has served as a critical region for multidirectional human dispersal across Eurasia in prehistoric and historical periods. It has been a melting pot of trans-Eurasian genetic, economic, and cultural exchanges, playing a pivotal role in shaping the genomic diversity patterns of geographically diverse Eurasians. Despite the considerable importance of Central Asia in the formation of mosaic genetic structures of present-day Eurasians, our knowledge of the demographic processes of contemporary Central Asians and the genetic architecture underlying their specific traits and diseases has been limited. Our comprehensive genomic analysis of 166 individuals from 20 Central Asian populations in the CAGDP revealed significant genetic diversity and complex patterns of ancestral lineages. Through WGS, we identified 22,994,949 SNPs and 2,952,662 small InDels with many novel variants, particularly at low frequencies. PCA and clustering analyses revealed genetic diversity among Central Asian populations, with distinct ties to West and East Eurasians. Uzbek and Turkmen were more closely related to West Eurasians, whereas Dungan was more closely related to East Eurasians. Tajiks and Karluks had mixed Indo-European and Turkic links, and Kyrgyz, Uyghur, Karakalpak, and Hazara clustered with Turkic groups. Dungan exhibited genetic connections with Mongolia, Tibet, and Nepal, reflecting historical migrations. Ancient DNA analysis revealed non-Dungan groups’ genetic ties to Iron Age Xinjiang and historical Central Asians, whereas Dungan shared links with ancient Yellow River and Qinghai□Tibet populations. Turkic-speaking populations presented geographically distinct genetic clusters, with recent gene flow in southwestern, southern, and northeastern Siberia. The genetic profiles of the Dungan and Hazara populations were influenced by long-distance migrations, with Dungan sharing alleles with East Asian groups and Hazara showing Uyghur-like admixture and Tungusic links. Pathogenic variants associated with cardiac conditions were identified, and pharmacogenomic analysis highlighted AF differences affecting drug metabolism, emphasizing personalized medicine. Positive selection signatures were found in genes related to metabolism, diseases, and immunity. Archaic introgression contributed to the genetic makeup of Central Asians, with Neanderthal and Denisovan genetic material influencing phenotypes and diseases, suggesting adaptive advantages. This study underscores the importance of diverse genomic research for understanding human genetic diversity and its health implications.

### Fine-scale population structure of CAAH

The fine-scale population structure of CAAH shows significant genetic heterogeneity, as revealed by PCA, ADMIXTURE, outgroup *f_3_*-statistics, and TreeMix analyses. TJK, KLKTL, TKM, and UZTK individuals shared more alleles with Indo-European and Turkic groups from West Eurasia, as well as with Iranian farmers, Anatolian farmers, and WSH-related ancestral populations. In contrast, the KGZ, UGAMT, KRKP, and HZAF populations presented genetic similarities with Turkic groups in Central Asia, Altaic groups in northern East Asia and southern Siberia, and ancient individuals from the MP with predominantly East Eurasian ancestry. Moreover, the DGJB group presented excess allele sharing with the Sino-Tibetan, Mongolic, and Turkic people in East Asia and ancient populations from the YRB, QXP, and Nepal. Ancient DNA studies have demonstrated that ancestral groups related to WSHG, EEHG, WSH, ANE, Iranian farmers, and Anatolian farmers were widespread in Central Asia from the Paleolithic period to the historical period^26–31^. High-resolution population admixture modeling confirmed previous population genetic discoveries. Yunusbayev *et al*. conducted an array-based study that demonstrated that the Kyrgyz, Uzbek, and Turkmen populations share more alleles with geographically diverse East Asian and Siberian populations than with West Eurasians do^33^. Studies on Uyghurs in Xinjiang have identified four primary ancestral components: European (approximately 25–37%), South Asian (12–20%), East Asian (29–47%), and Siberian (15–17%) ancestries^36^. To investigate the genetic legacy of these ancient populations in modern Central Asians, we performed admixture modeling via ADMIXTURE, *f_3_*-statistics, *f_4_*-statistics, and qpAdm. With the optimal K=11 model, CAAH can be modeled as a mixture of ANA, West Eurasians (Anatolian farmers, Iranian farmers, and EEHG), and East Asians (NEA, SEA, and Japanese_Tokyo1K), with varying proportions. TJK, KLKTL, UZTK, and TKM contained higher West Eurasian ancestral components, whereas KGZ, KRKP, HZAF, and UGAMT had more ANA-related ancestries, and DGJB harbored predominantly East Asian-derived ancestries. The admixture *f_3_* values indicated that most CAAH groups resulted from east-west admixture, except for KLKTL and DGJB, which presented more complex admixture scenarios. The qpAdm models further refined these findings: TJK, KLKTL, TKM, and UZTK derived most of their ancestry from a West Eurasian nomadic group (67%–86%) and a smaller proportion from an East Eurasian nomadic group (14%–33%). UGAMT, HZAF, and KRKP could be modeled as an equal mixture of West and East Eurasian nomadic groups, whereas KGZ had approximately 70% East Eurasian ancestry and 30% West Eurasian ancestry. DGJB derived most of their ancestry from ANEAs, with minor contributions from Tajikistan_Ksirov_Kushan (<10%). Haplotype-based analyses via SOURCEFIND and fastGLOBETROTTER confirmed these differentiated east-west admixture processes among CAAH. Our findings challenge historical assumptions. The Karakalpaks in Uzbekistan, thought to have emerged from the Kazakh Confederation’s division in the 17th century^88^, show more allele sharing with Turkic groups in southern Siberia and eastern Central Asia than with nearby Turkic groups. Two Karluks males buried at Butakty in the Tian Shan between 800 and 1,000 AD carried the paternal lineage of J2a and the maternal lineages of A and F1b1e^30^. The Karluks, a prominent nomadic confederacy, showed a mix of ancestries, with KLKTL having over 70% West Eurasian ancestry but East Asian-typical paternal and maternal lineages. Modern Central Asians exhibit remarkable genetic diversity, with some groups, such as TJK, being predominantly West Eurasian, whereas others, such as DGJB, are largely East Eurasian. These patterns likely reflect historical Turkicization by East Asian elites and the eastward migration of West Eurasians during the Türkic period, leading to the complex genetic landscape observed today^30^.

Estimates of admixture dates, effective population sizes, and population divergence times provide insights into the distinct demographic history of Central Asians and their neighbors. The analysis of admixture-induced LD decay suggested that east-west admixture in CAAH occurred approximately 23□34 generations ago (approximately 667□986 years ago), corresponding to the Song and Yuan Dynasties and overlapping with the rise and fall of the Mongol Empire. At its peak, the Mongol Empire encompassed nearly two-thirds of Eurasia. The empire’s westward expansion increased East Asian ancestry in Central Asia, significantly altering the political and genetic landscape of Eurasia. Haplotype-based estimates indicated similar admixture times for CAAH populations, except for TJK. TJK presented evidence of an earlier east-west admixture approximately 50 generations ago (around the Tang Dynasty) and a more recent admixture approximately nine generations ago (during the Qing Dynasty). Historical records and ancient DNA studies suggest that population expansion due to agriculture and nomadism intensified east-west admixture beginning in the BA era, implying that the estimated dates of admixture in CAAH might be conservative^32^. The dynamic patterns of Ne and divergence times further reveal complex admixture processes and distinct demographic histories within CAAH populations. These findings underscore the intricate interplay of historical migrations and population expansions in shaping the genetic diversity of modern Central Asian populations.

### The differentiated genetic architecture of linguistically close Turkic, Indo-European, and Sino-Tibetan groups

The patterns of population structure revealed through allele and haplotype sharing revealed several geographically distinct Turkic clusters. These clusters include populations primarily from the Caucasus, Eastern Europe, and Western Siberia; others from Siberian regions and Northeast Central Asia; another group from various Central Asian regions; and a final cluster primarily from East Asia, particularly recent migrants from the Gansu--Qinghai region. The proportions of different ancestral components in these Turkic groups showed distinct correlations with longitude and latitude. Populations farther west presented higher proportions of Anatolian farmers, Iranian farmers, and EEHG-related ancestors. In contrast, those further east had more East Asian, ANA, and Nganasan-related ancestries. Similarly, Turkic groups further north presented greater EEHG, ANA, and Nganasan-related ancestries, whereas those farther south presented greater East Asian and Iranian farmer-related ancestries. The origin of the Turkic people has long been a topic of debate. Some theories propose a South Siberian and Mongolian origin, supporting the “steppe hypothesis. “^33^ Others suggest a Northeast China origin, linking the Turkic people to ANEAs associated with the Xinglongwa and Hongshan cultures, supporting the “farming hypothesis“^89^. Nevertheless, others argue for an origin in the ARB, supporting the “Northeast Asia origin“^90^. Our findings indicate that geographically distinct Turkic groups possess varying proportions of ANA-related ancestry, ranging from 1.1% to 34.5%, with Central Asian Turkic groups showing 7.6% to 28.9% ANA-related ancestry. Furthermore, our modeling suggests that the newly sequenced Turkic people can be characterized as a mixture of China_AR_Xianbei_IA (approximately 29.7% to 72.3%) from the ARB and a WSH-derived population. In summary, our results support the “Northeast Asian origin” of the Turkic people, who adopted a nomadic lifestyle and migrated to Central Asia via the Eurasian steppe.

Our analysis identified two Indo-European genetic clines: one primarily encompassing individuals from the Caucasus and Central Asia and the other comprising people from Eastern Europe. The Caucasus and Eastern European Indo-European groups presented a significantly greater proportion of Anatolian farmer-related ancestry than did the Indo-European groups in Central and South Asia. In contrast, Indo-European individuals in the Caucasus, Central, and South Asia presented a greater proportion of Iranian farmer-related ancestry than did those in Eastern Europe. The distribution of EEHG-related ancestry in Indo-European populations was inversely correlated with that of Iranian farmer-related ancestry. The population structure patterns suggest that the TJK population shares more alleles with Iranian farmers, Anatolian farmers, WSH-related groups, and most ancient individuals from the Turan (excluding Kyrgyzstan). Admixture models revealed that TJK derived more than 80% of its ancestry from West Eurasian sources, with additional contributions from NEA, Japanese_Tokyo1K, Nganasan, ANA, and SEA. A recent study highlighted the genetic diversity among Tajiks and revealed that Tajik populations shared more alleles with several BA individuals from Xinjiang than with those from Central Asia^25^. This suggests that Tajiks could be modeled as a mixture of Anatolian farmers, Iranian farmers, ANE, Western European hunter-gatherers, ancient Southern East Asians, and East Siberian hunter-gatherers. These findings indicate possible genetic differentiation among geographically diverse Tajik populations or potential biases due to differences in the reference populations used. Notably, the Indo-European-speaking HZAF was distinct from the Indo-European genetic clines identified. As one of Afghanistan’s largest ethnic groups, the origins of Hazaras remain incompletely understood. Some studies suggest that the Hazara people are closely related to Turkic groups from Central Asia rather than East Asian or Indo-European-speaking populations^34^. However, other studies have demonstrated a strong genetic affinity between Hazaras and East Asians, particularly Mongolian-related populations^91^. Our findings indicate that HZAF shares more alleles with Turkic groups in Central Asia (e.g., UGAMT and KRKP), Altaic groups in northern East Asia and southern Siberia, and ancient populations from the ARB, WLR, and YRB. Additionally, HZAF shares longer IBD segments with Altaic groups in Siberia than with populations that are geographically or linguistically close. These findings suggest that ancient long-distance migrations and recent admixtures have reshaped the genetic profile of Hazara. Similarly, the DGJB is genetically closer to modern East Asians, particularly Sino-Tibetan speakers and ancient populations from the YRB, than to geographically proximate Central Asian groups. Historical records describe Dungan as a group of Muslim people of Hui origin who migrated from China to Central Asia. Our observations, along with recent findings showing no significant genetic differentiation between Dungan and southern Ningxia Hui populations, suggest that long-distance migrations of Sino-Tibetan-related East Asians have shaped the unique genetic structure of Dungan in Central Asia^92^.

### Central Asian genomes inform human health

The analysis of the newly generated Central Asian genomic dataset has significantly improved our understanding of how genetic variations influence the genetic architecture of Mendelian diseases, susceptibility to complex common diseases, and drug response in Central Asian populations. We identified 116 pathogenic variants, with approximately 73.3% being rare (MAF < 0.05). More than half of the sequenced individuals carried at least one of the ten reportable ACMG variants, and approximately 39.2% carried the *SCN5A*-rs1805124 variant associated with ventricular fibrillation. This finding suggests a need to revise the ACMG variant interpretation guidelines. Additionally, we identified 3,436 predicted pLoF variants, nearly all of which (∼98.4%) were classified as PTVs. Notably, approximately 76% of these PTVs were unique to our dataset. We detected 129 homozygous PTVs, including eight novel PTVs, expanding the catalog of variants that appear to tolerate homozygous loss of function. These findings provide essential insights into the biological functions of these variants, especially when combined with phenotypic information. Overall, our dataset significantly improves the ability to identify pathogenic variants in Central Asian populations and better assess the pathogenicity of these variants.

Reconstructing the AF spectrum of known pharmacogenomic variants in genetically diverse populations is crucial for enhancing drug efficacy, preventing adverse drug reactions, guiding personalized medication, and advancing precision medicine. Our analysis revealed significant differences in the AFs of certain variants that affect drug response among CAAH populations, as well as between CAAH and other intercontinental populations. Notable examples include variants in *VKORC1*, *CYP2C9*, and *CYP4F2* related to warfarin metabolism; *ATIC* associated with methotrexate response; *ADRB2* related to salmeterol response; and *NAT2*5*, *NAT2*6*, and *NAT2*12* associated with the isoniazid response. These findings underscore the need for preemptive pharmacogenetic testing before drugs are prescribed to genetically diverse Central Asian populations to ensure optimal therapeutic outcomes.

### Signatures of local adaptation

The effects of admixture, selection, and archaic introgression on human health and disease are critical issues in human genetics. Analyzing the genetic basis of local adaptation in CAAH populations enhances genome-wide scans for positive selection signatures in Central Asian populations. Significant AF deviations were observed across CAAH groups, indicating distinct evolutionary pressures following admixture. Local adaptation signals were detected in genes associated with metabolism, development, and immune responses. Notably, *EBF2* variants in CAAH1 and CAAH2 presented ancestry-biased components, impacting adipogenic processes and metabolic health. Additionally, *FNDC1* variants were linked to cellular hypoxia responses and cardiomyocyte apoptosis, and *THOP1* variants in CAAH1 were associated with testosterone and cholesterol levels. A large-scale GWAS of testosterone levels in the UK Biobank indicated that higher testosterone levels increase the risk of metabolic diseases in women but are protective in men^93^. Adaptive signals in *CLDN10*, which are essential for kidney function, were identified in CAAH3, whereas CAAH4 showed adaptation in obesity and metabolic regulation pathways, with *LINGO2* and *HSD17B12* variants diverging under selection. The frequency of these variants diverged significantly from the expected values, indicating that strong selection pressures are likely related to changes in subsistence strategies over time.

Adaptive evolution of immune system-related genes, particularly *TRAF1* and *PHF19*, which play crucial roles in the immune response and inflammation, was evident. Variants associated with diseases such as RA and allergic conditions identified in CAAH1 have undergone natural selection, reflecting ongoing pressures faced by these populations. Recent research indicates that RA risk variants were most prevalent in hunter-gatherer populations prior to the advent of agriculture and have been subject to negative selection over the past 15,000 years^94^. CAAH1, which is predominantly composed of individuals with significant West Eurasian ancestry—more than 15% attributed to EEHG—likely inherited RA-associated risk variants through admixture with EEHG populations (**Fig. 1E**). In CAAH2, the *MTAP* variants rs871024 and rs7023329 were associated with melanoma, with increased DAFs during the transition from hunter-gatherers to farmers. This contrasted with CAAH4, where the rs10896794 variant in *LPXN* was linked to inflammatory bowel disease and displayed a lower frequency than expected post-admixture. Variants within the *GIMAP6* gene in CAAH4 plateaued in DAFs approximately 3,000 years ago, indicating recent potential stabilization of selective pressures on immune-related traits.

Additionally, adaptive evolution related to nervous system function is prevalent across CAAH groups. The variant rs4708181 in *FILIP1*, found in CAAH2, CAAH3, and CAAH4, has been associated with cortical surface area, suggesting adaptive significance in neuroanatomical development. The presence of variants in the *CHRNA3* gene in CAAH2 highlights its connection to pulmonary function, with AFs declining for specific variants approximately 5,500 years ago, potentially reflecting historical selective pressures favoring alternative adaptive traits in response to environmental changes. In contrast, variants within the *HDAC9* gene, particularly those identified in CAAH2, demonstrate a complex selection history, with divergent DAF changes suggesting differential selection pressures linked to shifts in lifestyle or environmental conditions. In CAAH3, the *SPON1* gene is associated with adolescent idiopathic scoliosis, emphasizing the importance of nervous system adaptations. Strong selection signals in the *TCF4* gene in CAAH4, linked to neuroticism and other behavioral traits, have been observed, with increasing DAFs for rs2919451 and rs2924328 since approximately 8,000 years ago, followed by a decline approximately 3,000 years ago. The identification of variants associated with mental health and neurodevelopmental disorders underscores the complex interplay between genetics and the environment in shaping population health outcomes. Overall, these findings underscore the intricate relationships among genetic variation, adaptive evolution, and shifts in subsistence strategies within CAAH groups.

### Landscape of archaic introgression

Our findings provide critical insights into the extent of archaic introgression from Neanderthals and Denisovans in Central Asian populations. The identification of Neanderthal-derived segments, particularly within CAAH populations with varying ancestral backgrounds, suggests that genetic influences from archaic hominins differ according to regional demographic histories. The distribution of Neanderthal-derived AISs varies across CAAH groups, with CAAH3 and CAAH4 exhibiting the shortest sequences, which may reflect the influence of sample size bias or admixture dynamics^77^. The discovery of 104 novel Neanderthal-inherited segments not detected in the 1KGP underscores the complexity of introgression and its potential role in shaping genetic diversity in Central Asia. Moreover, CAAH-specific AISs, including the *BCL11A* fragment, indicate localized evolutionary pressures that may have influenced phenotypic traits such as hemoglobin levels and susceptibility to sickle cell anemia^95^.

In this study, the phenotypic effects of high-confidence AISs were systematically examined, revealing significant associations with various traits and diseases. The replication of immune-related genes, such as *TLR1/10* and *OAS1/2/3*, underscores the potential role of Neanderthal-derived sequences in modulating immune responses. The identification of novel immunity-related genes, including *CXCR5* and *TLR5*, further highlights the intricate interplay between Neanderthal-associated AISs and modern human phenotypes. The significant associations of Neanderthal-derived segments with lipid metabolism, particularly at *BUD13*, *ZPR1*, and *POA5*, suggest that these genetic contributions may influence metabolic health and the predisposition to cardiovascular diseases. Additionally, population-specific associations, including *L3MBTL3* and *RAD51B* in CAAH1 with lipid metabolite levels and immune-related disorders, respectively, *MAD1L1* in CAAH2 with psychiatric conditions, *SLCO1B1* in CAAH3 with drug metabolism, and *UBE2L3* in CAAH4 with autoimmune diseases, imply that Neanderthal-derived variants may exert distinct impacts on health outcomes across different CAAH groups.

The Denisovan-derived sequences were predominantly associated with height, type 2 diabetes, and Takayasu arteritis, indicating that these AISs may also affect phenotypic traits relevant to modern health. The identification of specific variants linked to obsessive-compulsive traits and grey matter density suggests that Denisovan ancestry may influence neurodevelopmental and psychological traits as well. Pathway analyses revealed shared biological pathways associated with muscle cell differentiation and nervous system and muscle tissue development between Neanderthal-and Denisovan-derived sequences, indicating that archaic contributions may influence not only individual traits but also broader physiological processes.

Our observations indicate that while evidence for adaptive introgression exists within Central Asian populations, its overall impact appears to be limited. The identification of a Neanderthal-derived haplotype at *CLOCK* in CAAH1, with a variant linked to male-pattern baldness, suggests potential adaptive significance. Additionally, several aSNPs related to immune conditions were found in CAAH1, indicating a possible role of these variants in shaping immune responses. The decreasing DAF of rs2069772 over the past 10,000 years suggests that environmental factors, such as changing pathogen exposures, may have driven the selection of immune-related variants. Conversely, the increasing DAF of rs12221306 prior to the advent of agriculture, alongside the decreasing trend of rs12349094, highlights the dynamic interplay between genetic variants and environmental pressures. Despite the identification of Neanderthal-like haplotypes in CAAH3 and CAAH4, their phenotypic effects were deemed insignificant. Overall, these findings suggest that while archaic introgression may contribute to the genetic landscape of genetically distinct CAAH groups, it does not seem to play a substantial role in the genetic basis of local adaptation. This underscores the complexity of genetic adaptation in response to diverse environmental factors within these populations.

### Conclusion

Central Asia has historically served as a crucial crossroads for human migration, significantly shaping the genetic landscape of Eurasia; however, its genomic resources remain underrepresented in genetic research. A comprehensive genomic analysis of 166 individuals from 19 Central Asian populations and Afghanistan Hazara revealed substantial genetic diversity and complex ancestral lineages. Admixture modeling and unsupervised clustering identified clear genetic boundaries, placing these populations at the intersection of Western and Eastern Eurasian genetic clines. Non-Dungan groups were genetically linked to spatiotemporally distinct ancient Central Asian and Siberian populations. The Tajik, Karluks, Uzbek, and Turkmen populations presented the strongest genetic affinities to Western Eurasians, followed by the Uyghur, Karakalpak, and Hazara populations. In contrast, Dungan aligned more closely with Eastern Eurasians, followed by Kyrgyz. The close genetic affinity between Dungan or Hazara and geographically distinct East Eurasians underscores the critical role of long-distance migration in shaping the genetic architecture of Central Asians. Significant genetic differentiation was observed among Turkic-speaking populations, with distinct genetic clusters corresponding to specific regions. A substantial number of pathogenic and pharmacogenomic variants were identified, revealing notable differences in AFs that influence clinical outcomes, drug metabolism, and responses, highlighting the necessity for genotype-guided drug administration. Evidence of positive selection was also documented, particularly in genes associated with metabolic traits, immune responses, and neurological disorders, suggesting the influence of local adaptation on genetic diversity. Additionally, substantial archaic introgression from Neanderthal and Denisovan sources has been recorded, contributing to diverse phenotypic traits and disease susceptibility. These findings emphasize the importance of incorporating diverse populations in genomic research to enhance the understanding of human genetic diversity and its implications for health and disease.

## Materials and methods

### Study participants and sequencing

Saliva samples were collected from 166 individuals from five Central Asian countries (Kyrgyzstan, Kazakhstan, Uzbekistan, Tajikistan, and Turkmenistan) and Afghanistan in South Asia via the Oragene DNA Self-Collection Kit (OG-500, DNA Genotek, Canada). The sampled individuals included Kyrgyz from various regions in Kyrgyzstan (Talas, Issyk-Kul, Naryn, Osh, Batken, and Jalal-Abad), Uzbeks from the Turkistan Region in Kazakhstan, Karakalpaks from the Karakalpakstan Region in Uzbekistan, Karluks from the Khatlon Region in Tajikistan, Uyghurs from the Almaty Region in Kazakhstan, Turkmen from Daşoguz and Mary Regions in Turkmenistan, Dungans from the Jambyl Region in Kazakhstan, Tajiks from several regions in Tajikistan (Sughd, Gorno-Badakhshan, and Khatlon), and Hazaras from various provinces in Afghanistan (Bamyan, Ghazni, Oruzgan, and Ghor). The participants were selected on the basis of the criterion that all four grandparents were indigenous to the respective ethnic groups. Informed consent was obtained from all participants. The study was approved by the Medical Ethics Committee of the National Centre for Biotechnology of Kazakhstan (protocol No. 2 of 10 June 2020) and the Nazarbayev University Institutional Research Ethics Committee (protocol No. 17 of 16 April 2019), and all procedures were conducted in accordance with the Declaration of Helsinki^96^. Genomic DNA was extracted via the prepIT-L2P kit (DNA Genotek, Canada) following the manufacturer’s protocol, and the DNA concentration was measured via a Qubit 3.0 fluorometer (Thermo Fisher Scientific). Library preparation was performed according to the standard Illumina protocol, with all libraries purified via Agencourt AMPure XP (Beckman Coulter). The final libraries were quantified via qPCR. WGS was conducted on the Illumina HiSeq 4000 platform, with a preset sequencing depth of 10× for all samples.

### Sequencing quality control and variant discovery

Samples were retained if at least 80% of all bases had a quality score of 30 or higher. Sentieon Genomics tools (version 202010.04) were used to process newly generated genomic data, following the GATK Best Practices guidelines^97^. The raw sequencing reads were aligned to the human reference genome assemblies GRCh37 and GRCh38 via BWA v0.7.17^98^. PCR duplicates were removed with Sentieon Dedup. The aligned sequencing reads from different lanes were then sorted and merged via SAMtools v1.10^99^. GVCF files were generated with the HaplotypeCaller module, and joint variant calling was performed via the CombineGVCFs and GenotypeGVCFs modules^100^. Variant evaluation, including the number of raw or filtered variant counts, the proportion of novel variants not present in dbSNP, and the ratio of Ti/Tv mutations, was conducted via the VariantEval tool. The identified SNPs were categorized into four groups on the basis of their MAF: (i) singleton variants, (ii) rare variants (MAF < 0.01), (iii) intermediate variants (0.01 ≤ MAF < 0.05), and (iv) common variants (MAF ≥ 0.05).

### Dataset mering

The newly generated GRCh37-based dataset was merged with the published HO and 1240K datasets of contemporary and ancient populations worldwide, as included in the Allen Ancient DNA Resource (AADR)^101^. Additionally, genotype profiles of Northwest Chinese populations from the pilot study of 10K_CPGDP were incorporated into this study^102^. Data filtering was performed via PLINK v1.90^103^, excluding variants with missing call rates greater than 0.05 (--geno 0.05), MAFs less than 0.05 (--maf 0.05), and exact test p values for Hardy□Weinberg equilibrium less than 10^−6^ (--hwe 10^−6^). Samples with missing call rates exceeding 0.1 (-- mind 0.1) were also removed. Relatedness coefficients between individuals within populations were estimated via KING v.2.3.0^104^ and PLINK v.1.90^103^. The final merged HO dataset comprised 13,347 individuals and 593,050 SNPs, whereas the merged 1240K dataset included 16,758 individuals and 1,135,264 SNPs. These datasets were primarily utilized for exploring population structure, modeling ancestral admixture, and reconstructing demographic history. Furthermore, the GRCh38-based dataset was merged with 929 high-coverage whole-genome sequences from 54 diverse human populations from the HGDP^105^ and 317 high-coverage genomes from 20 Oceanian populations^19^ to create the WGS dataset. Only biallelic autosomal variants were retained for analyses requiring high-density markers, such as inferring effective population sizes, estimating population divergence times, and detecting signatures of natural selection.

### Population structure and admixture

To investigate the genetic affinity and population stratification of CAAH within Eurasian and regional contexts, we conducted PCAs using the smartpca program in EIGENSOFT v7.2.1^106^, with parameters set to numoutlieriter: 0 and lsqproject: YES. Modern Mlabri and ancient populations were projected onto the first two principal components. Scatter plots were generated using R v4.2.2 and custom scripts. An unsupervised model-based clustering analysis was performed using the maximum likelihood method implemented in ADMIXTURE (v1.3.0) within Eurasian and regional populations^107^. The number of ancestral components (K) ranged from 2 to 20, with a 10-fold cross-validation setting (--cv=10). SNPs in strong LD were pruned using the “--indep-pairwise 200 25 0.4” command in PLINK v1.90^103^. For each value of K, 100 bootstrap resamplings were conducted with different random seeds. Fst distances between population pairs were calculated using PLINK v1.90^103^, excluding populations with sample sizes less than five. The phylogenetic relationships among ethnolinguistically diverse target populations were further estimated using TreeMix v1.13^108^. The frequency spectrum for each population was estimated using PLINK v1.90^103^ and subsequently utilized as input for constructing TreeMix-based topologies. Migration edges, ranging from 0 to 7, were explored using the -m parameter to assess potential gene flow events.

### F-statistical analysis

Various *f_3_*- and *f_4_*-statistics were computed using ADMIXTOOLS v.7.0.2 to measure shared genetic drift between two populations since their divergence from a common ancestral population^109^. Outgroup *f_3_*-statistics of the form *f_3_*(Target, Reference; Mbuti) were performed to estimate overall allele sharing between all target and reference populations. Additionally, admixture *f_3_*-statistics of the form *f_3_*(Ref1, Ref2; Target) were calculated for all pairs of modern or ancient references to test whether the target population was derived from the admixture of populations related to two predefined ancestry surrogates. Tree-like relatedness between four populations was further evaluated using the qpDstat package in ADMIXTOOLS with the *f_4_* model (*f_4_*Mode: YES)^109^. The block jackknife method was employed to assess the statistical significance of the *f_4_*-values. To quantify genetic homogeneity and heterogeneity between two target populations, we conducted symmetrical *f_4_*(Target1, Target2; Ref, Mbuti). Furthermore, asymmetrical estimates of *f_4_* (Ref1, Ref2; Target, Mbuti) and *f_4_*(Ref1, Target; Ref2, Mbuti) were calculated to determine whether the target populations derived more ancestry from reference populations related to Ref1 or Ref2.

### Admixture modeling with qpWave and qpAdm

To determine whether target population pairs (left populations) formed a single clade relative to a set of outgroups (right populations), we conducted rank tests using the qpWave program (version 1520)^109^. Admixture modeling based on the *f_4_*-statistics was performed using the qpAdm program (version 1520) within the ADMIXTOOLS package^109^. This approach assessed the number of distinct ancestry streams from surrogate populations needed to explain the genetic makeup of the target populations and estimated the corresponding admixture proportions. The parameters “allsnps: YES” and “details: YES” were applied to include all SNPs across the four populations and display the specific entries of the *f_4_* matrix and Z scores.

### Admixture time estimation

Long-distance migrations and subsequent admixtures between populations can lead to an exponential decay of admixture-induced LD. To compute the weighted LD decays and infer the admixture dates, we utilized MALDER v1.0 with additional parameters set to mindis: 0.005 and jackknife: YES^110^. Multiple modern populations from East and West Eurasia were employed as potential ancestral sources, testing all possible combinations.

### Haplogroup classification and clustering analysis

Y-chromosomal haplogroups were classified using Y-LineageTracker 1.3.0^111^ and the hGrpr2.py package within HaploGrouper^112^. For the HaploGrouper-based analysis, two reference files, treeFileNEW_isogg2019.txt, and snpFile_b38_isogg2019.txt, were utilized. Haplogroup frequencies of the targeted terminal lineages were estimated at various levels. Maternal haplogroups were classified using HaploGrep2^113^ and HaploGrouper, based on PhyloTree17.

### Inference of sex-biased admixture

To evaluate potential sex-biased admixture processes in CAAH, ADMIXTURE was used to estimate ancestry proportions of surrogate source populations on both autosomes and X chromosomes. Z scores were calculated to assess differences between autosome- and X chromosome-based admixture proportions for a given ancestry, using the formula 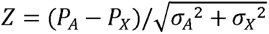, where P_A_ and P_X_ represent the ancestry proportions on autosomes and X chromosomes, respectively, and σ_A_ and σ_X_ are their corresponding standard errors^114^. A positive Z score indicates a higher ancestry proportion on the autosomes than on the X chromosomes, suggesting a male-driven admixture; conversely, a negative Z score indicates a female-driven admixture. Due to the original use of this formula for assessing differences in qpAdm-based ancestry proportions and the unavailability of X chromosome genotype data for source populations modeled with qpAdm in this study, ADMIXTURE was employed to estimate admixture proportions based on the WGS dataset. Additionally, a Student’s t-test was performed^92^, with a P value less than 0.05, indicating the occurrence of sex-biased admixture in the target population.

### Haplotype-based fine-scale population structure reconstruction

#### Segmented haplotype estimation and fine-scale population structure identification

Stricter filtering criteria of geno: 0.1 and mind: 0.1 were applied to prune genome-wide SNPs. The Segmented HAPlotype Estimation & Imputation Tool (SHAPEIT v2.r904) was then used to estimate haplotypes based on the genetic maps from HapMap phase II b37^115,116^. To identify an optimal starting point for the estimated haplotypes and generate more parsimonious graphs, we performed haplotype phasing with ten burn-in iterations (--burn 10), ten pruning iterations (--prune 10), and 30 main iterations (-- main 30) of MCMC (Markov chain Monte Carlo). The fine-scale population structure was subsequently analyzed using fineSTRUCTURE v4.0 83 based on the phased haplotypes^117^.

#### Shared IBD fragment estimation

Refined IBD (refined-ibd.17Jan20.102.jar) was used to detect shared IBD segments between each pair of individuals^116^. A minimum length of 0.1 cM was set for the identified IBD fragments using the parameter length=0.1. The detected IBD segments were classified into three categories: 1–5 cM, 5–10 cM, and >10 cM. Segments in the 1–5 cM range likely reflect ancient genetic connections dating back 1,500–2,500 years, those in the 5–10 cM range correspond to a time interval of approximately 500–1,500 years ago, and segments greater than 10 cM likely indicate recent shared ancestry within the last 500 years. To provide an overview of IBD sharing, we summed the total count and length of IBD segments for each pair of individuals, and the population mean for each block was calculated.

#### Admixture events inferred from fastGLOBETROTTER

ChromoPainterv2 was used to paint each chromosome of the recipient populations as a series of haplotype chunks and to estimate the number of ancestry chunks inherited from donor populations^117^. Initially, ChromoPainterv2 was run to paint each phased haploid of the first ten recipient individuals, using ten iterations of expectation-maximization with the ‘‘-in’’ switch and ‘‘-iM’’ emission parameters to estimate the chunk size and switch error rate. A Perl script was subsequently applied to estimate the switch (-n) and emission (-M) rates across individuals. The estimated parameters were then used to rerun ChromoPainterv2 to construct the copy vectors, modeling both recipient and donor individuals as a patchwork of donor haplotypes. The ‘‘chunk length’’ output files obtained across all chromosomes were summed. FastGLOBETROTTER was then employed to identify, date, and describe admixture events in target populations, using the chunk length output and painting samples, with the options ‘‘prop.ind: 1’’ and ‘‘null.ind: 1’’^118^. The significance of the inferred admixture dates was estimated using ‘‘bootstrap.date.ind: 1’’ and ‘‘bootstrap.num: 100’’ options. SourcefindV2 was also used to infer admixture models based on haplotype sharing identified by ChromoPainter.

### Demographic history inference

For each newly sequenced population, SMC++ was used to infer the population size history^119^. Input files were prepared on the basis of the genotypes of all called biallelic SNPs and included all individuals from each population. The two samples with the highest sequencing coverage in each population were specified as the distinguished lineages. SMC++ was run with a mutation rate of 1.25 × 10^−8^ per base pair per generation and a constant generation time of 29 years. To infer the population divergence time, MSMC2 (multiple sequentially Markovian coalescent) was employed^120^. The two samples with the highest sequencing coverage were selected from each target population for MSMC2 analysis. The French and Han Chinese populations from the HGDP were chosen as representatives of European and East Asian ancestries, respectively. MSMC2 analyses were conducted following standard recommendations^119^, utilizing mappability masks for the human reference genome GRCh38 generated by the SNPable pipeline. MSMC2 was run three times independently to estimate coalescence rates within population1/population2 and across populations. Absolute time estimates were calculated using the same mutation rate and generation time as those adopted in SMC++. The relative cross coalescence rate (rCCR) was calculated, and the divergence time for each population pair was inferred at the point where the rCCR reached 0.5.

### Medical relevance and biological adaptation

#### Variant annotation

Variant annotation of the GRCh37-based dataset was performed using Ensembl VEP, with a focus on variants annotated as pathogenic or likely pathogenic^121^. These pathogenic variants were reviewed against the ACMG-SF v3.2 gene panel following the criteria outlined by the ACMG^122^. SnpEff v4.3, with default parameters, was employed for LoF predictions^123^, defining a variant as a PTV if it was annotated as “frameshift_variant,” “transcript_ablation,” “splice_acceptor_variant,” “splice_donor_variant,” or “stop_gained.” To generate a list of high-confidence PTVs, we restricted variants to high-confidence regions as determined by Genome in a Bottle (ftp://ftp-trace.ncbi.nlm.nih.gov/ReferenceSamples/giab/data/AshkenazimTrio/analysis/NIST_v4.1_SmallVariantDraftBenchmark_12182019/GRCh37/HG002_GRCh37_1_22_v4.1_draft_benchmark.bed).

#### Estimation of AF deviations and HDVs

To screen for signals of natural selection, CAAH was divided into four groups based on haplotype-based clustering patterns and results from admixture modeling. CAAH1 primarily encompassed populations such as TJK, KLKTL, TKM, and UZTK, which exhibited a greater proportion of West Eurasian ancestry. CAAH2 mainly included UGAMT, HZAF, and KRKP, showing a balanced mixture of West and East Eurasian ancestries. CAAH3 consisted of KGZ, characterized by a relatively higher proportion of East Eurasian ancestry, while CAAH4 predominantly comprised DGJB, which displayed a strong genetic affinity with East Asian populations. The AFd_e_ of genome-wide variations within CAAH groups was estimated based on the absolute difference between AF_obs_ and AF_exp_. The AF_exp_ was calculated as the admixture proportion weighted by the frequencies in ancestral populations. Weights were assigned based on global admixture proportions estimated through ADMIXTURE analysis. A window-based approach, as described in previous studies^40^, was employed to identify variants exhibiting significant AFd_e_, which serve as genomic signatures of local adaptation.

Signals of natural selection were further identified by screening for HDVs between East and West Eurasians with significant AF deviations. CAAH was treated as a simple composite of east–west Eurasian ancestral populations to facilitate HDV identification. The Han Chinese and French populations from the HGDP were utilized as representative ancestral populations of East and West Eurasia, respectively. Pairwise Fst values of overlapping SNPs between these ancestral populations were calculated to identify HDVs, with SNPs having Fst values greater than 0.2 classified as HDVs. The AF deviation was measured by calculating AFd_e_ (|AF_obs_ - AF_exp_|), where AF_exp_ = *f*_East_×□ + *f*_West_×(1-□)^41,42^. Here, *f*_East_ and *f*_West_ represent the AFs of a specific SNP in the East and West Eurasian ancestral populations, respectively, while □ denotes the admixture proportion of East Eurasian ancestry in CAAH, and (1-□) represents the proportion of West Eurasian ancestry. HDVs exhibiting an AFd_e_ greater than 0.1 in CAAH groups suggest potential adaptive significance. Functional annotations of candidate variants were performed using the GWAS Catalog (v.2022-09-14), and pathway enrichment was assessed with Metascape^124^. The frequency trajectory of the detected natural selection signal was visualized via the AGES browser (https://reich-ages.rc.hms.harvard.edu/#/). To identify evidence of natural selection, X-statistic values derived from ancient DNA time series data^54^ were applied.

### Archaic introgression estimation

The IBDmix (v.1.0.1) method was initially employed to detect AISs likely derived from Neanderthals^77^. Owing to the limited sensitivity of IBDmix in detecting Denisovan-derived AISs, Sprime v.07Dec18.5e2 was applied exclusively to identify AISs in CAAH that are likely derived from Denisovans^125^. Additionally, Sprime was utilized to detect Neanderthal-derived AISs. The newly generated data were merged with the 1KGP WGS data for IBDmix analysis. All multi-allelic variants and InDels from the archaic genome and the merged dataset were removed, with a focus on autosomal variants. Archaic ancestry detection was then conducted within each CAAH subgroup to mitigate the impact of population structure. To minimize misclassification due to incomplete lineage sorting, only AISs with a logarithm of the odds ratio for the linkage score above 4 and a length greater than 50 kb were included in downstream analyses. To maximize the detection of Neanderthal-like sequences, a conservative approach was used for filtering the callset. After the initial identification of Neanderthal- and Denisovan-derived sequences via IBDmix, the Neanderthal-related callset was refined by masking regions identified as Denisovan-like segments in Africans that were also classified as Neanderthal-derived sequences in any population.

For the Sprime analysis, the four previously defined CAAH groups were designated as target populations, with the YRI population (Yoruba in Ibadan, Nigeria, n = 108) serving as the outgroup. A score threshold of 150,000 and default parameters were employed to optimize performance and accuracy. To ascertain the origin of AISs and the proportion of putative archaic alleles matching reference sequences, we calculated the match rate for each AIS, which was defined as the ratio of matched sites to the total number of compared sites. A contour plot was generated via the kde2d function from the MASS package in R to visualize distinct waves of archaic introgression. Segments with clear Neanderthal or Denisovan origins were extracted for further exploration of the biological functions of the identified AISs. Only segments with at least 30 putatively introgressed alleles comparable to the Vindija Neanderthal genome or at least 30 comparable to the Denisovan genome were considered. Neanderthal-derived segments were defined as those with a match rate exceeding 0.6 with the Vindija Neanderthal genome and below 0.3 with the Altai Denisovan genome. Conversely, Denisovan-derived segments were characterized by a match rate above 0.3 with the Altai Denisovan genome and below 0.3 with the Vindija Neanderthal genome. To minimize misclassification due to incomplete lineage sorting, only AISs with a logarithm of the odds ratio for the linkage score above 4 and a length greater than 50 kb were included in downstream analyses^126^. Finally, the Neanderthal-inherited AISs identified via both methods were intersected via bedtools to generate a high-confidence dataset for adaptive introgression signal identification^127^. To obtain high-confidence signals of adaptive introgression, we also employed the haplotype-based integrated haplotype score (iHS) to examine the genomic signatures of natural selection. Estimates of iHS scores were conducted via Selscan v1.2.0^128^, with normalization performed via norm v1.2.1 with the parameters “--bp-win --winsize 100000 --qbins 10”. SNPs with normalized absolute iHS values exceeding two were selected as candidates for selective sweeps. The focus was narrowed to archaic variants identified in both frequency- and haplotype-based natural selection signals.

## Compliance and ethics

### Ethics approval and consent to participate

The Medical Ethics Committee of the National Centre for Biotechnology of Kazakhstan (No. 2 of 10 June 2020) and Nazarbayev University Institutional Research Ethics Committee (No. 17 of 16 April 2019) approved this study. This study was conducted following the principles of the Helsinki Declaration of 2013.

### Consent for publication

Not applicable.

### Data availability

All the data used here are included in the supplementary materials. Reference populations can be found in publicly available datasets, such as the Human Genetic Diversity Project dataset, Oceania genomic resource, and AADR (HO dataset and 1240K dataset) from the David Reich Lab (https://reich.hms.harvard.edu/datasets). The raw data of 166 individuals were submitted to the Genome Variation Map database under accession number GVM000900 (https://bigd.big.ac.cn/gvm/getProjectDetail?Project=GVM000900). The acquisition and use of these data complied with the regulations of the People’s Republic of China on the administration of human genetic resources. The requests for access to raw data should be directed to Guanglin He.

### Funding

This work was supported by the National Natural Science Foundation of China (82402203 and 82202078), the Science Committee of the Ministry of Education and Science of the Republic of Kazakhstan (Grant Nos. BR18574101 and AP19677968), the Collaborative Research Program of Nazarbayev University (Grant No. 091019CRP2119), the Faculty-development competitive research grants programs of Nazarbayev University (Grant No. SST2019012), the National Natural Science Foundation of Chongqing (CSTB2024NSCQ-LZX0005), the Major Project of the National Social Science Foundation of China (23&ZD203), the Open Project of the Key Laboratory of Forensic Genetics of the Ministry of Public Security (2022FGKFKT05 and 2024FGKFKT02), the Center for Archaeological Science of Sichuan University (23SASA01 and 24SASB03), the 1□3□5 Project for Disciplines of Excellence, West China Hospital, Sichuan University (ZYJC20002), and the Sichuan Science and Technology Program (2024NSFSC1518).

### Authors’ contributions

G.H., M.W., M.Z., and L.W. conceived and supervised the project. M.Z. and Z.S. collected the samples. G.H., Q.S., H.S., and M.W. extracted the genomic DNA and coordinated the genome sequencing. G.H., Q.S., H.S., and M.W. performed variant calling. Q.Y., L.L., G.H., J.Z., H.S., and M.W. performed the population genetic analysis. M.W. analyzed the Y-chromosome and mtDNA haplotype data. M.W. and G.H. drafted the manuscript. R.T., H.Y., Y.L., F.B., J.C., G.H., M.W., M.Z., and C.L. revised the manuscript.

## Acknowledgments

We thank Prof. Etienne Patin and Prof. Lluis Quintana-Murci from the Human Evolutionary Genetics Unit of the Institut Pasteur for sharing the high-coverage genomes of 317 individuals from the Pacific region. We thank Prof. Yerlan Ramankulov from Nazarbayev University for his support and consultation.

