## Supplementary material for "Whole-Genome Sequencing Pilot Study of the Central Asian Genetic Diversity Project Reveals Distinct Genetic Histories, Adaptive Processes, and Introgression Events": Figure S

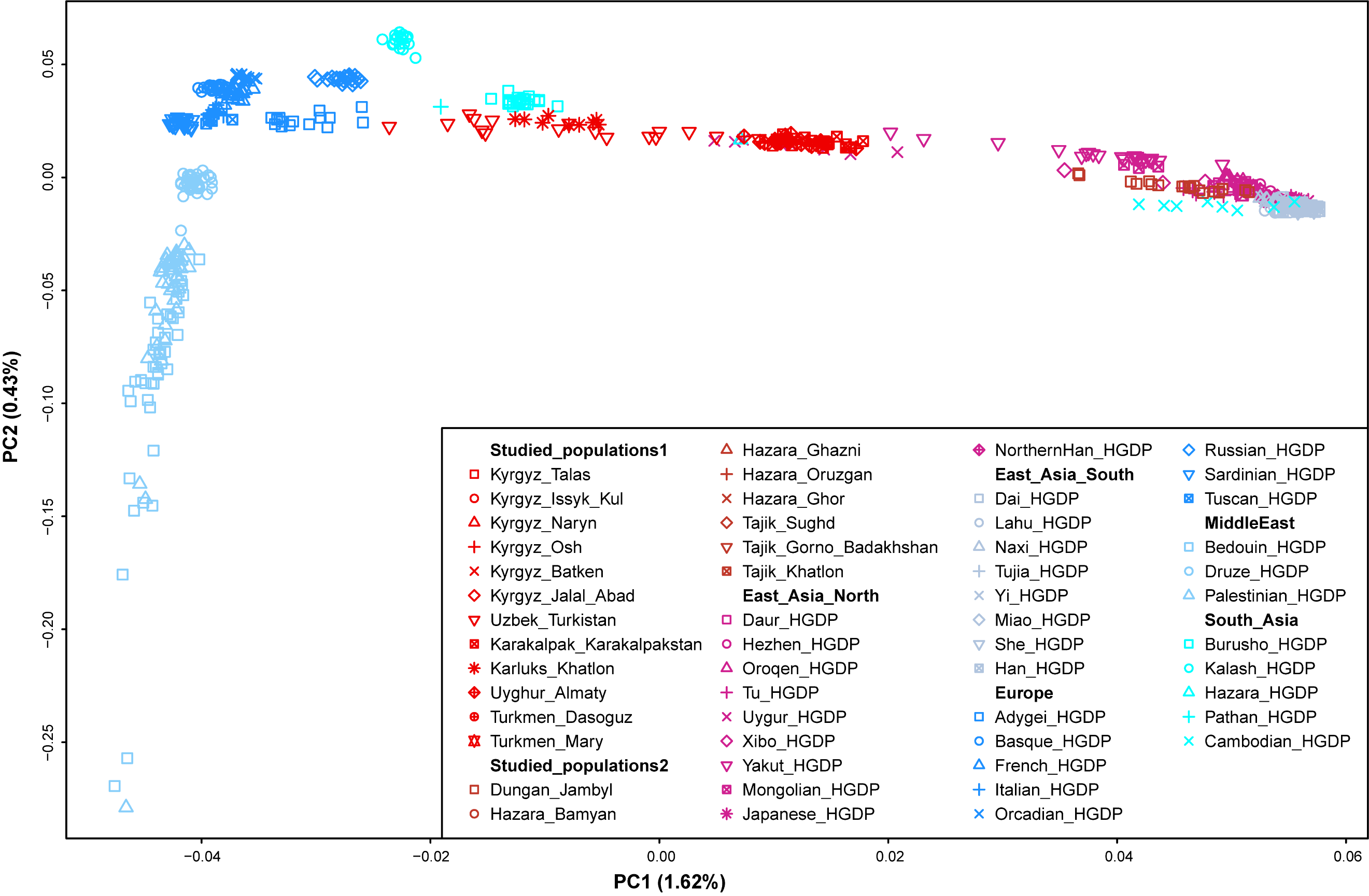

**Figure S1. Genetic relationships between newly sequenced Central Asians and Afghanistan Hazaras (CAAH) and Eurasian reference populations revealed based on the whole-genome sequencing (WGS) dataset.**

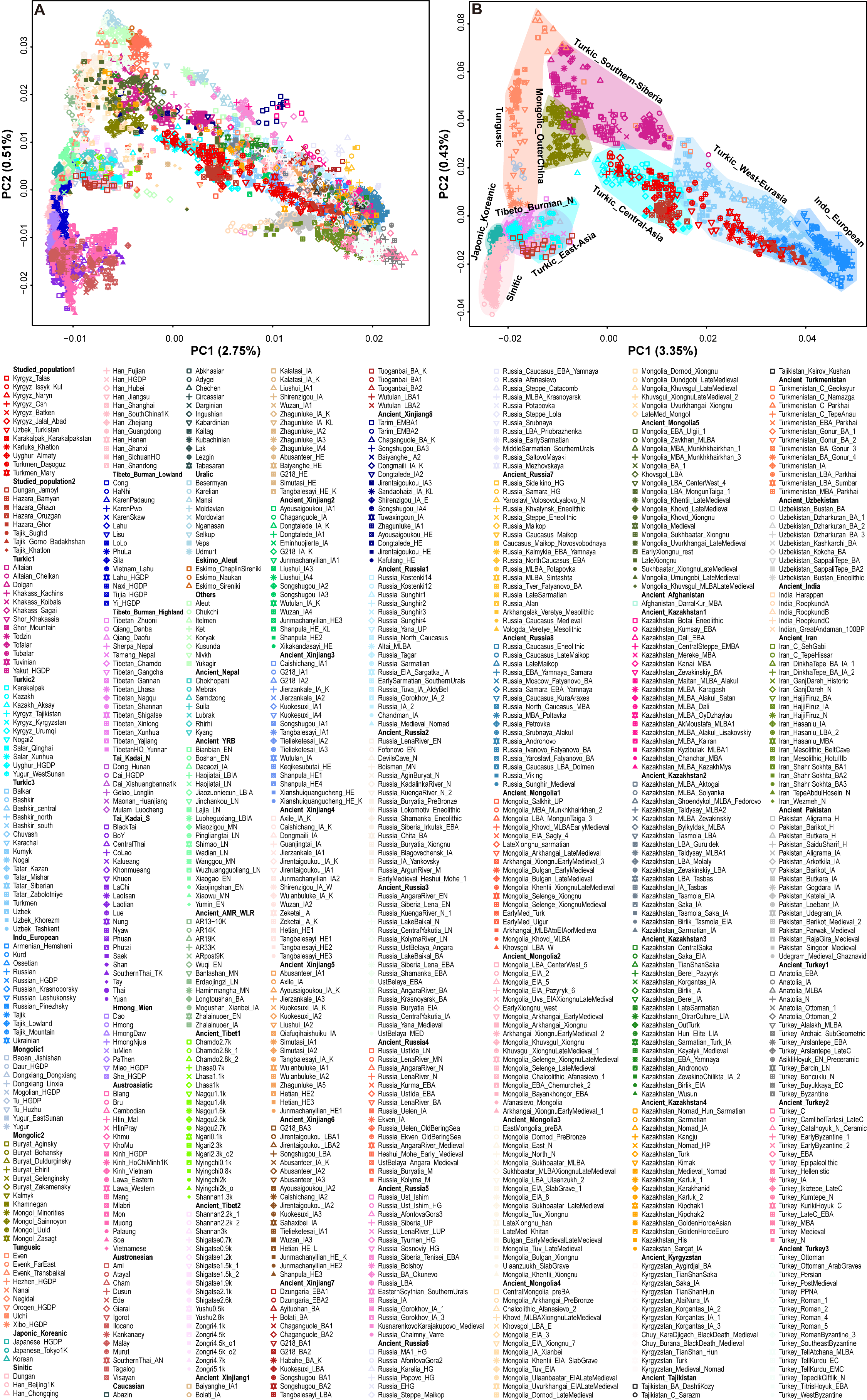

**Figure S2. Genetic relationships between newly sequenced CAAH and spatiotemporally diverse Eurasian reference populations revealed based on the merged Human Origins (HO) dataset.** (**A**), The population structure of 897 modern and ancient Eurasian populations included in the merged HO dataset, in which ancient populations were projected onto the first two PCs. Modern populations were color-coded according to language classifications and ancient populations were color-coded according to geographical locations. (**B**), Patterns of genetic affinity between CAAH and 117 linguistically close (Altaic, Indo-European, and Sino-Tibetan) Eurasian populations. The genetic clusters or clines are labeled.

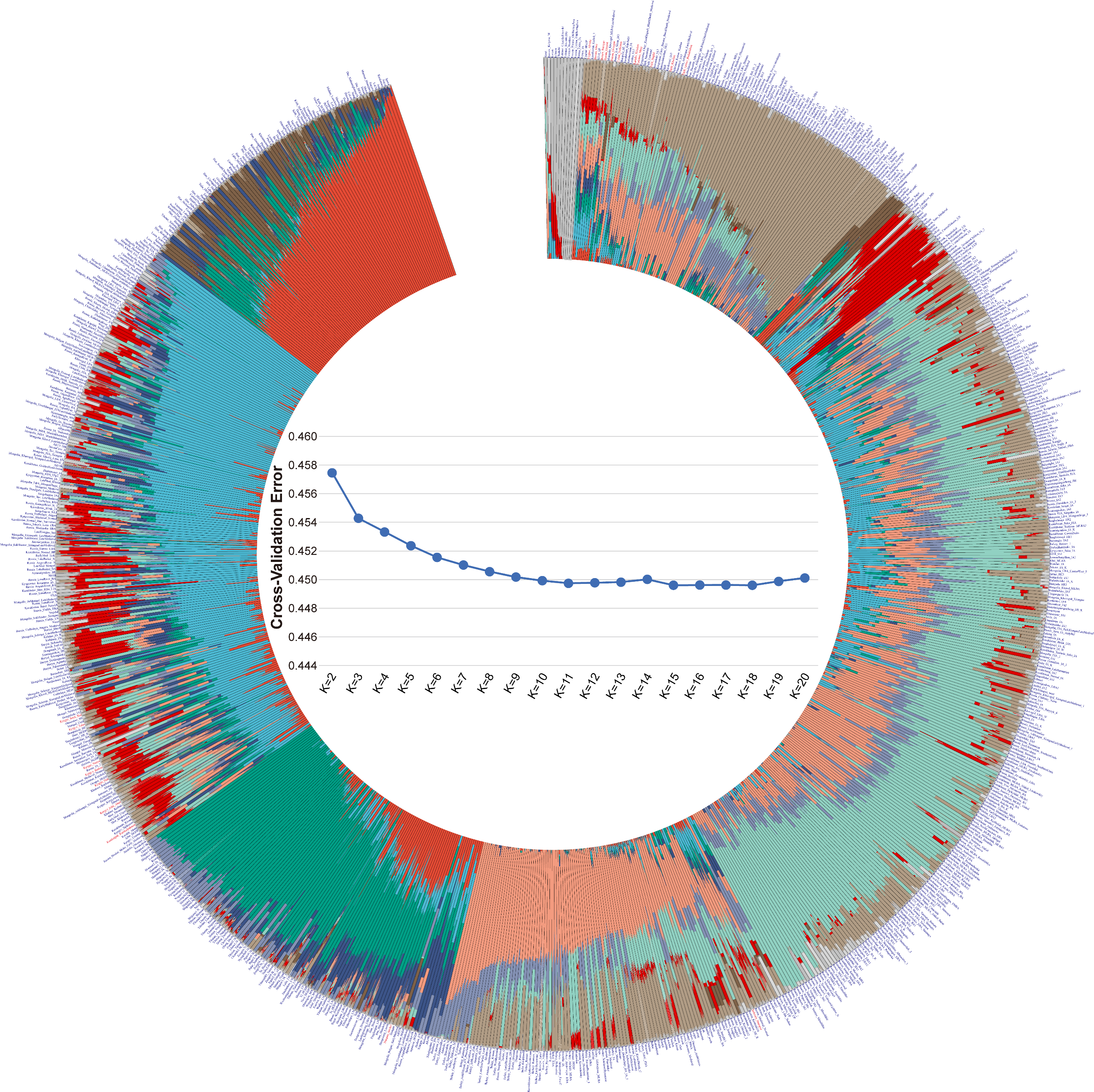

**Figure S3. Model-based ADMIXTURE results of 897 spatiotemporally diverse modern and ancient Eurasian populations included in the merged HO dataset at K=11.** The cross-validation error values are shown in the middle of the graphical Admixture results.

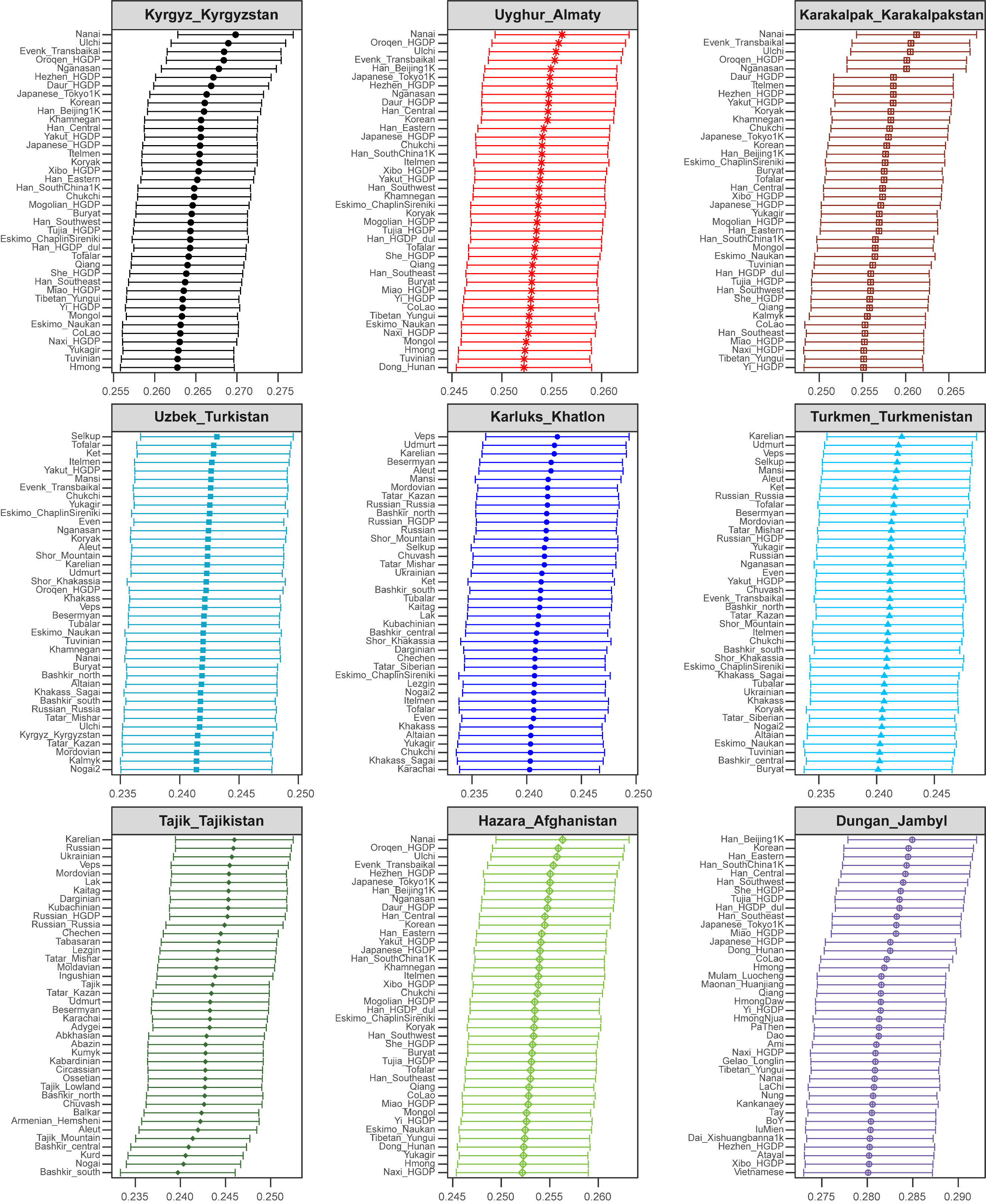

**Figure S4. The shared drift between CAAH and modern Eurasian reference populations estimated using outgroup *f_3_*-statistics of the form *f_3_*(Target, Modern Eurasians; Mbuti).** The top 40 *f_3_*-values of each population are presented.

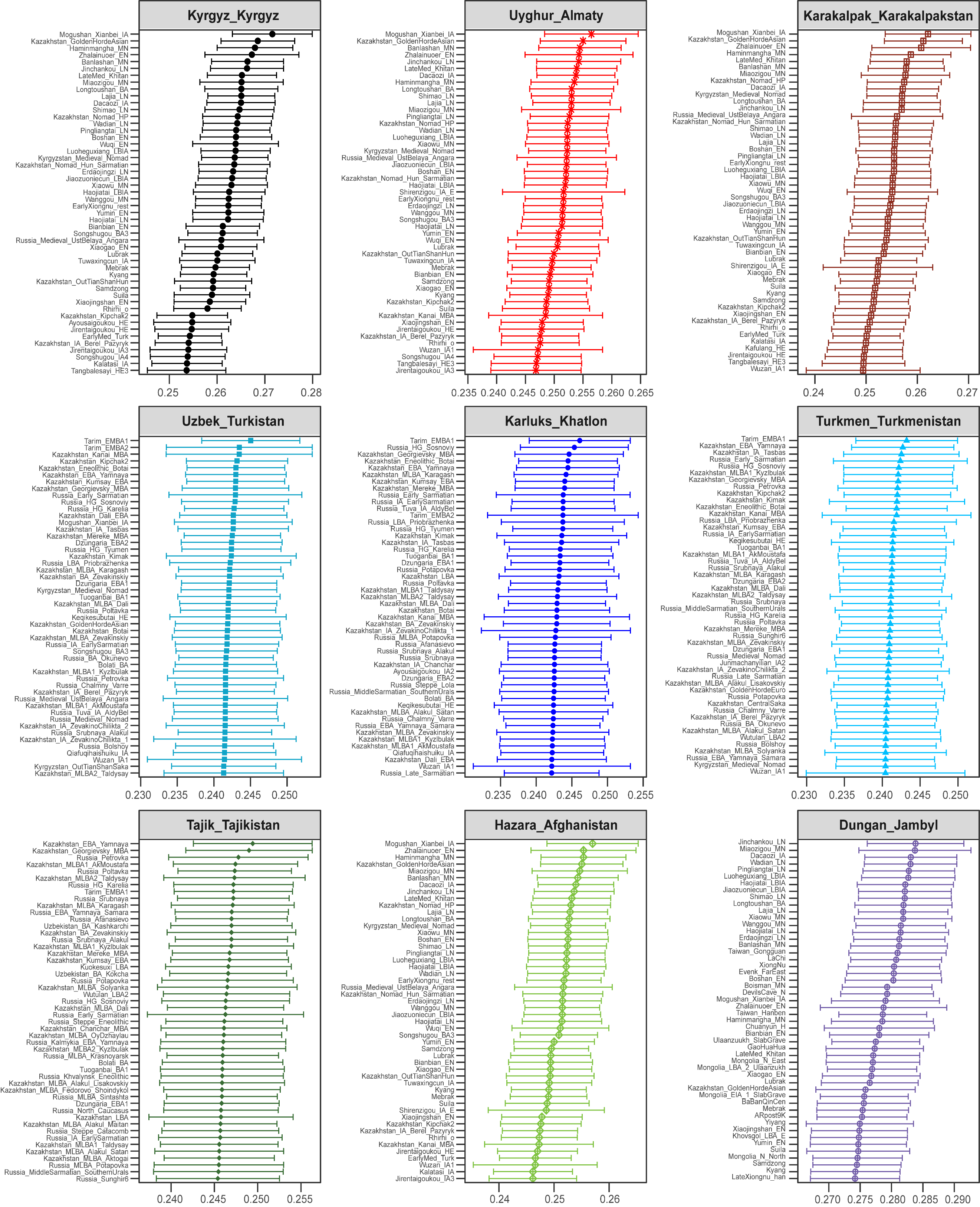

**Figure S5. The shared drift between CAAH and ancient Eurasian reference populations estimated using outgroup *f_3_*-statistics of the form *f_3_*(Target, Ancient** **Eurasians; Mbuti).** The top 40 *f_3_*-values of each population are presented.

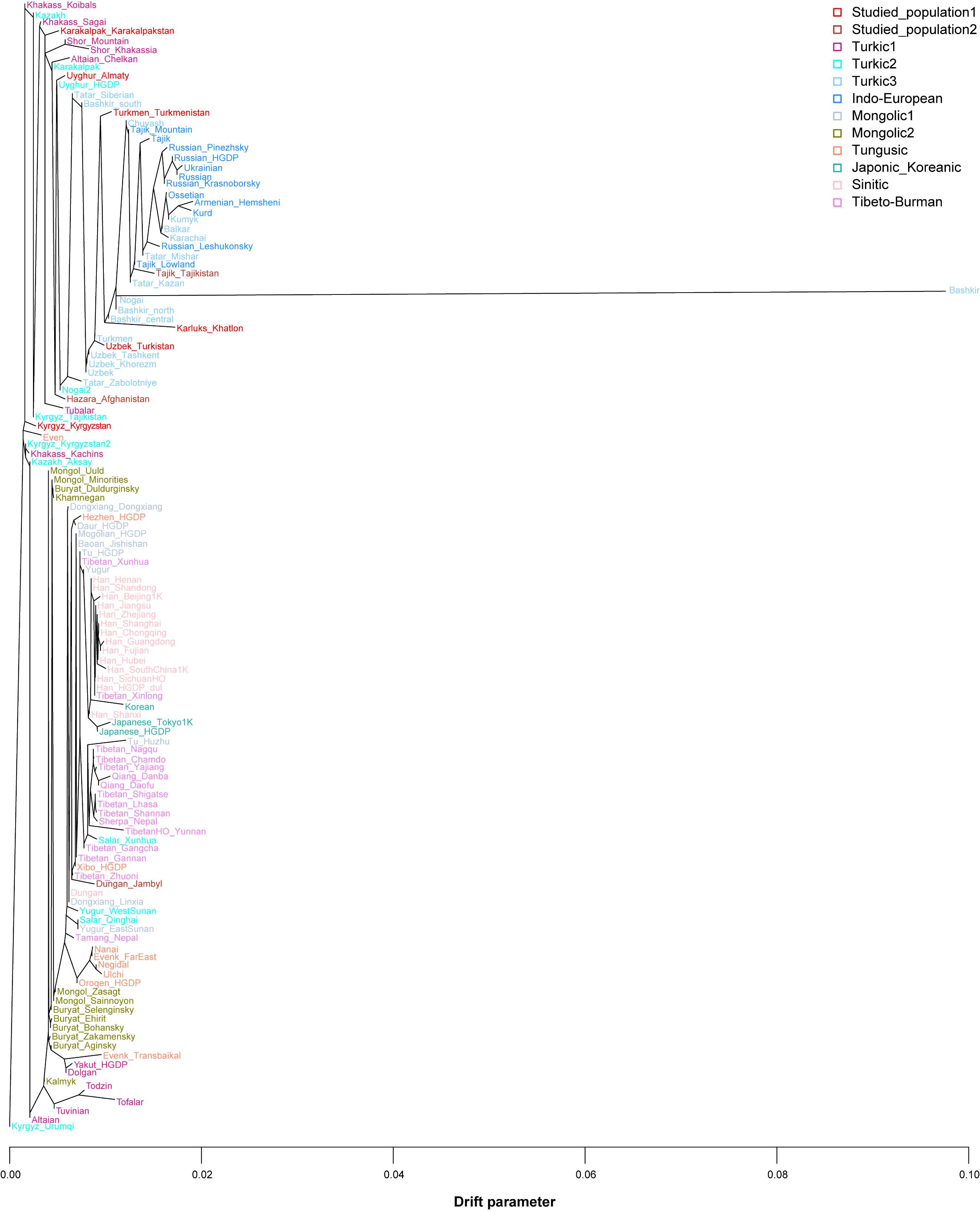

**Figure S6. The Treemix-based topology of CAAH and linguistically close but geographically distinct Altaic and Sino-Tibetan-speaking populations.** The small sample size resulted in Bashkir having an unusually long branch length. The color of the font is consistent with the legend in principal component analysis (PCA).

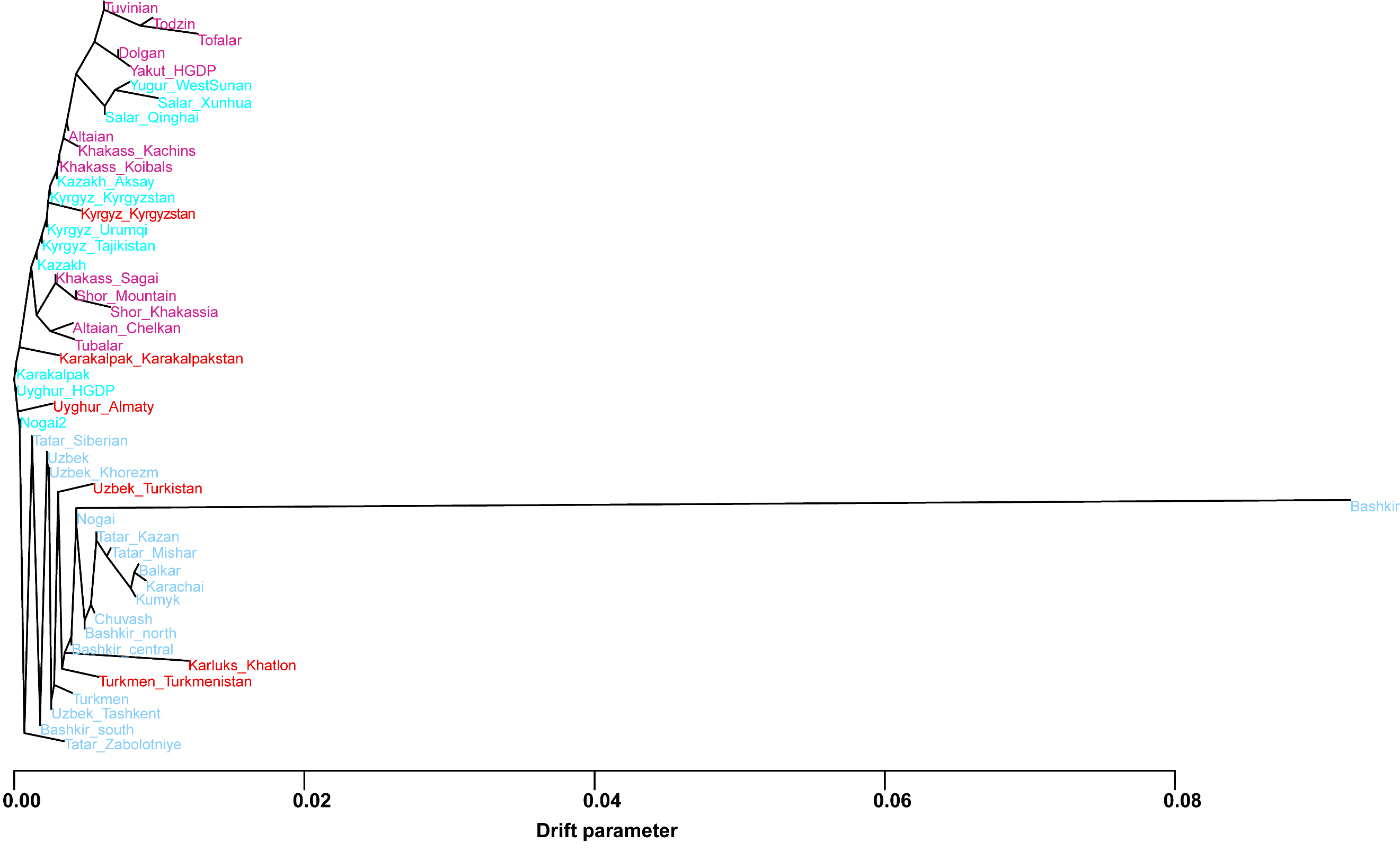

**Figure S7. The Treemix-based topology of 47 geographically distinct Turkic-speaking populations.** The small sample size resulted in Bashkir having an unusually long branch length. The color of the font is consistent with the legend in PCA.

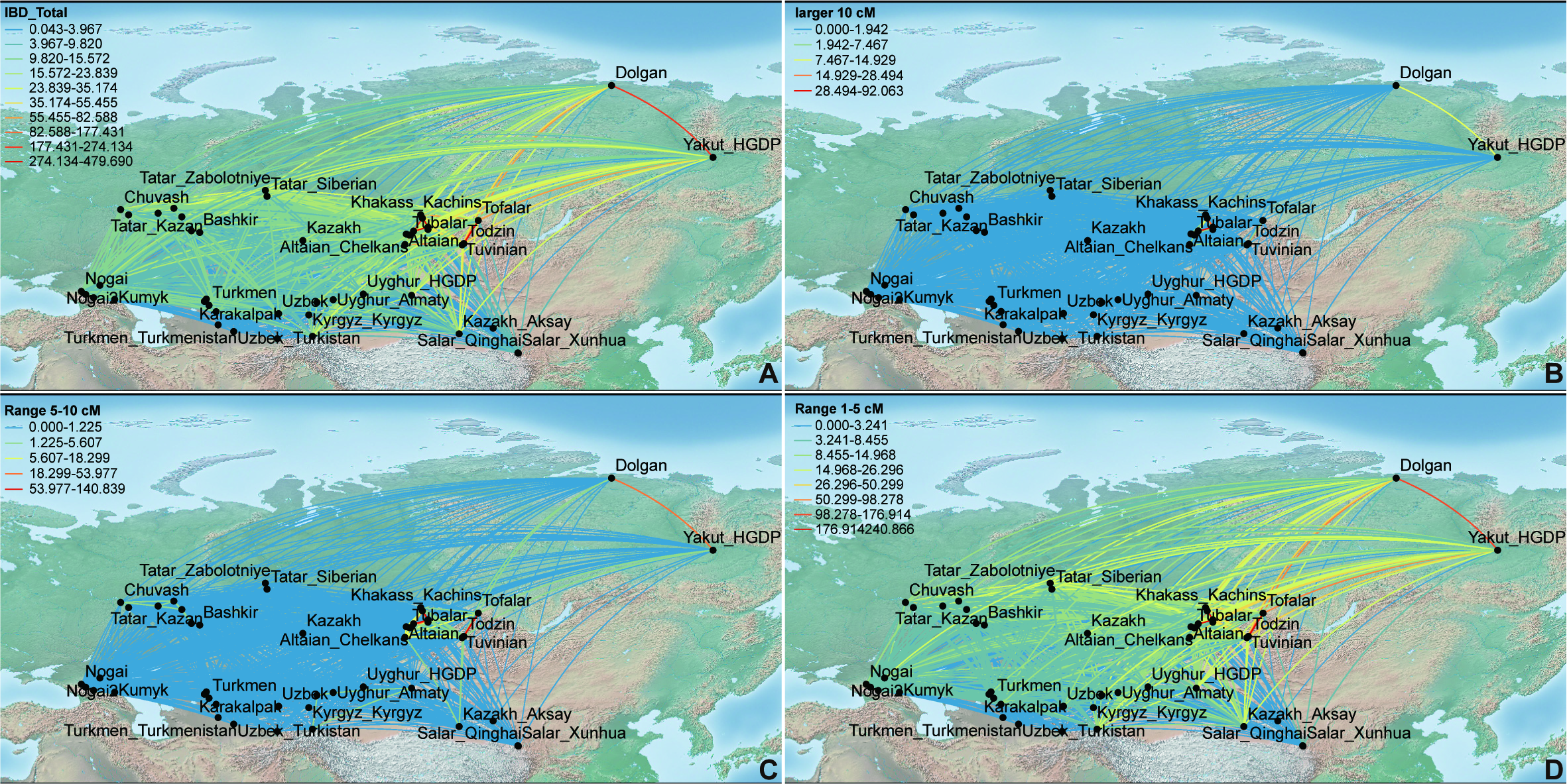

**Figure S8. The** **identity-by-descent (IBD) sharing between all involved Turkic-speaking populations.** (**A**), The total length of shared IBD for each pair of Turkic people included in the merged HO dataset. (**B‒D**), The summed length of identified IBD blocks over 10 cM (youngest), in the range of 5–10 cM or 1–5 cM (oldest) for each pair of Turkic people.

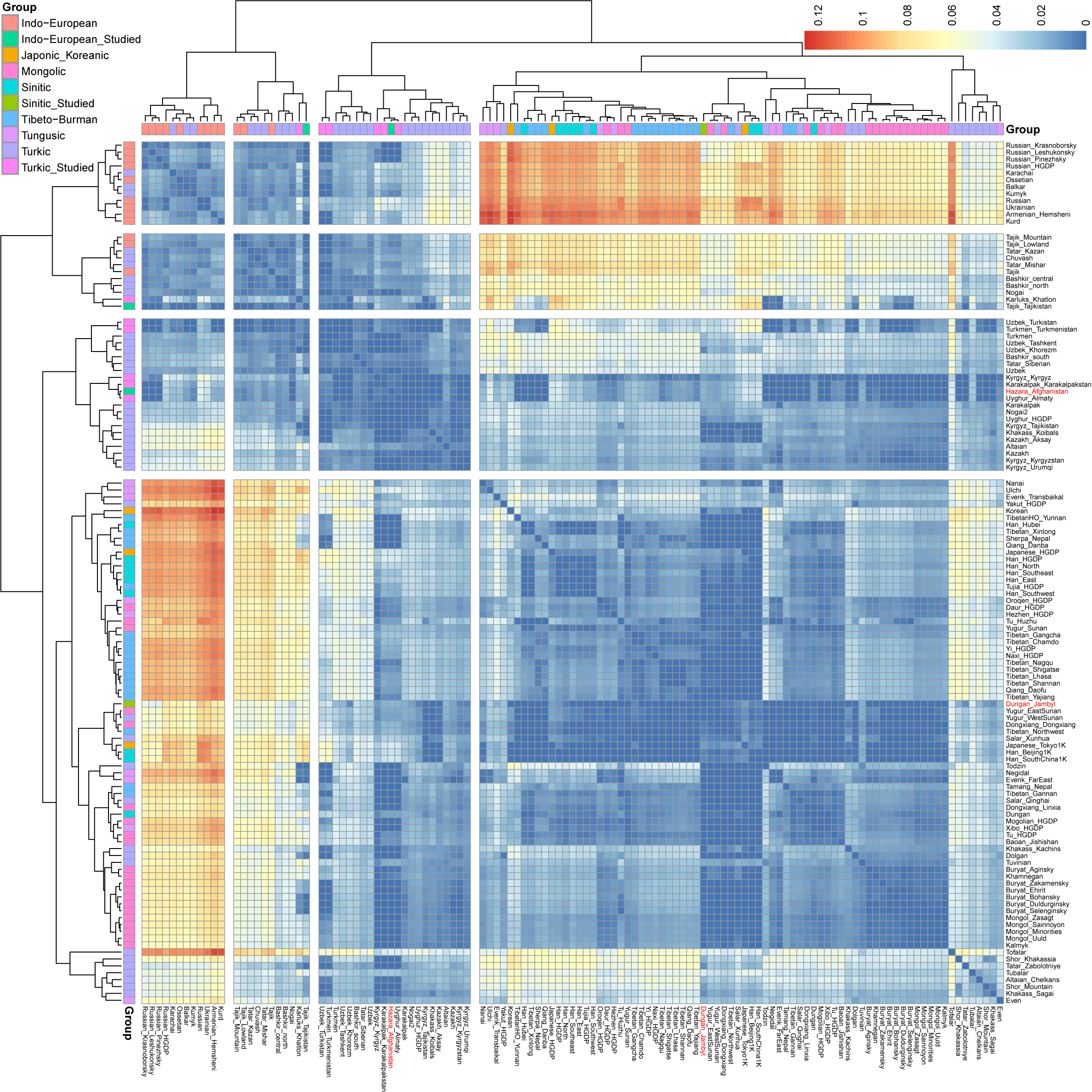

**Figure S9. Pairwise Fst genetic distances between geographically distinct Eurasian populations.**

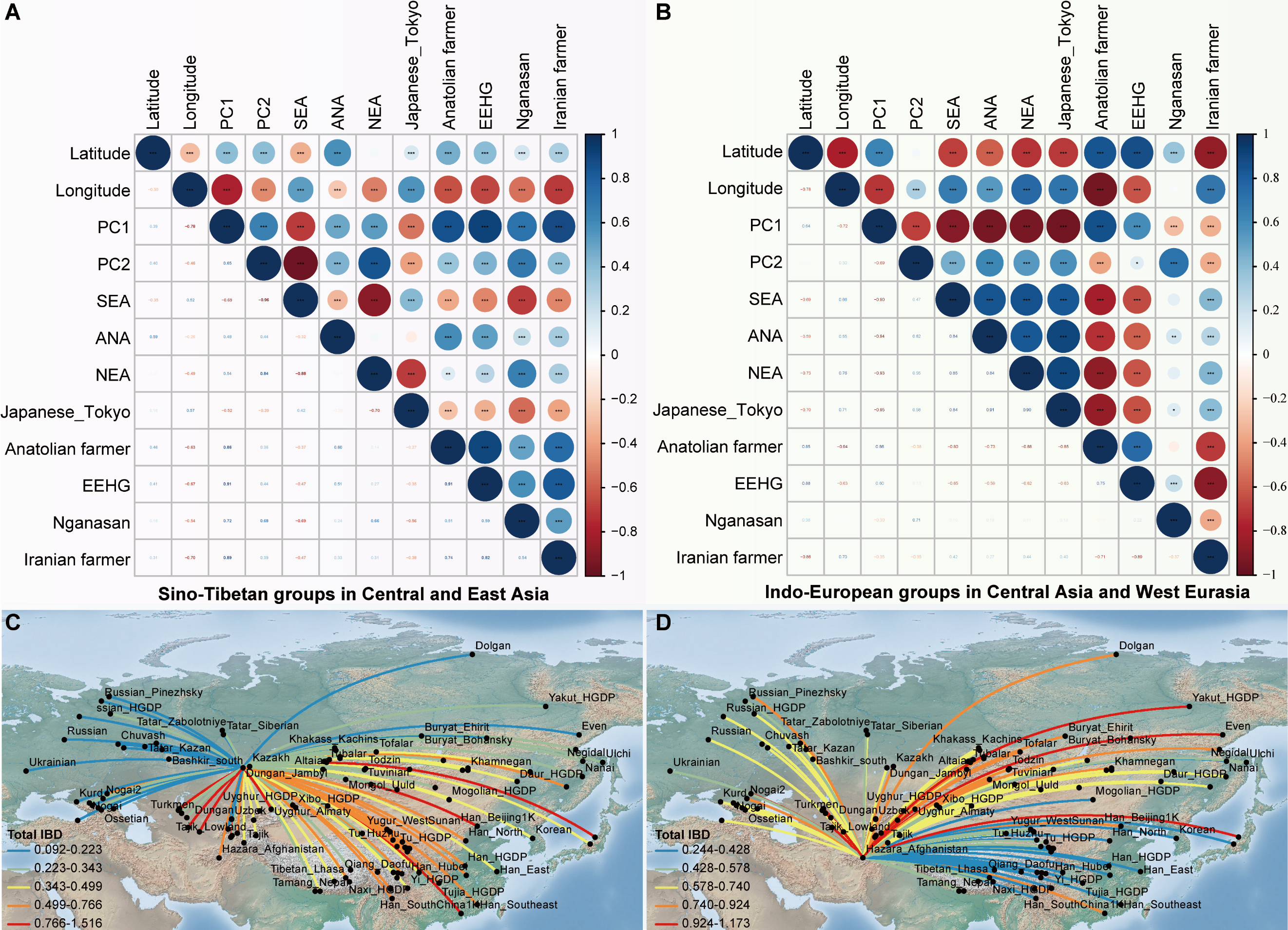

**Figure S10. The correlation between geographical and genetic patterns and IBD sharing between Dungan_Jambyl (DGJB)/Hazara_Afghanistan (HZAF) and Eurasian reference populations**. The correlation between latitude/longitude and genetic features (PC1, PC2, and estimated ancestral components in Fig. S3) and between estimated ancestral components among Sino-Tibetan groups in Central and East Asia (**A**) and among Indo-European-speaking populations in Central Asia and West Eurasia (**B**). SEA: southern East Asian ancestry; ANA: Ancient Northeast Asian ancestry; NEA: northern East Asian ancestry; EEHG: Eastern European hunter-gatherers. (**C**), The total length of shared IBD between DGJB and Eurasian reference populations. (**D**), The total length of shared IBD between HZAF and Eurasian reference populations.

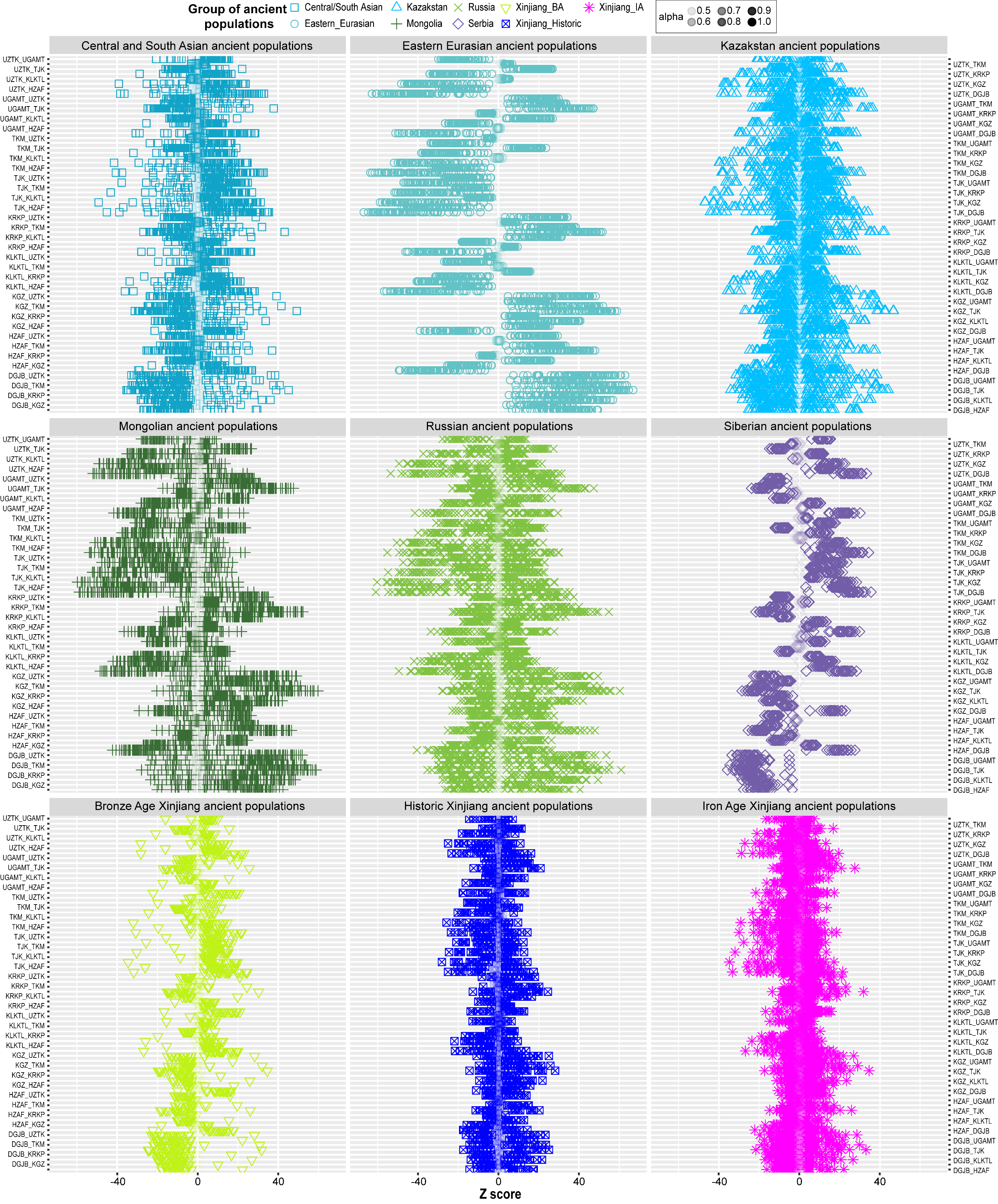

**Figure S11. The Z scores of *f*_4_-statistics of the form *f*_4_(CAAH_1_, CAAH_2_; Reference, Mbuti).** Ancient Eurasian groups were used as reference populations. Different colored shapes represent various groups of ancient reference populations. UZTK: Uzbek_Turkistan, UGAMT: Uyghur_Almaty, TJK: Tajik_Tajikistan, KLKTL: Karluks_Khatlon, TKM: Turkmen_Turkmenistan, KRKP: Karakalpak_Karakalpakstan, and KGZ: Kyrgyz_Kyrgyzstan.

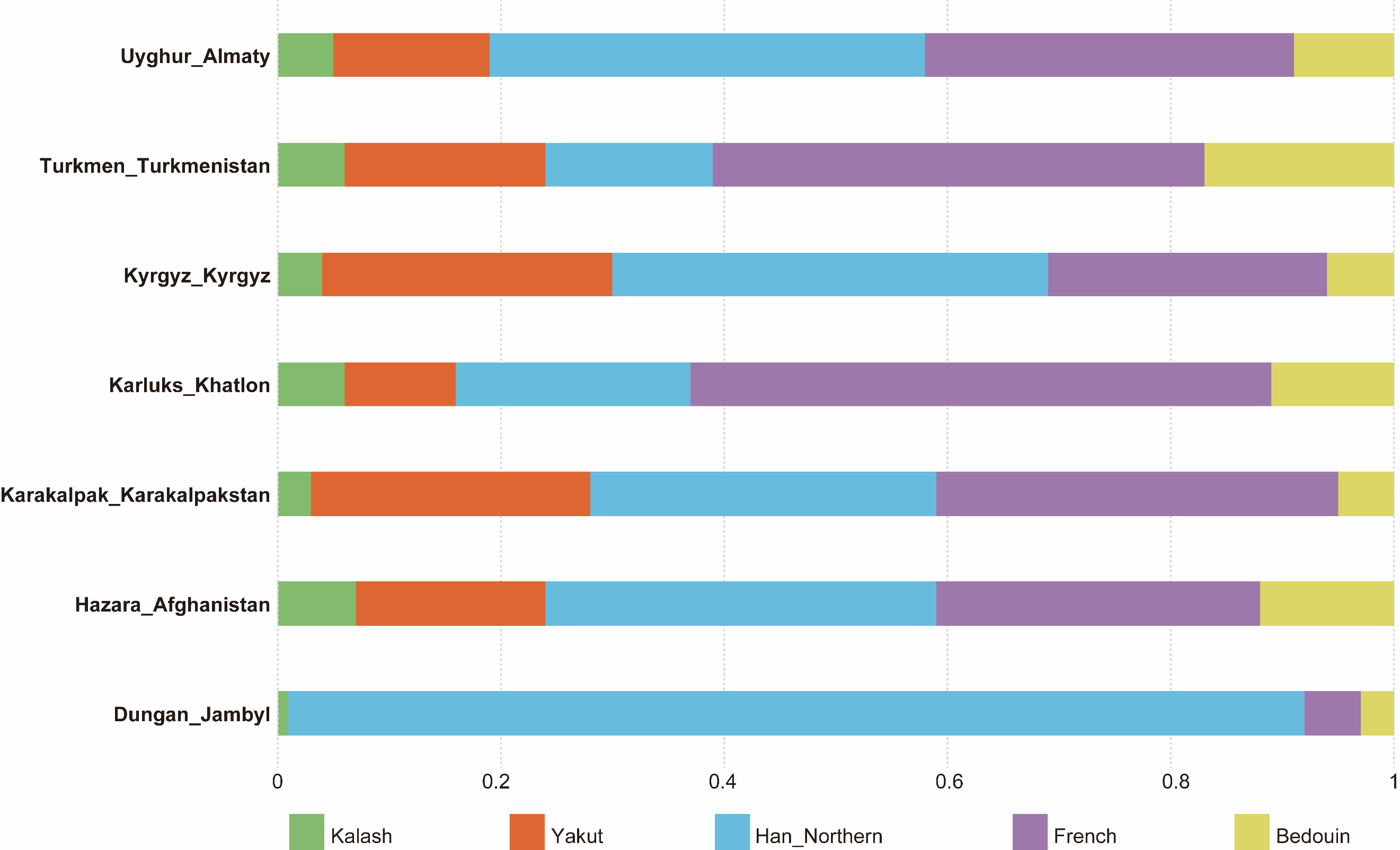

**Figure S12. The admixture profiles of CAAH estimated based on SOURCEFIND.**

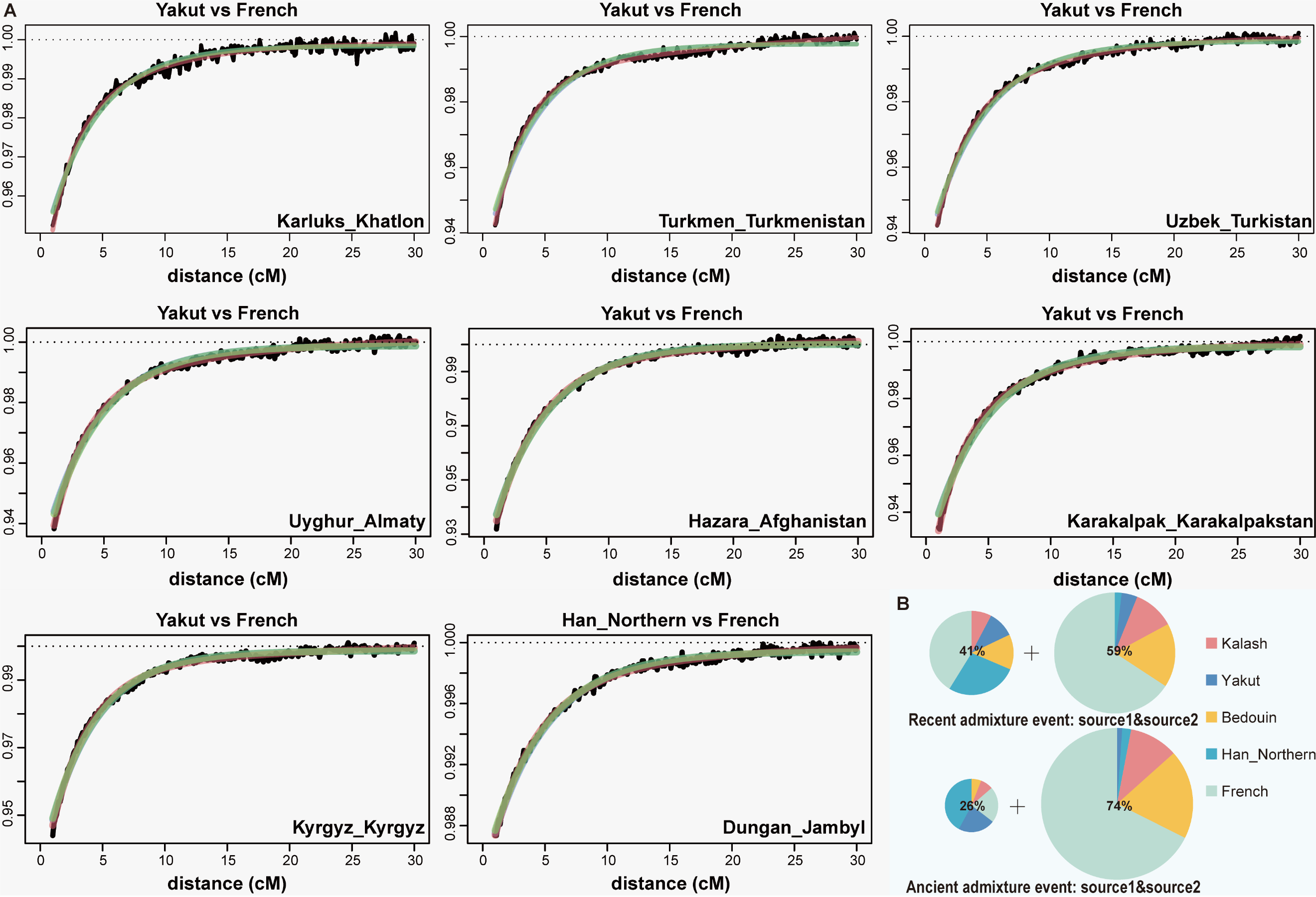

**Figure S13. The ancestry patterns of CAAH revealed via haplotype-based fastGLOBETROTTER.** (**A**), Admixture probability curves for CAAH except for TJK. The black line represents the scaled probability, the green line shows the fitted model assuming a single wave of admixture, the cyan line shows the fitted model assuming a single wave of admixture between two source populations, and the red line shows the fitted model assuming two waves of admixture with different dates. (**B**), The ancestry profile of TJK estimated with the original surrogate populations.

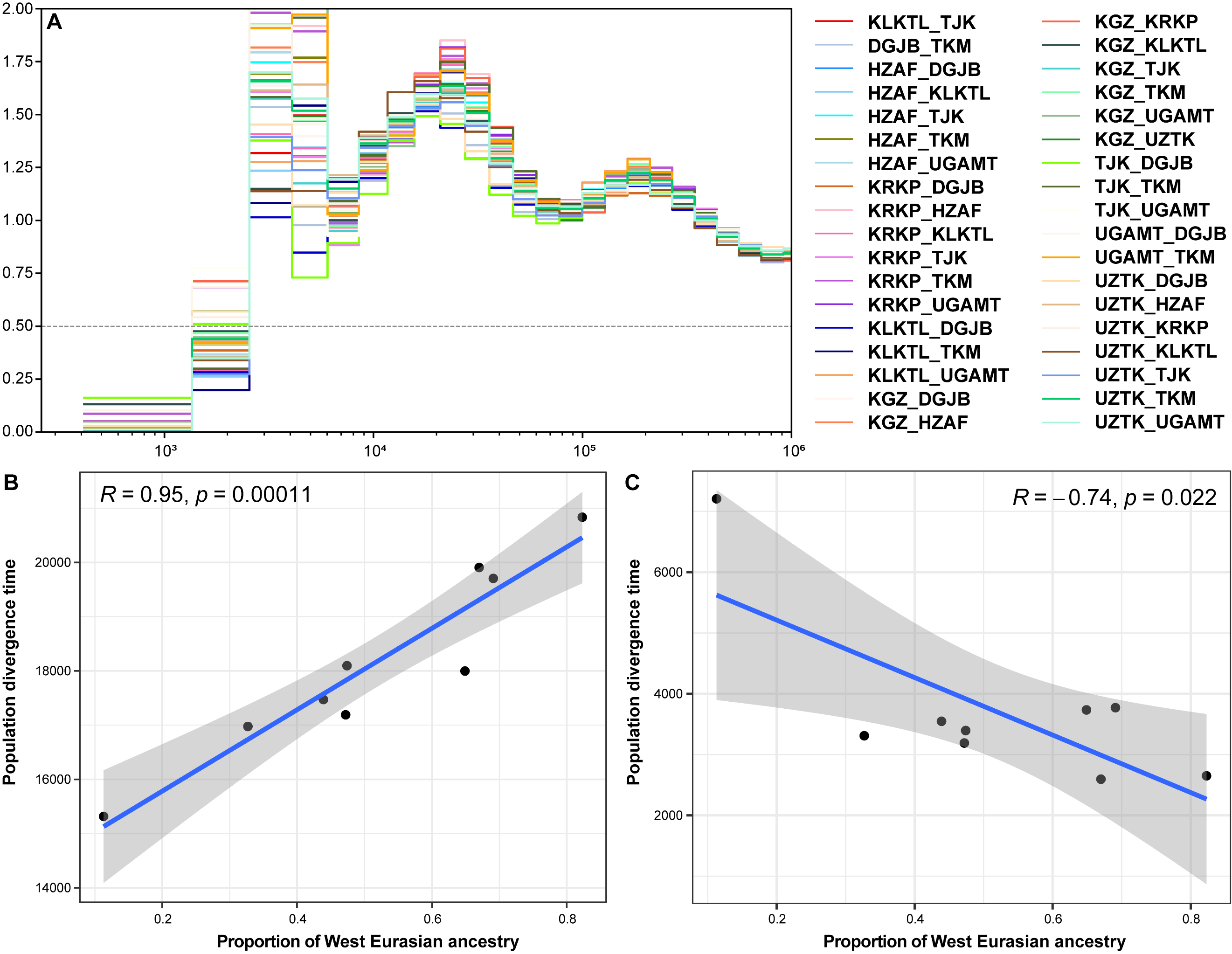

**Figure S14. Population divergence times among CAAH groups.** (**A**), The inferred population split times of pairwise CAAH groups. (**B**), The correlation between the estimated proportion of West Eurasian ancestry and population divergence times between Northern Han Chinese and CAAH groups.

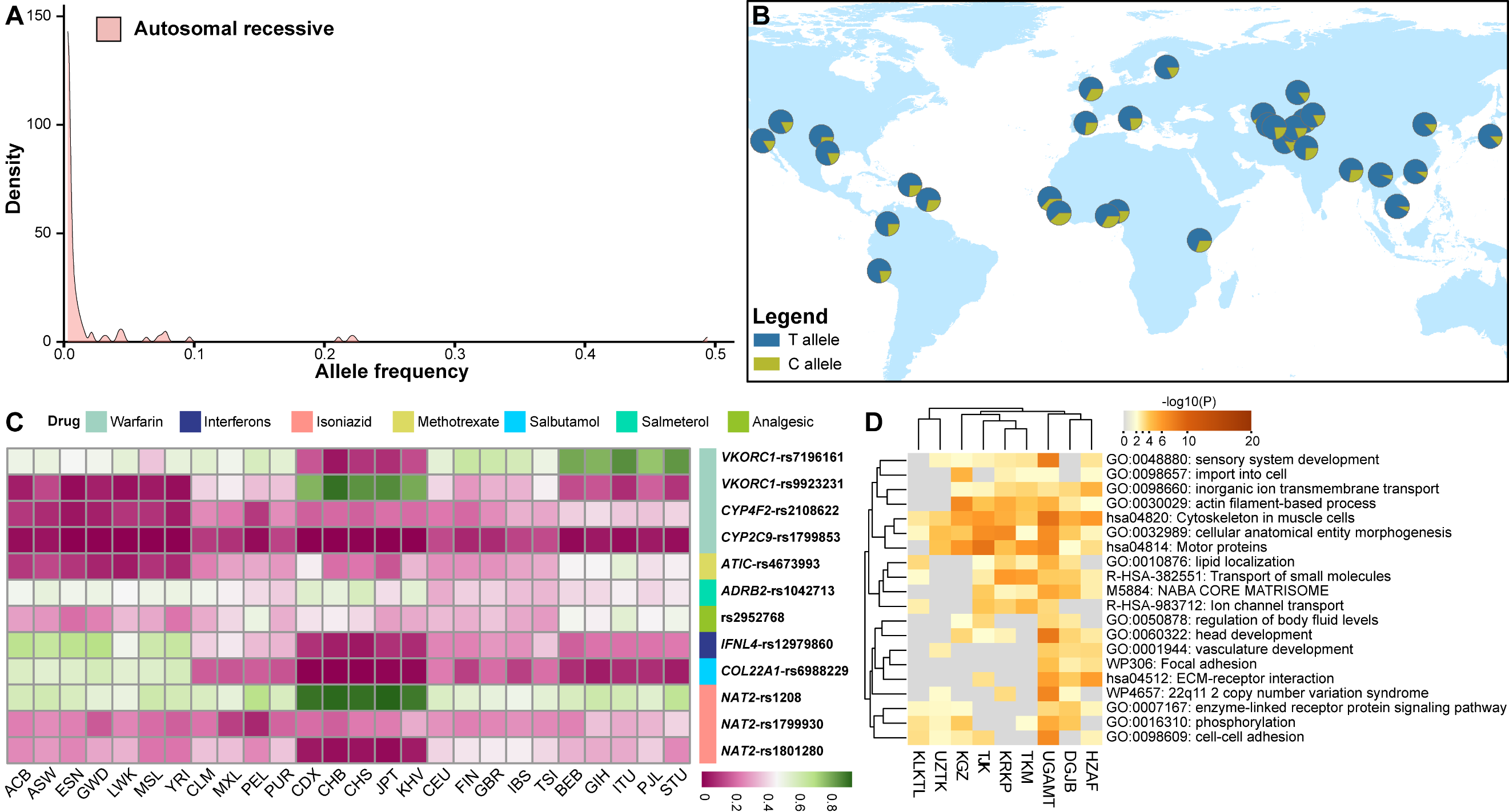

**Figure S15. The distribution of medically relevant and pharmacogenomic variants.** (**A**), Density plot of frequencies of pathogenic and likely pathogenic ClinVar variants, inherited in an autosomal recessive manner. (**B**), Allele frequency distribution of *SCN5A*-A1673G (rs1805124, H558R) variant. (**C**), Detailed allele frequencies of known pharmacogenomic variants identified in CAAH. ACB: African Caribbean in Barbados, ASW: African Ancestry in Southwest US, ESN: Esan in Nigeria, GWD: Gambian in Western Division-Mandinka, LWK: Luhya in Webuye, Kenya, MSL: Mende in Sierra Leone, YRI: Yoruba in Ibadan, Nigeria, CLM: Colombian in Medellin, Colombia, MXL: Mexican Ancestry in Los Angeles, California, PEL: Peruvian in Lima, Peru, PUR: Puerto Rican in Puerto Rico, CDX: Xishuangbanna Dai Chinese, CHB: Beijing Han Chinese, CHS: Southern Han Chinese, JPT: Tokyo Japanese, KHV: Ho Chi Minh Kinh, CEU: Utah residents with Northern and Western European ancestry, FIN: Finnish in Finland, GBR: British in England and Scotland, IBS: Iberian populations in Spain, TSI: Toscani in Italy, BEB: Bengali in Bangladesh, GIH: Gujarati Indian in Houston, Texas, ITU: Indian Telugu in the UK, PJL: Punjabi in Lahore, Pakistan, STU: Sri Lankan Tamil in the UK. (**D**), Enrichment results of heterozygous protein-truncating variants identified in CAAH.

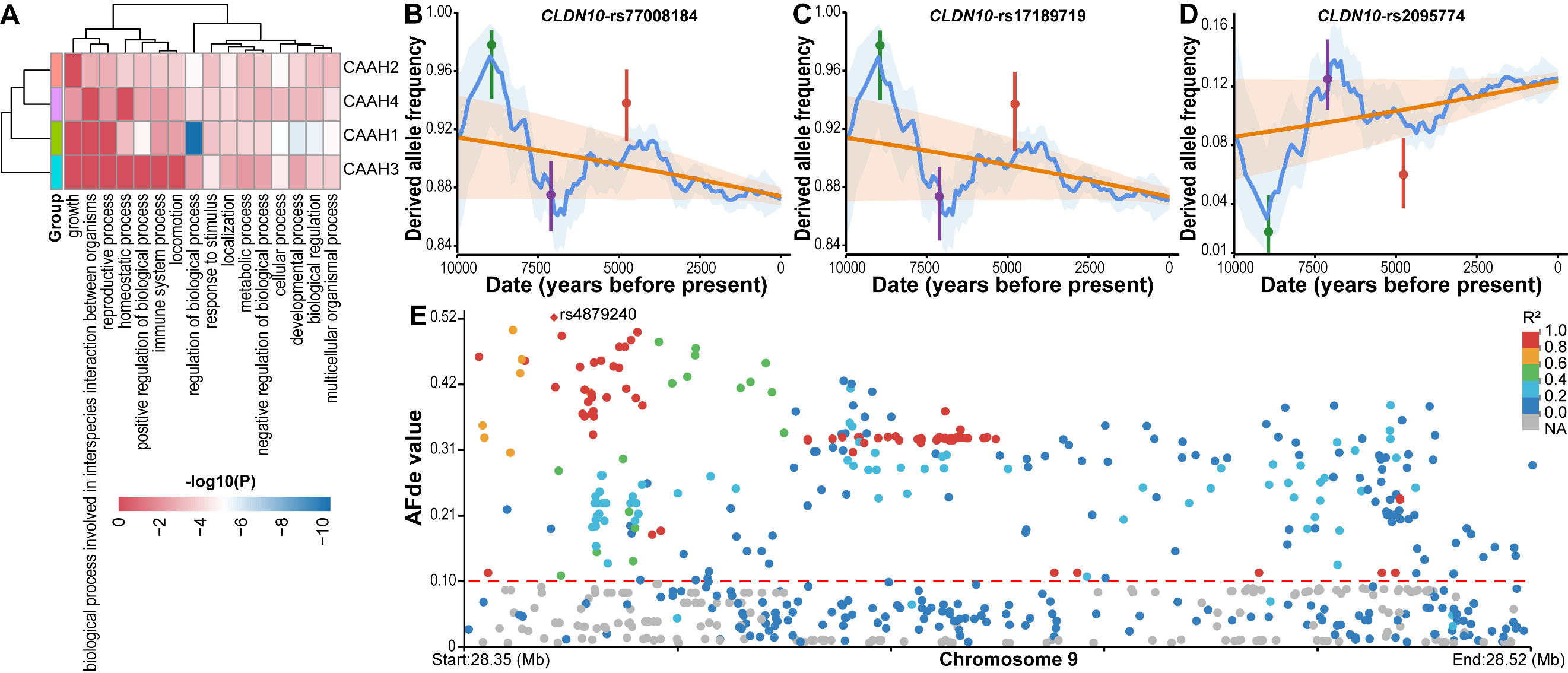

**Figure S16. The heatmap focuses on parent Gene Ontology (GO) terms, the trajectory of derived allele frequencies (DAFs) of *GLDN10* variants, and linkage patterns of variants in *LINGO2*.** (**A**), The heatmap presents parent GO terms associated with candidate signatures of local adaptation. The DAF trajectory over time for rs77008184 (**B**), rs17189719 (**C**), and rs2095774 (**D**) in the *GLDN10* gene. (**E**), Linkage patterns of variants within the *LINGO2* gene, with rs4879240 exhibiting the highest AFd_e_ in natural selection signals within this region as the focal locus. The positions of the variants in **Fig. S15E** are the same as in **Fig. 5E**. Different degrees of linkage disequilibrium (LD) are represented by various colors, with red (R² = 1) indicating complete LD. AFd_e_: the deviation between observed and expected allele frequencies.

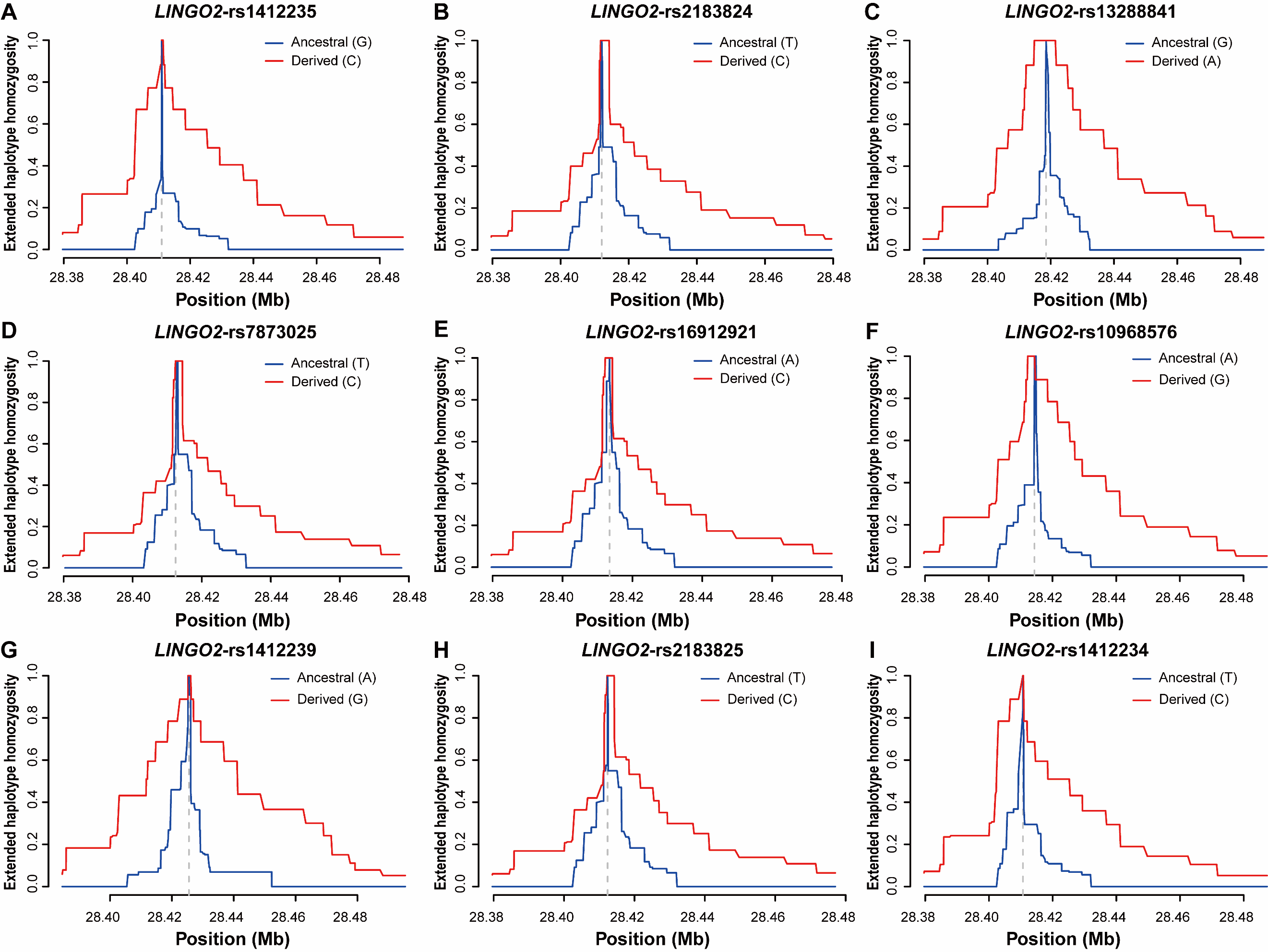

**Figure S17. Extended haplotype homozygosity (EHH) curves of *LINGO2* variants.** The EHH curves of rs1412235 (**A**), rs2183824 (**B**), rs13288841 (**C**), rs7873025 (**D**), rs16912921 (**E**), rs10968576 (**F**), rs1412239 (**G**), rs2183825 (**H**), and rs1412234 (**I**) in the *LINGO2* gene.

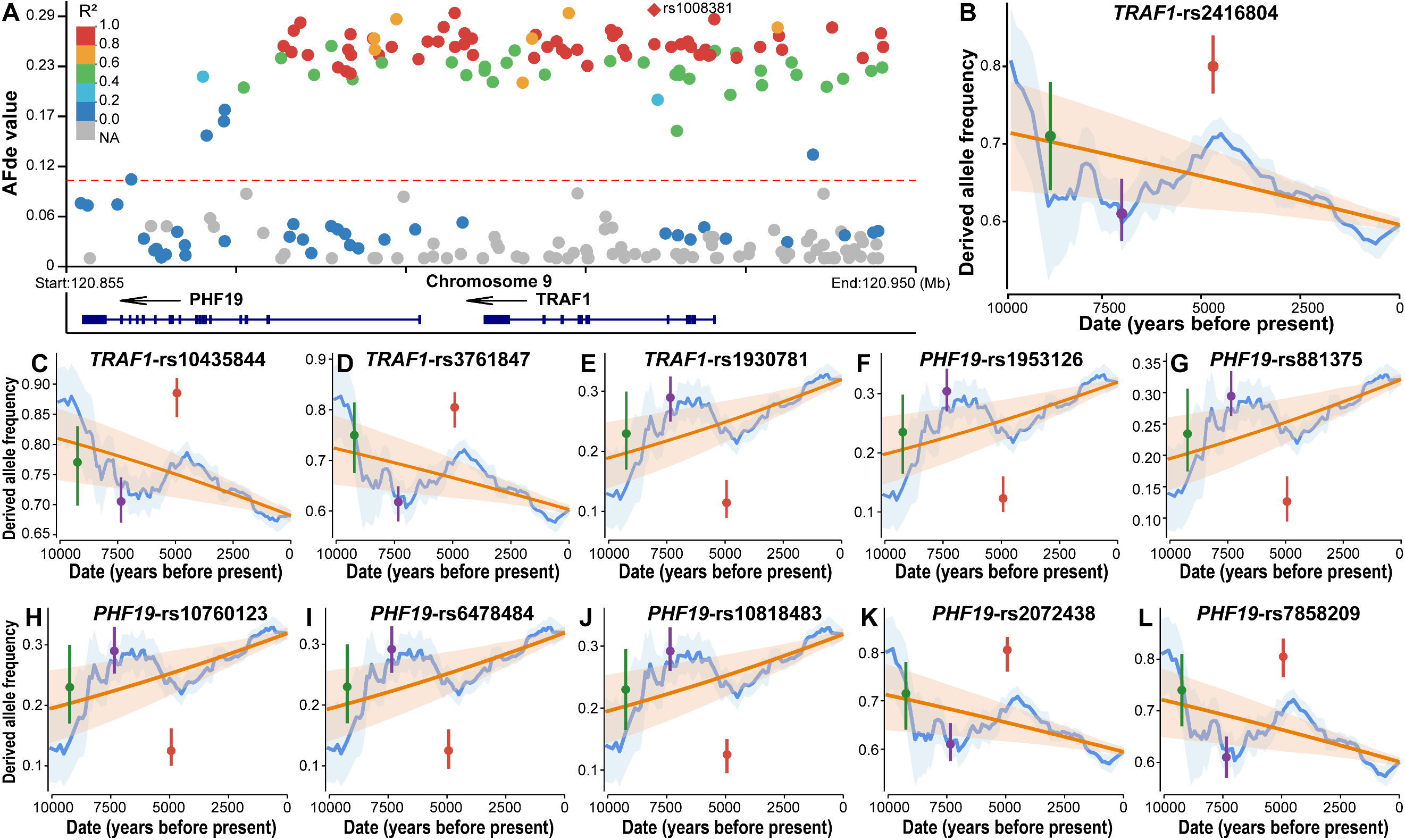

**Figure S18. Linkage patterns of variants in the *TRAF1* and *PHF19* genes and trajectories of DAFs of *TRAF1*/*PHF19* variants.** (**A**), Linkage patterns of variants within the *TRAF1* and *PHF19* genes, with rs1008381 exhibiting the highest AFd_e_ in natural selection signals within this region as the focal locus. The positions of the variants in **Fig. S17A** are the same as in **Fig. 5F**. Different degrees of LD are represented by various colors, with red (R² = 1) indicating complete LD. The DAF trajectory over time for rs2416804 (**B**), rs10435844 (**C**), rs3761847 (**D**), and rs1930781 (**E**) in the *TRAF1* gene. The DAF trajectory over time for rs1953126 (**F**), rs881375 (**G**), rs10760123 (**H**), rs6478484 (**I**), rs10818483 (**J**), rs2072438 (**K**), and rs7858209 (**L**) in the *PHF19* gene.

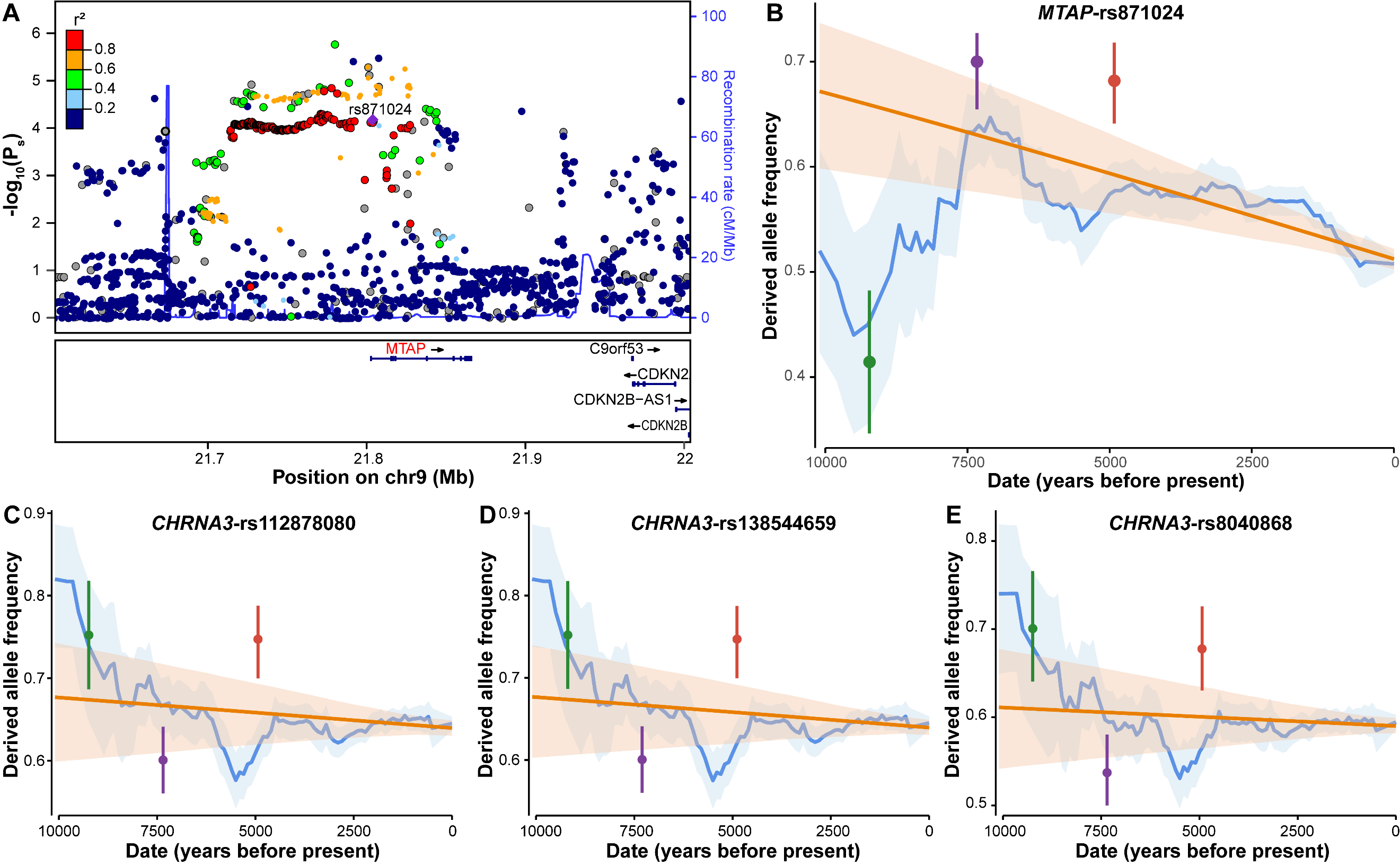

**Figure S19. The selection plot and linkage patterns of variants in the *MTAP* gene, and trajectories of DAFs of *MTAP/CHRNA3* variants.** (**A**), Selection plot of *MTAP* variants and linkage patterns of *MTAP*-rs871024 identified in CAAH2 with adjacent loci. The plot was generated using selection X-statistic values derived from ancient West Eurasian populations (Harvard Dataverse: https://doi.org/10.7910/DVN/7RVV9N). The DAF trajectory over time for *MTAP*-rs871024 (**B**), *CHRNA3*-rs112878080 (**C**), *CHRNA3*-rs138544659 (**D**), and *CHRNA3*-rs8040868 (**E**).

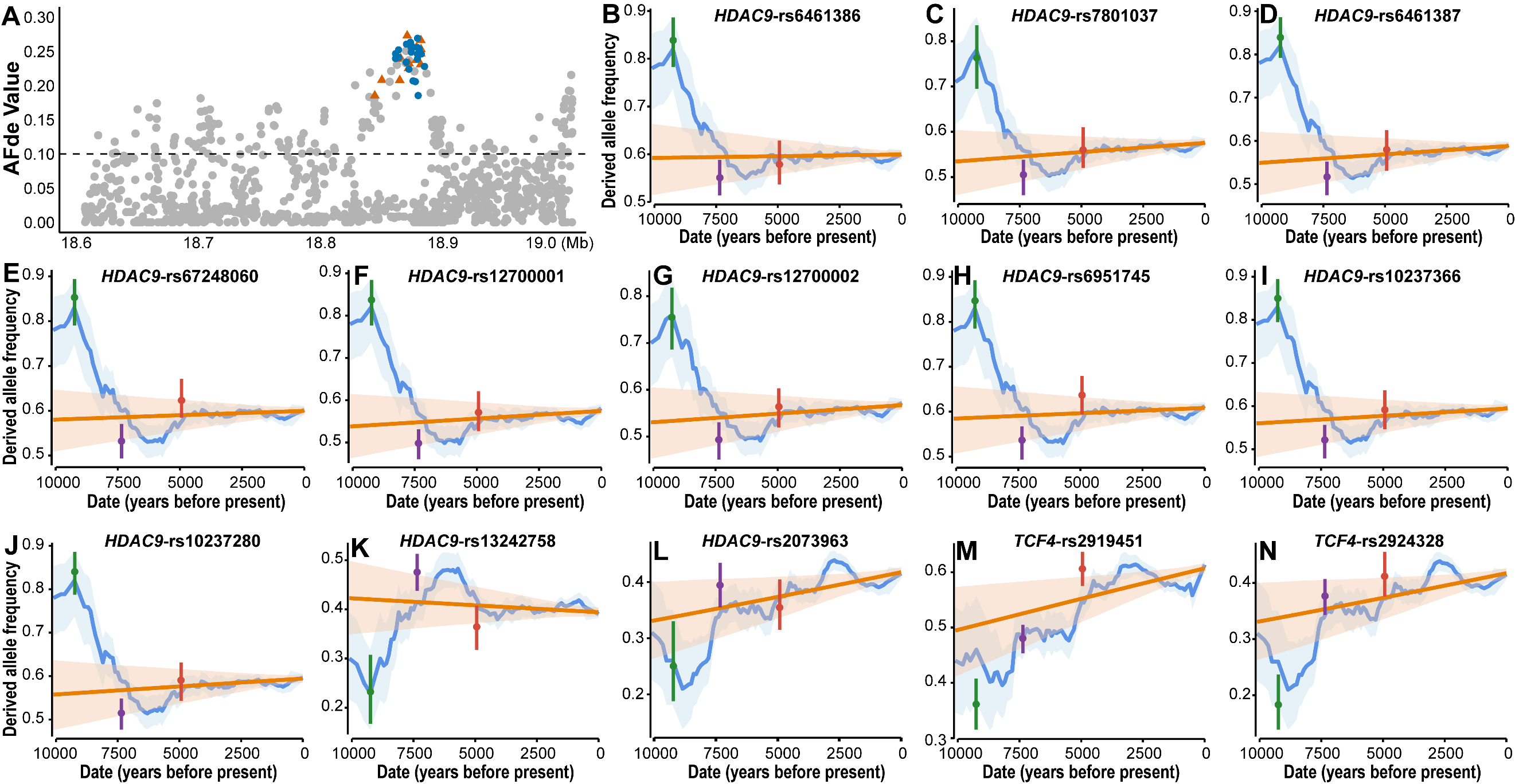

**Figure S20. The AFd_e_ distribution of variants within the *HDAC9* gene and trajectory of DAFs of variants in the *HDAC9* and *TCF4* genes.** (**A**), The AFd_e_ distribution of variants between 18.6 and 19.0 Mb within the *HDAC9* gene. Variants highlighted in blue indicate natural selection signals without reported phenotypic effects, whereas those in orange represent natural selection signals with phenotypic effects documented in the GWAS Catalog. The DAF trajectory over time for rs6461386 (**B**), rs7801037 (**C**), rs6461387 (**D**), rs67248060 (**E**), rs12700001 (**F**), rs12700002 (**G**), rs6951745 (**H**), rs10237366 (**I**), rs10237280 (**J**), rs13242758 (**K**), rs2073963 (**L**) in the *HDAC9* gene, *TCF4*-rs2919451 (**M**), and *TCF4*-rs2924328 (**N**).

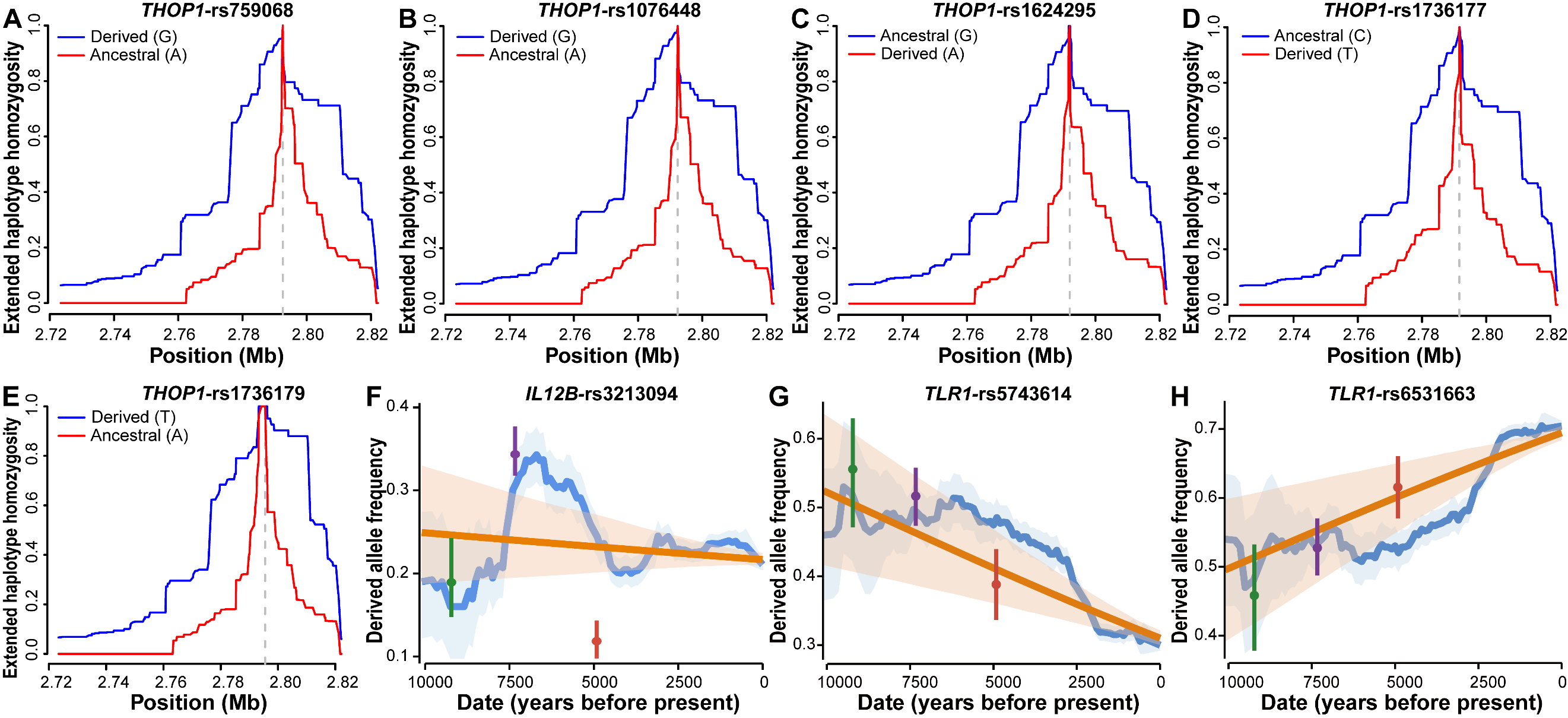

**Figure S21. EHH curves of *THOP1* variants and DAF trajectories of variants in the *IL12B* and *TLR1* genes.** The EHH curves of rs759068 (**A**), rs1076448 (**B**), rs1624295 (**C**), rs1736177 (**D**), and rs1736179 (**E**) in the *THOP1* gene. The DAF trajectory over time for *IL12B*-rs3213094 (**F**), *TLR1*-rs5743614 (**G**), and *TLR1*-rs6531663 (**H**).

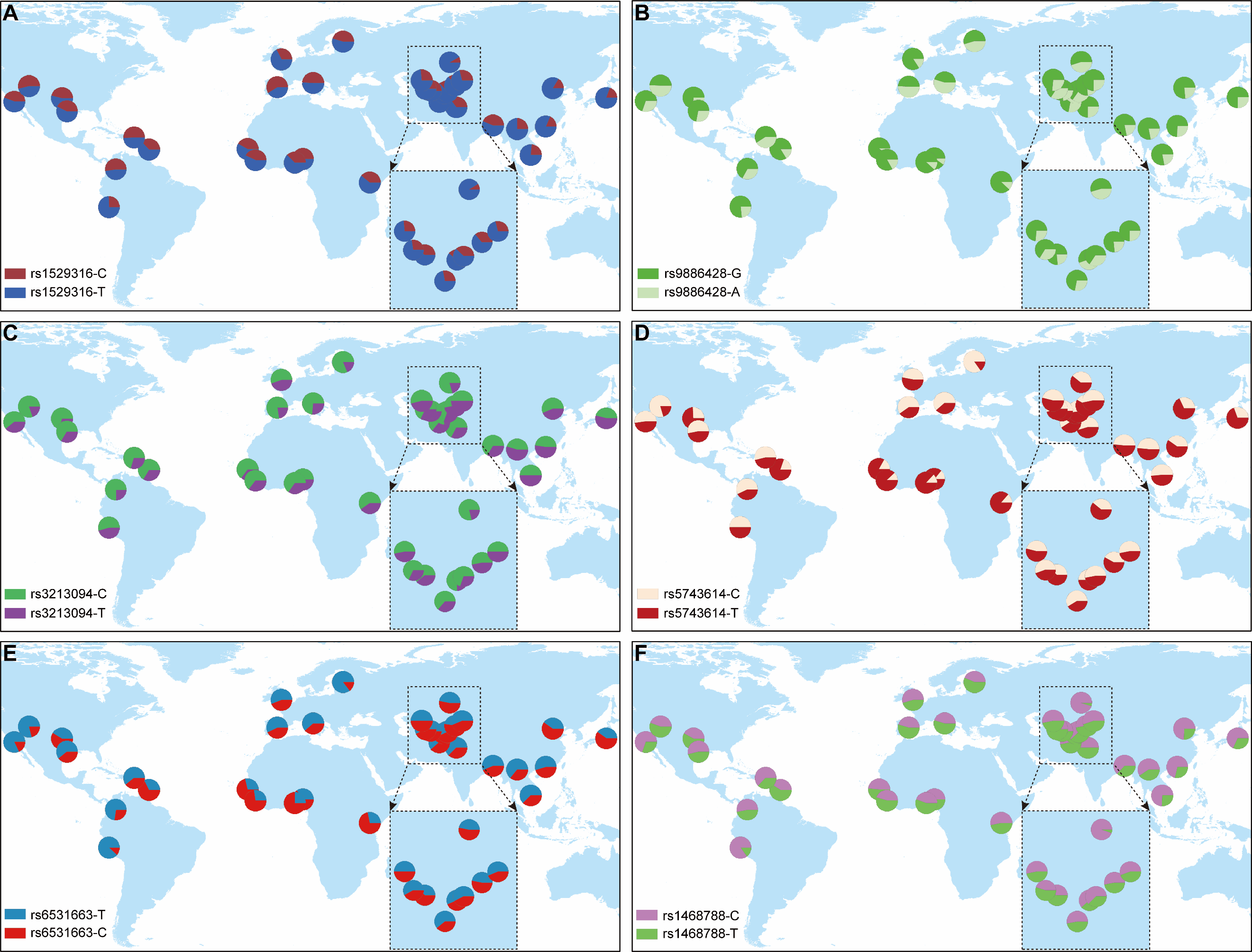

**Figure S22. Geographical distribution of allele frequencies of natural selection signals associated with immune-related conditions.** The allele frequency distributions of *CSMD1*-rs1529316 (**A**), *SGCZ*-rs9886428 (**B**), *IL12B*-rs3213094 (**C**), *TLR1*-rs5743614 (**D**), *TLR1*-rs6531663 (**E**), and *SLC9A4*-rs1468788 (**F**).

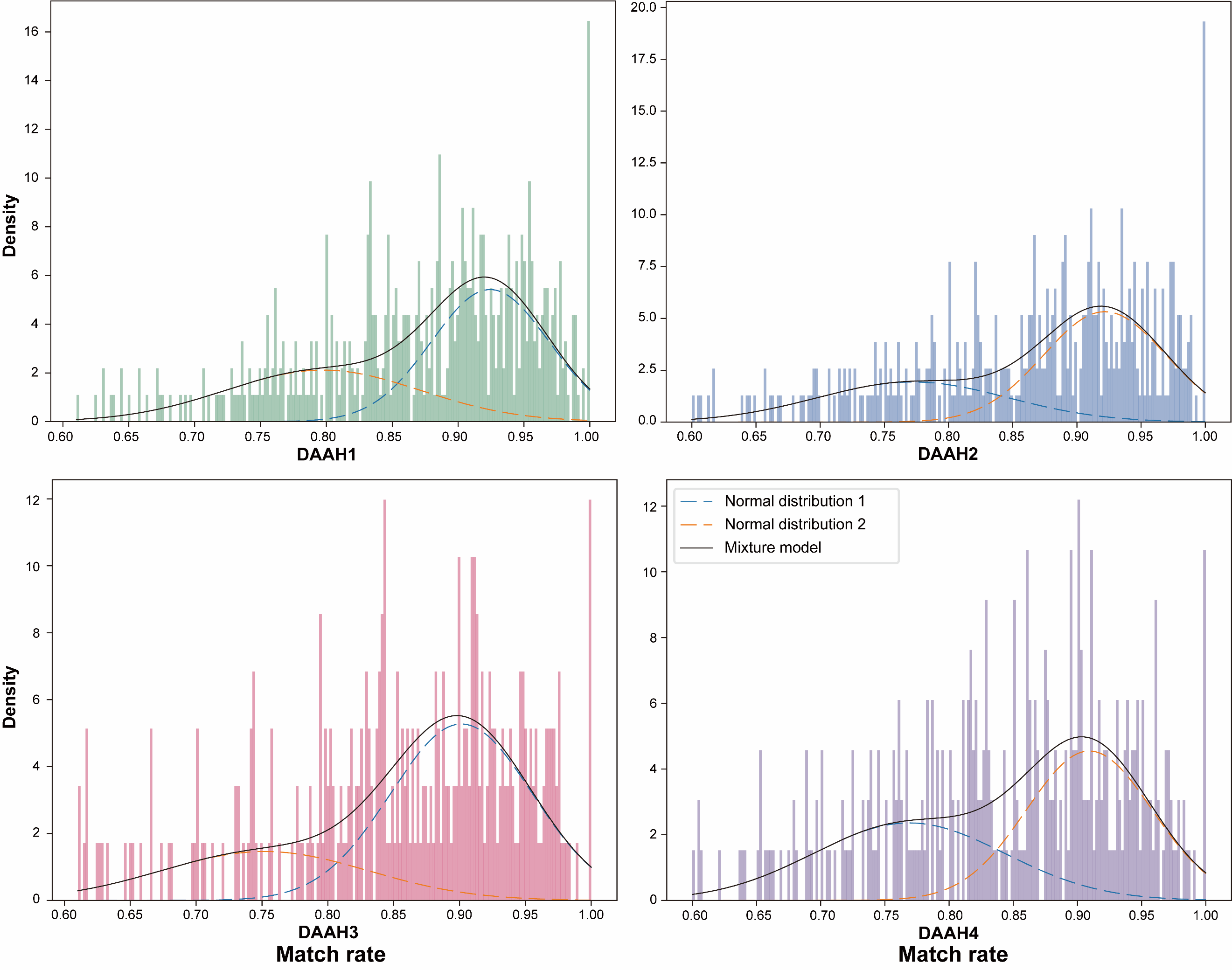

**Figure S23. Density distribution of match rates to the Neanderthal genomes across potentially introgressed segments for each CAAH group.**

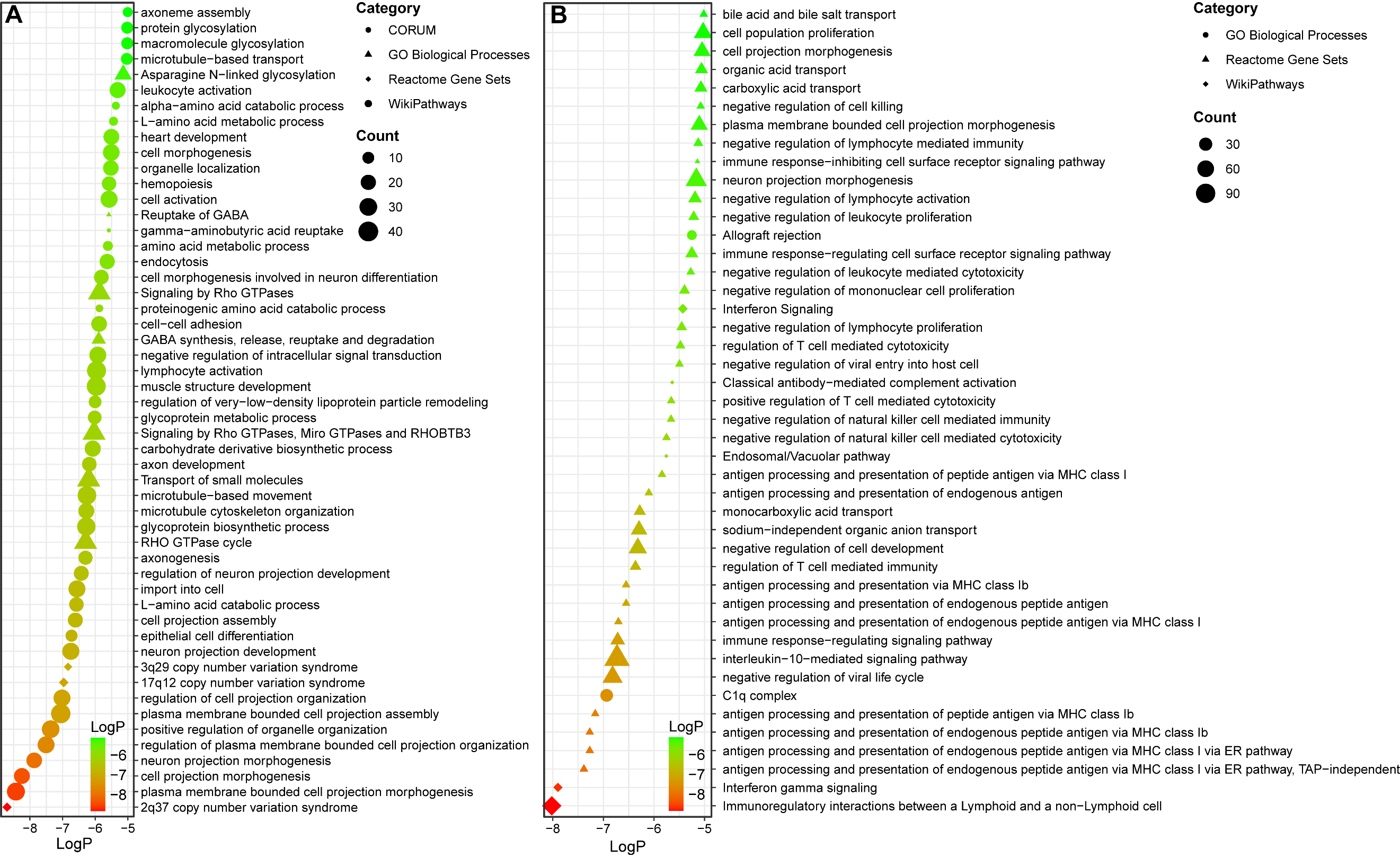

**Figure S24. The bubble chart shows GO terms that have been selected for analysis.** The significant biological pathways obtained based on the Neanderthal- (**A**) and Denisovan-derived sequences (**B**). Biological pathways with FDR < 0.05 and log10(P-value) < -5 were selected for bubble plot generation.
